# Sex-specific developmental scales improve early childhood developmental surveillance

**DOI:** 10.1101/2023.04.12.23288336

**Authors:** Tamar Sudry, Guy Amit, Deena R Zimmerman, Meytal Avgil Tsadok, Ravit Baruch, Hadar Yardeni, Pinchas Akiva, Dror Ben Moshe, Eitan Bachmat, Yair Sadaka

## Abstract

**Introduction:** Routine developmental surveillance is fundamental for timely identification of developmental delays. We explored sex-related differences in milestone attainment rate and evaluated the clinical need for sex-specific developmental scales.

**Methods:** This is a retrospective cross-sectional study, utilizing data from a national child surveillance program. The study included children from birth to six years of age, assessed between 2014-2021 (n=643,958 and n=309,181 for the main and validation cohorts, respectively).

We measured the differences between sexes in normative attainment age of 59 milestones from four developmental domains and calculated the projected error rates when conducting unified vs. sex-specific surveillance.

**Results:** Girls preceded boys in most milestones of all domains. Conducting developmental surveillance using unified rather than sex-specific scales resulted in potential missing of girls at risk of developmental delay (19.3% of failed assessments), and false alerts for boys (5.9%).

**Conclusion:** These findings suggest that using sex-specific scales may improve the accuracy of early childhood developmental surveillance.

## Introduction

Developmental surveillance is a longitudinal process that involves taking a developmental history based on milestone attainment age, observing milestones and other behaviors, examining the child, and applying clinical judgment during health supervision visits^1^. It is used worldwide by pediatricians and healthcare providers at routine encounters, as well as by teachers and parents for evaluating the strengths and weaknesses of each child, helping parents choose relevant exposures for their children, and identifying children in need of further follow-up^2–7^. The importance of developmental surveillance tools was recently stressed by the Center for Disease Control and Prevention (CDC) and the American Academy of Pediatrics (AAP), who convened a panel of subject matter experts to update the CDC’s checklists of developmental milestones. This recent work pointed out the need to establish an evidence-based milestones surveillance scale to improve developmental surveillance^1^. Currently, developmental surveillance tools are applied identically to both sexes^8–12^, although boys and girls exhibit differences in various aspects, including growth and neurodevelopment^13,14^. Attempts to describe and explain the magnitude and extent of these sex-related variations pose a scientific and medical challenge^15^. Nevertheless, there is an increasing body of evidence suggesting possible differences between the sexes in specific domains of child development^16–19^. Recent studies suggest that girls exhibit some advantage in language and communication skills^20–26^ compared to boys, as well as in personal-social and fine motor skills^27–29^. The data regarding gross motor developmental differences are controversial: some studies concluded that there are no sex-related differences^30–32^, while other studies suggested that boys outperform girls^10,29^ or, contradictingly, that girls precede boys^33,34^. Most of these studies reviewed a relatively small sample of the population and often defined the observed differences as clinically insignificant. Although there is accumulating evidence indicating sex-related differences in the rate of skill acquisition during early childhood, the possible need for different sex-specific scales for developmental surveillance has not been considered previously.

The aim of this nationwide study is to quantitatively describe sex-related differences in milestones attainment rate during early childhood years, and accordingly assess the clinical need for separate sex-specific scales for developmental surveillance. To establish and compare separate scales for boys and girls, we analyzed 59 milestones of the recently introduced Tipat Halav Israeli Screening (THIS) developmental scale^35^, an evidence-based national developmental surveillance scale, based on more than 3.7 million developmental assessments of over 640,000 children aged 0-6 years. In the current study we used the original dataset of these developmental assessments to construct separate sex-specific scales, and calculated, using a new validation dataset, the projected error rate when conducting unified rather than sex-specific developmental surveillance.

## Methods

### Data collection

Developmental assessments of children included in this study were performed in Israel between January 2014 and September 2021. Data was collected and analyzed as described in detail elsewhere^36^. Briefly, developmental surveillance (ages birth to 6 years) in Israel is performed routinely according to national standards by trained public health nurses in approximately 1,000 maternal child health clinics (MCHC). Collected data of approximately 70% of the Israeli population of this age group is documented in a single common database. The developmental assessments^35^ include 59 milestones across four domains: personal-social, language, fine motor, and gross motor. Description of these milestones can be found in supplementary eTable 1. During each visit, a predefined group of age-related milestones is evaluated by the nurses. The child’s performance is documented within an electronic medical record (EMR).

### Study cohorts

The main study cohort consisted of children born between January 1^st^ 2014 and September 1^st^ 2020 who were followed at the MCHCs that used the common database. All children with at least one developmental evaluation record during the study period were included in this study. Preterm newborn (gestational age < 37 weeks), low birth weight (<2.5 kg), abnormal weight measurement (<3% according to standardized child growth charts), abnormal head circumference measurement (microcephaly < 3% or macrocephaly > 97% according to standardized child growth charts) were excluded, as well as infants with missing gestational age, visits without developmental data or without the child’s age. The main cohort was used to establish and compare the developmental norms of boys and girls.

A secondary validation cohort of all MCHC visits between October 2020 and October 2021 was used to assess the effect of sex-specific scales on developmental surveillance results.

### Determining developmental norms

For each child, the first evaluation of each milestone was included in the analysis, to avoid a potential bias towards children with developmental delays.

Developmental norms for each milestone were established separately for boys and girls, using the main cohort data, following the approach described in our previously published study^36^. Briefly, children were grouped by their age during milestone evaluation and the achievement rate at each age was calculated from the empirical data. The obtained curve of achievement rate by age was then smoothed and interpolated, and the threshold ages at which the achievement rate surpassed 75%, 90% and 95% were derived.

### Comparison of boys and girls developmental norms

To explore the sex differences in milestones attainment age, we compared the milestone norms of boys and girls. THIS developmental scale originally included 59 milestones. Among them, 17 milestones exceeded 95% success rate for both sexes during their initial evaluation. Therefore, for these milestones only the initial attainment age was calculated, however a threshold comparison was not conducted. For the remaining 42 milestones, we compared the threshold ages for milestone attainment by 75%, 90% and 95% of both sexes.

Formally, we denote by 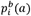 and 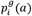 the proportion of attaining milestone *i* at age *a* by boys and girls, respectively. We denote the threshold ages of milestone *i’s* attainment by 75%, 90%, and 95% of the boys by 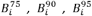 respectively, such that 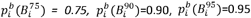. Similarly, the threshold ages for the girls are denoted by 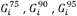.

For each milestone *i*, we examined the differences between the threshold ages for boys and girls, denoted by: 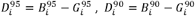 and 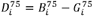.

We applied a two-proportion z-test to assess the significance of the difference between the success rates of boys and girls at the earlier threshold age. If 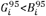, we test the that 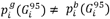, and otherwise we test whether 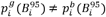. The notations used are illustrated in supplementary eFigure 1.

We compared the sex-specific threshold ages to those of the unified THIS scale. A milestone with a clinically significant change was considered as any milestone for which the attainment age by ≥90% of the children changed by at least one month.

Data analysis was performed using Python version 3.6.

### The effect of sex-specific scales on developmental surveillance results

Conducting routine sex-specific developmental surveillance may result in earlier identification of children at risk for developmental delay, while reducing unnecessary referral to further developmental workup of others. We examined the potential effect of utilizing separate sex-specific scales by assessing the disagreements between the unified THIS scale and the sex-specific scales (denoted *THIS-Boys, THIS-Girls*). For each visit with an unattained milestone, the failure to achieve the milestone was classified as ‘suspected delay’ if the age of the child was greater or equal to the threshold age at which 90% of the children achieved the milestone. The milestones of each visit were grouped by the four domains and the visit was labeled as ‘suspected delay’ or ‘no suspected delay’ by the labels of the milestones of each domain. We then calculated the false-positive and false-negative rates of the visit labeling by the original THIS scale, compared to the ‘ground-truth’ labeling obtained by applying the thresholds of the sex-specific norms 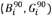. The calculation was performed on both main and validation cohorts.

### Analysis of the effect of sex on attaining developmental milestones

We performed multivariable logistic regression analysis to assess the effect of the sex-group variable on the probability of failing to attain a developmental milestone while controlling for socio-demographic variables that may be associated with the outcome. For each milestone, we identified the age at which the largest number of children were evaluated and selected all evaluations within a time window around that age (±2 weeks for age>24 months, ±1 week age≤24 months). We fitted an L1-regularized logistic regression model to predict the probability of not attaining the milestone, with the child’s sex as a single variable (unadjusted) and with adjustments for additional variables, including the mother’s ethnicity, country of birth, education level, marital status, and age group. We calculated the unadjusted odds ratio of failing to attain the milestone by boys, as well as the adjusted odds ratio while controlling for the additional predictor variables. Missing values in categorical socio-demographic variables were handled as a distinct category. Binary variables for which data was missing were set to 0.

### Ethics declarations

All analyses were carried out in accordance with relevant guidelines and regulations. The study protocol was approved by the Soroka University Medical Center institutional ethical committee and was conducted in accordance with the principles of the Declaration of Helsinki (MHC-0006-18).

## Data and code availability statement

The de-identified patient-level data used for this study contains sensitive information and therefore is not available outside the secured research environment of the Israel Ministry of Health. Summary aggregate level data and analysis code for this study can be made available upon reasonable request to the corresponding author.

## Results

### Study population

The flowchart of study participants is shown in Figure 1. For the main cohort, the electronic medical records of 839,574 children who visited the MCHCs from January 2014 until September 2020 were extracted. Following the exclusion of children with abnormal developmental potential or missing information, as described in the methods, a total of 643,958 children, 319,562 girls, and 324,396 boys, with 3,774,517 developmental evaluations were available for the analysis.

**Figure 1.**
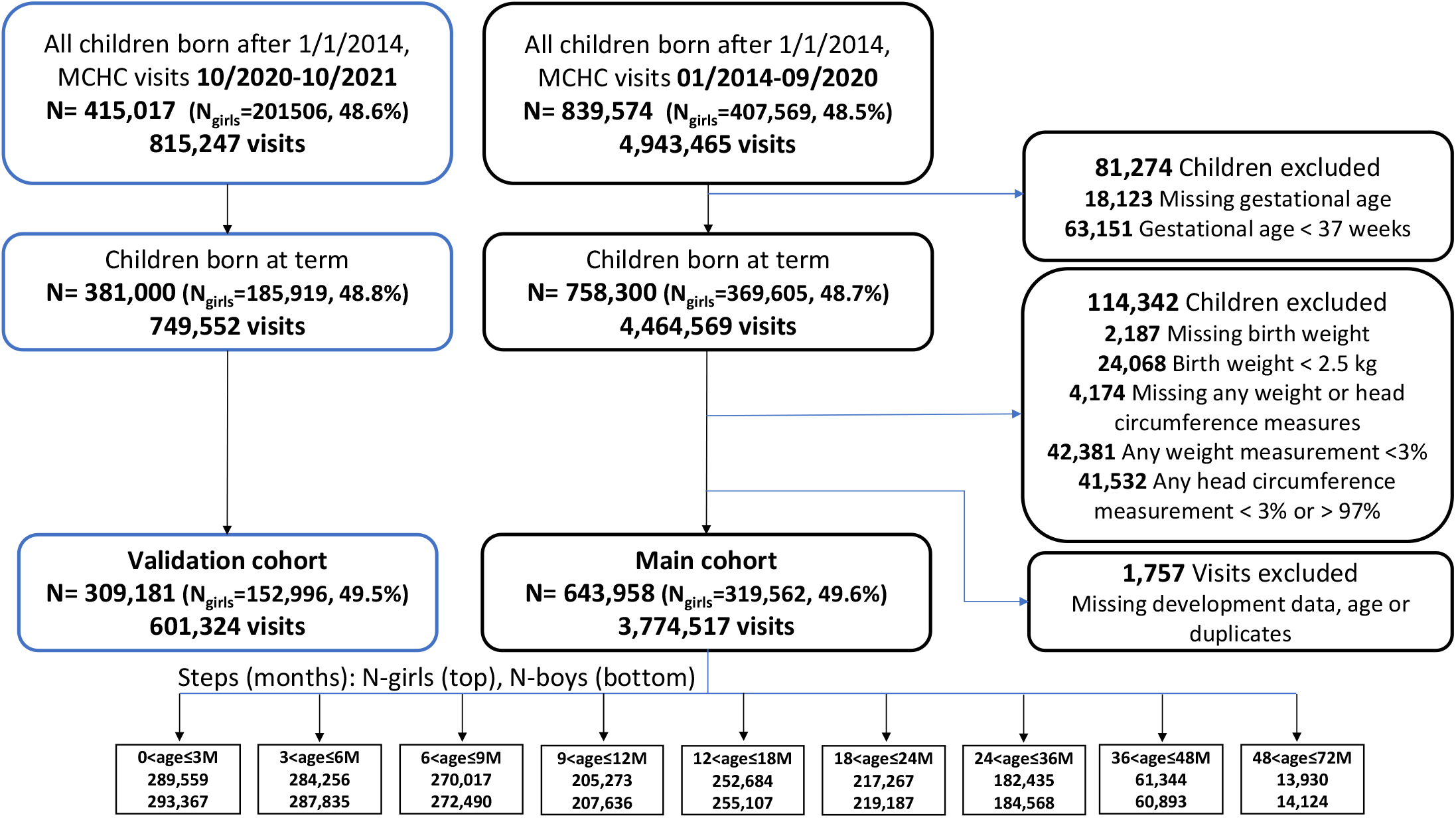
study population. Study population included all children born between January 2014 to September 2021, who visited maternal child health clinics. Children with abnormal developmental potential or missing data were excluded. The main cohort included 3,774,517 developmental assessment visits of 643,958 children (319,562 girls, 49.6%) between January 2014 and September 2020. The validation cohort included additional 601,324 visits of 309,181 children (152,996 girls, 49.5%) between October 2020 and October 2021.

The validation cohort consisted of additional 601,324 developmental evaluations from 309,181 children (49.6% girls), performed between October 2020 and October 2021. The exclusion criteria used for the validation cohort were identical to the main cohort.

To assess potential selection bias, the socio-economic characteristics of the cohort population, which consists of about 70% of all children born in Israel during the study period, were compared to the general population of children born in Israel as documented by the Israeli Central Bureau of Statistics^36^ and no major differences were observed^35^.

The baseline demographic, birth, and maternal characteristics for boys and girls in the two cohorts are presented in Table 1. Both groups had similar gestational and neonatal characteristics and were composed of a nearly equal distribution of ethnic groups and maternal features. Approximately one-third of the mothers had an academic degree and a quarter had high-school education. Most mothers were married (>87%), working (>44%), and born in Israel (>88%).

**Table 1.** Study population characteristics of the main and validation (Val) cohorts.

| Cohort characteristics |  | Main - All | Main - Girls | Main - Boys | Val. - All | Val. - Girls | Val. - Boys |
| --- | --- | --- | --- | --- | --- | --- | --- |
| n |  | 643958 | 319562 | 324396 | 309181 | 152996 | 156185 |
| Visits per child (SD) |  | 5.86 (2.4) | 5.85 (2.3) | 5.87 (2.4) | 1.95 (1.1) | 1.94 (1.1) | 1.95 (1.1) |
| Ethnic group, n (%) | **Arab** | 140525 (21.8) | 69546 (21.8) | 70979 (21.9) | 77259 (25.0) | 38090 (24.9) | 39169 (25.1) |
|  | **Druse** | 11796 (1.8) | 5812 (1.8) | 5984 (1.8) | 6659 (2.2) | 3252 (2.1) | 3407 (2.2) |
|  | **Jewish** | 392634 (61.0) | 195353 (61.1) | 197281 (60.8) | 185195 (59.9) | 91944 (60.1) | 93251 (59.7) |
|  | **Missing** | 76333 (11.9) | 37687 (11.8) | 38646 (11.9) | 28478 (9.2) | 14035 (9.2) | 14443 (9.2) |
|  | **Other** | 22670 (3.5) | 11164 (3.5) | 11506 (3.5) | 11590 (3.7) | 5675 (3.7) | 5915 (3.8) |
| Mother birth country, n (%) | **Israel** | 528861 (82.1) | 262613 (82.2) | 266248 (82.1) | 252729 (81.7) | 125121 (81.8) | 127608 (81.7) |
|  | **Other** | 58983 (9.2) | 29125 (9.1) | 29858 (9.2) | 28399 (9.2) | 13935 (9.1) | 14464 (9.3) |
|  | **Europe** | 25180 (3.9) | 12472 (3.9) | 12708 (3.9) | 12428 (4.0) | 6165 (4.0) | 6263 (4.0) |
|  | **Former Soviet Un.** | 22240 (3.5) | 11011 (3.4) | 11229 (3.5) | 10914 (3.5) | 5403 (3.5) | 5511 (3.5) |
|  | **Ethiopia** | 8644 (1.3) | 4313 (1.3) | 4331 (1.3) | 4690 (1.5) | 2361 (1.5) | 2329 (1.5) |
|  | **Missing** | 50 (0.0) | 28 (0.0) | 22 (0.0) | 21 (0.0) | 11 (0.0) | 10 (0.0) |
| Employment status, n (%) | **Missing** | 199544 (31.0) | 98638 (30.9) | 100906 (31.1) | 80165 (25.9) | 39823 (26.0) | 40342 (25.8) |
|  | **Not Working** | 131703 (20.5) | 65436 (20.5) | 66267 (20.4) | 61389 (19.9) | 30521 (19.9) | 30868 (19.8) |
|  | **Student** | 28514 (4.4) | 14100 (4.4) | 14414 (4.4) | 14952 (4.8) | 7327 (4.8) | 7625 (4.9) |
|  | **Working** | 284197<br>(44.1) | 141388<br>(44.2) | 142809<br>(44.0) | 152675<br>(49.4) | 75325<br>(49.2) | 77350<br>(49.5) |
| Mother's education level, n (%) | **Academic** | 195799<br>(30.4) | 97369<br>(30.5) | 98430<br>(30.3) | 100347<br>(32.5) | 49727<br>(32.5) | 50620<br>(32.4) |
|  | **Elementary** | 13442<br>(2.1) | 6797<br>(2.1) | 6645<br>(2.0) | 5961<br>(1.9) | 2940<br>(1.9) | 3021<br>(1.9) |
|  | **High School** | 171411<br>(26.6) | 84884<br>(26.6) | 86527<br>(26.7) | 81464<br>(26.3) | 40319<br>(26.4) | 41145<br>(26.3) |
|  | **Missing** | 199021<br>(30.9) | 98751<br>(30.9) | 100270<br>(30.9) | 87448<br>(28.3) | 43367<br>(28.3) | 44081<br>(28.2) |
|  | **Tertiary Education** | 64285<br>(10.0) | 31761<br>(9.9) | 32524<br>(10.0) | 33961<br>(11.0) | 16643<br>(10.9) | 17318<br>(11.1) |
| Mother's family status, n (%) | **Divorced** | 6343<br>(1.0) | 3137<br>(1.0) | 3206<br>(1.0) | 2666<br>(0.9) | 1342<br>(0.9) | 1324 (0.8) |
|  | **Married** | 566747<br>(88.0) | 281432<br>(88.1) | 285315<br>(88.0) | 269206<br>(87.1) | 133362<br>(87.2) | 135844<br>(87.0) |
|  | **Missing** | 43380<br>(6.7) | 21350<br>(6.7) | 22030<br>(6.8) | 23128<br>(7.5) | 11281<br>(7.4) | 11847<br>(7.6) |
|  | **Other** | 27165<br>(4.2) | 13485<br>(4.2) | 13680<br>(4.2) | 14043<br>(4.5) | 6948<br>(4.5) | 7095<br>(4.5) |
|  | **Widower** | 323 (0.1) | 158 (0.0) | 165 (0.1) | 138 (0.0) | 63 (0.0) | 75 (0.0) |
| Consanguinity, n (%) | **no** | 606227<br>(94.1) | 300904<br>(94.2) | 305323<br>(94.1) | 289882<br>(93.8) | 143506<br>(93.8) | 146376<br>(93.7) |
|  | **yes** | 37731<br>(5.9) | 18658<br>(5.8) | 19073<br>(5.9) | 19299<br>(6.2) | 9490<br>(6.2) | 9309<br>(6.3) |
| Pregnancy week, med [Q1,Q3] |  | 39.5<br>[38.6,40.3] | 39.6<br>[39.0,40.3] | 39.5<br>[38.6,40.3] | 39.5<br>[38.6,40.3] | 39.5<br>[38.6,40.3] | 39.5<br>[38.6,40.3] |
| Birth weight, mean (SD) |  | 3.3 (0.4) | 3.3 (0.4) | 3.4 (0.4) | 3.3 (0.4) | 3.3 (0.4) | 3.4 (0.4) |
| Apgar score one minute, n (%) | **<8** | 18862<br>(2.9) | 7971<br>(2.5) | 10891<br>(3.4) | 9018<br>(2.9) | 3729<br>(2.4) | 5289<br>(3.4) |
|  | **>=8** | 613747<br>(95.3) | 306023<br>(95.8) | 307724<br>(94.9) | 294715<br>(95.3) | 146584<br>(95.8) | 148131<br>(94.8) |
|  | **Missing** | 11349<br>(1.8) | 5568<br>(1.7) | 5781<br>(1.8) | 5448<br>(1.8) | 2683<br>(1.8) | 2765 (1.8) |
| Apgar score five minutes, n (%) | **<8** | 4112<br>(0.6) | 1823<br>(0.6) | 2289<br>(0.7) | 1444<br>(0.5) | 588 (0.4) | 856 (0.5) |
|  | **>=8** | 624688<br>(97.0) | 310238<br>(97.1) | 314450<br>(96.9) | 299928<br>(97.0) | 148554<br>(97.1) | 151374<br>(96.9) |
|  | **Missing** | 15158<br>(2.4) | 7501<br>(2.3) | 7657<br>(2.4) | 7809<br>(2.5) | 3854<br>(2.5) | 3955 (2.5) |
| Head circum. (SD) |  | 34.5 (1.3) | 34.2 (1.3) | 34.7 (1.3) | 34.4 (1.3) | 34.2 (1.3) | 34.7 (1.3) |
| Type of birth, n (%) | **Caesarean section** | 92540<br>(15.1) | 43819<br>(14.4) | 48721<br>(15.8) | 48413<br>(15.7) | 22874<br>(15.0) | 25539<br>(16.4) |
|  | **Instrumental** | 34008<br>(5.6) | 14870<br>(4.9) | 19138<br>(6.2) | 18356<br>(5.9) | 8044<br>(5.3) | 10312<br>(6.6) |
|  | **Missing** | 31946<br>(5.0) | 15977<br>(5.0) | 15969<br>(4.9) | 678 (0.2) | 333 (0.2) | 345 (0.2) |
|  | **Spontaneous** | 485464<br>(79.3) | 244896<br>(80.7) | 240568<br>(78.0) | 241734<br>(78.2) | 121745<br>(79.6) | 119989<br>(76.8) |
| Newborn position, n (%) | **Breech** | 14532<br>(2.3) | 7746<br>(2.4) | 6786<br>(2.1) | 7162<br>(2.3) | 3782<br>(2.5) | 3380 (2.2) |
|  | **Head** | 540304<br>(83.9) | 267786<br>(83.8) | 272518<br>(84.0) | 276744<br>(89.5) | 136679<br>(89.3) | 140065<br>(89.7) |
|  | **Missing** | 82832<br>(12.9) | 40942<br>(12.8) | 41890<br>(12.9) | 21955<br>(7.1) | 10933<br>(7.1) | 11022<br>(7.1) |
|  | **Other** | 6290<br>(1.0) | 3088<br>(1.0) | 3202<br>(1.0) | 3320<br>(1.1) | 1602<br>(1.0) | 1718 (1.1) |
| Mother's age, n (%) | **<=20** | 7101<br>(1.1) | 3517<br>(1.1) | 3584<br>(1.1) | 2791<br>(0.9) | 1370<br>(0.9) | 1421 (0.9) |
|  | **>20, <=40** | 584833<br>(90.8) | 290030<br>(90.8) | 294803<br>(90.9) | 281121<br>(90.9) | 139121<br>(90.9) | 142000<br>(90.9) |
|  | **>40** | 45095<br>(7.0) | 22647<br>(7.1) | 22448<br>(6.9) | 19596<br>(6.3) | 9748<br>(6.4) | 9848 (6.3) |
|  | **Missing** | 6929<br>(1.1) | 3368<br>(1.1) | 3561<br>(1.1) | 5673<br>(1.8) | 2757<br>(1.8) | 2916 (1.9) |

### Establishment of sex-specific scales for developmental surveillance

Comparison of milestone achievement age by 90% and 95% of the boys and the girls (denoted 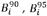 and 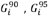 for the boys and girls, respectively), alongside the difference in attainment age between the two groups (denoted 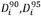), is presented in Figure 2, with milestones grouped by the four developmental domains: language, personal-social, gross motor and fine motor. Overall, girls preceded boys in most of the evaluated milestones of all developmental domains, with an evident gap in attainment age between sexes, which increased at an older age (>12 months).

**Figure 2 (a).**
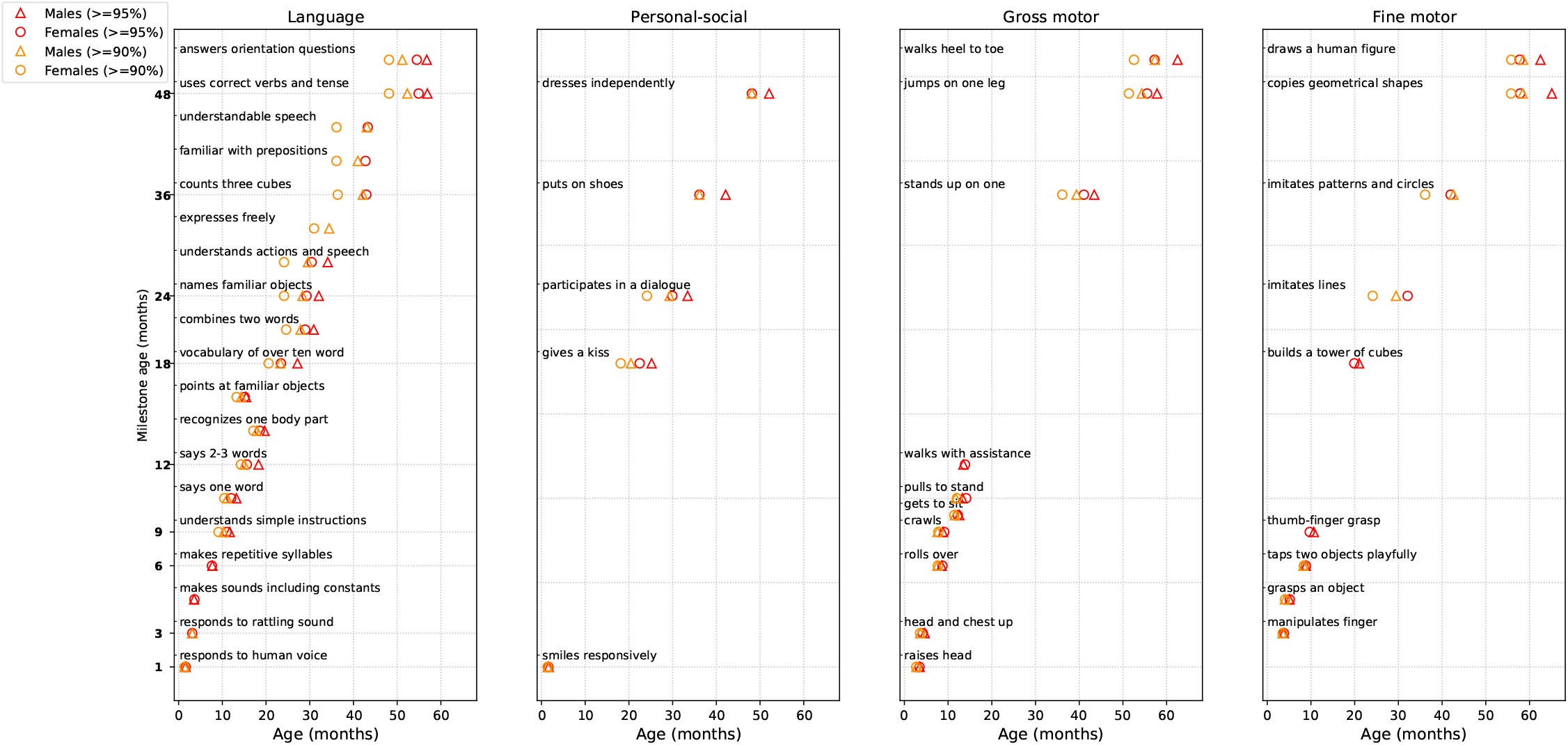
(a) – A comparison of milestones attainment age between sexes, grouped by developmental domain. A comparison of milestones attainment age between sexes, grouped by developmental domain is presented. The thresholds for the 90% and 95% achievement age of each milestone are demonstrated, for both sexes. The data analysis is based on the main cohort, which includes 3,774,517 developmental assessment visits of 643,958 children (319,562 girls, 49.6%).

**Figure2 (b).**
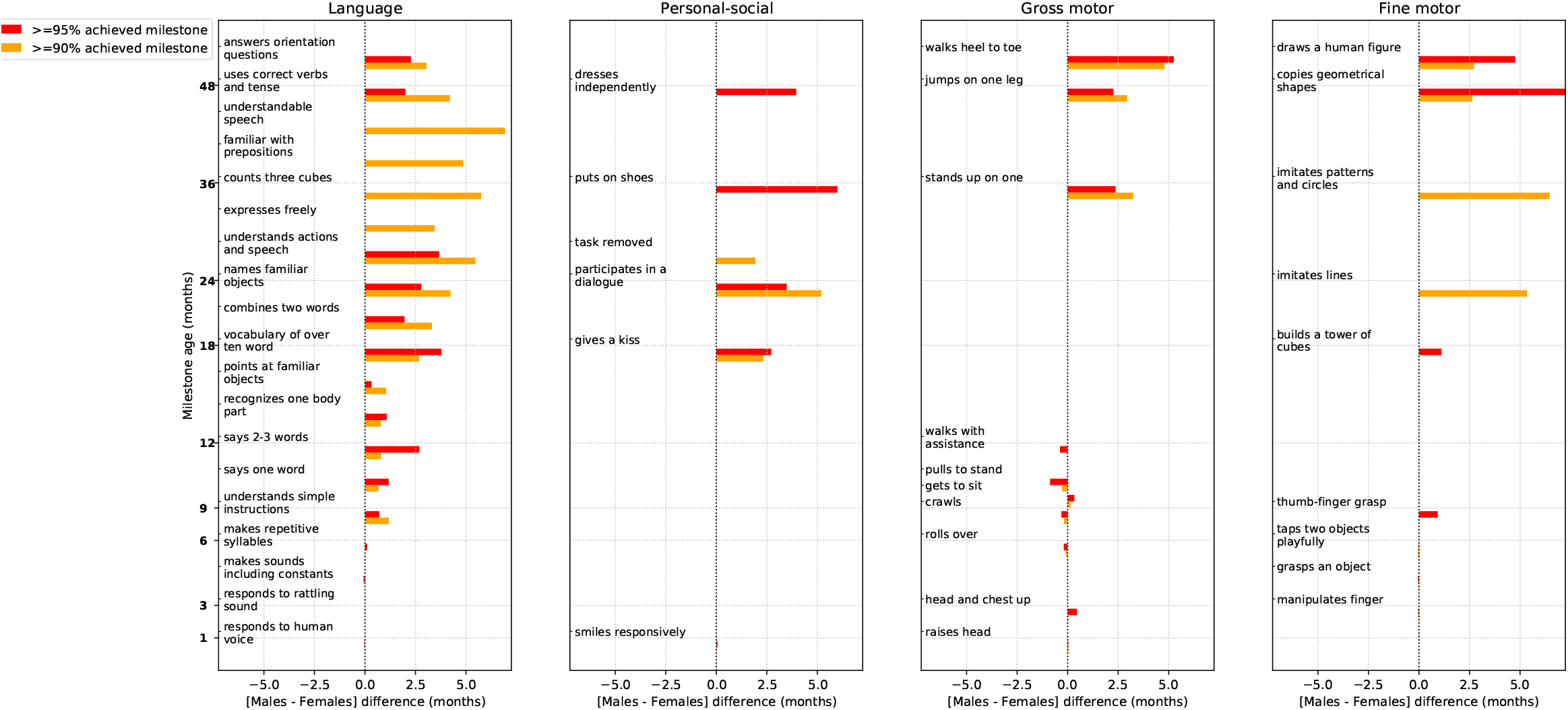
The time difference of milestones attainment age between sexes, grouped by developmental domain. The time difference of milestones attainment age between sexes is presented, grouped by developmental domain. The difference is calculated as a subtraction of the girl’s achievement age from the boy’s achievement age, at the 90% and 95% thresholds (, ). The data analysis is based on the main cohort, which includes 3,774,517 developmental assessment visits of 643,958 children (319,562 girls, 49.6%).

In total, 59 developmental milestones were evaluated from birth to the age of six years. In 17 milestones, the 95% success rate was already achieved by the two groups at the earliest evaluation age, rendering the threshold attainment age uncertain. In 6 additional milestones, the 95% success rate was not yet achieved by either group at the latest evaluation age, allowing only a comparison of the age thresholds of the 90% success rate. The comparison of the remaining 36 milestones (supplementary eTable 2) showed that in 24 milestones (66%) girls preceded boys, with a statistically significant difference in milestone attainment rate at age 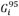 (denoted 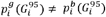, an earlier attainment age by 95% of the group 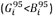, and an average attainment age gap 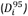 of 2.6±1.9 months (median 2.3, IQR [1.1,3.7] months). Boys slightly preceded girls (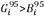 and 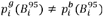) in 6 milestones (16%) with an average and 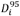 of 0.3±0.3 months (median 0.13, IQR [0.05,0.33] months), and in 6 milestones the difference between 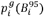 and 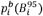 was not significant (*P*≥0.01). Among 35 milestones with comparable thresholds of 90% success rate, girls preceded boys (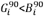 and 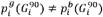, *P*<0.01) in 28 milestones (80%), with an average age gap 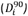 of 3.0±2.1 months (median 3.0, IQR [1.0,4.8] months). Boys preceded girls (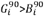 and 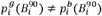, *P*<0.01) in 6 milestones (17%) with a small average 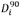 of 0.1±0.1 months (median 0.07, IQR [0.02,0.16] months). For most milestones, the attainment rates at the earliest evaluation age were higher than 75%. Nevertheless, a comparison of 9 milestones with available ages for 75% attainment rates demonstrated a similar trend - girls preceded boys in 7/9 milestones (78%), with an average age gap 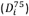 of 2.6±2.3 months, while boys marginally preceded girls by 0.2 months in a single milestone (“pulls to stand”), and there was no difference in the one remaining milestone.

Within the four developmental domains, profound differences were observed in the language milestones: girls preceded boys in achieving a 95% success rate in 10/14 milestones (71%, average 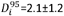 months), and in 15/16 milestones (94%) girls preceded boys in achieving the 90% success rate (average 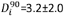 months). Girls also preceded boys in all 5 personal-social milestones (average 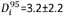 months), in 4/7 fine-motor milestones (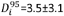 months), and in 5/10 gross-motor milestones (average 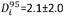 months). In the remaining tasks, there was either no statistical difference or boys preceded girls by negligible age differences. The earlier achievements of milestones by girls were more emphasized at older ages: while in the first year of life, there was no clear advantage to either sex group (6/16 vs. 5/16 milestones preceded by girls and boys, respectively), all milestones evaluated at age >12 months were achieved at an earlier age by 95% of the girls (16/16, average 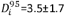 months), as well as by 90% of the girls (19/19, average 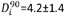 months).

supplementary eFigures 2-10 present the detailed curves of success rate for each milestone, for boys and girls.

### A comparison between THIS unified scale and sex-specific scales for developmental surveillance

Figure 3 presents the proposed separate sex-specific scales for the developmental surveillance of boys and girls. The rows indicate the evaluated milestones and the columns indicate the age in months, with green, yellow, orange, and red colors representing attainment age by less than 75%, 75-90%, 90-95%, and 95% of children, respectively.

**Figure 3.**
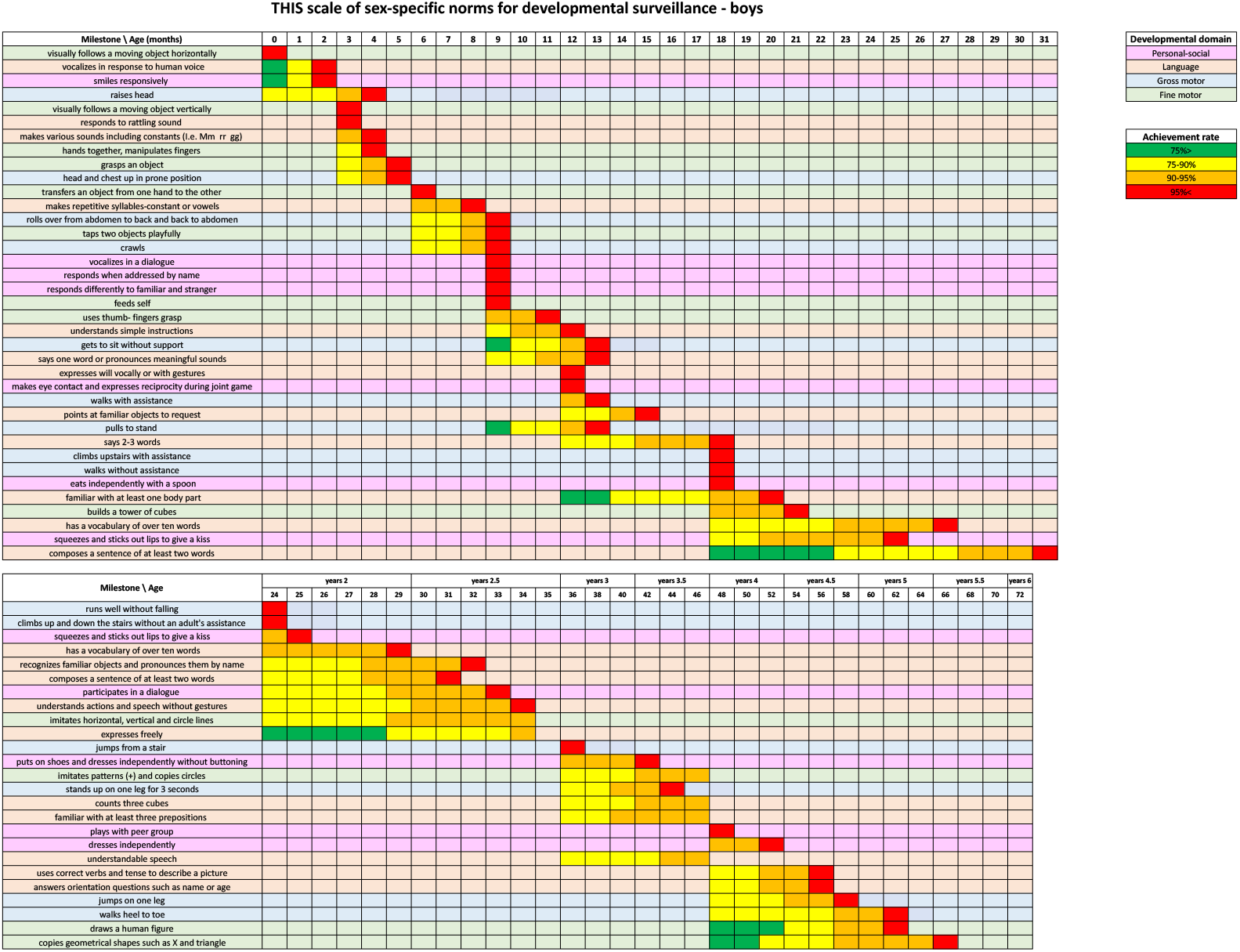
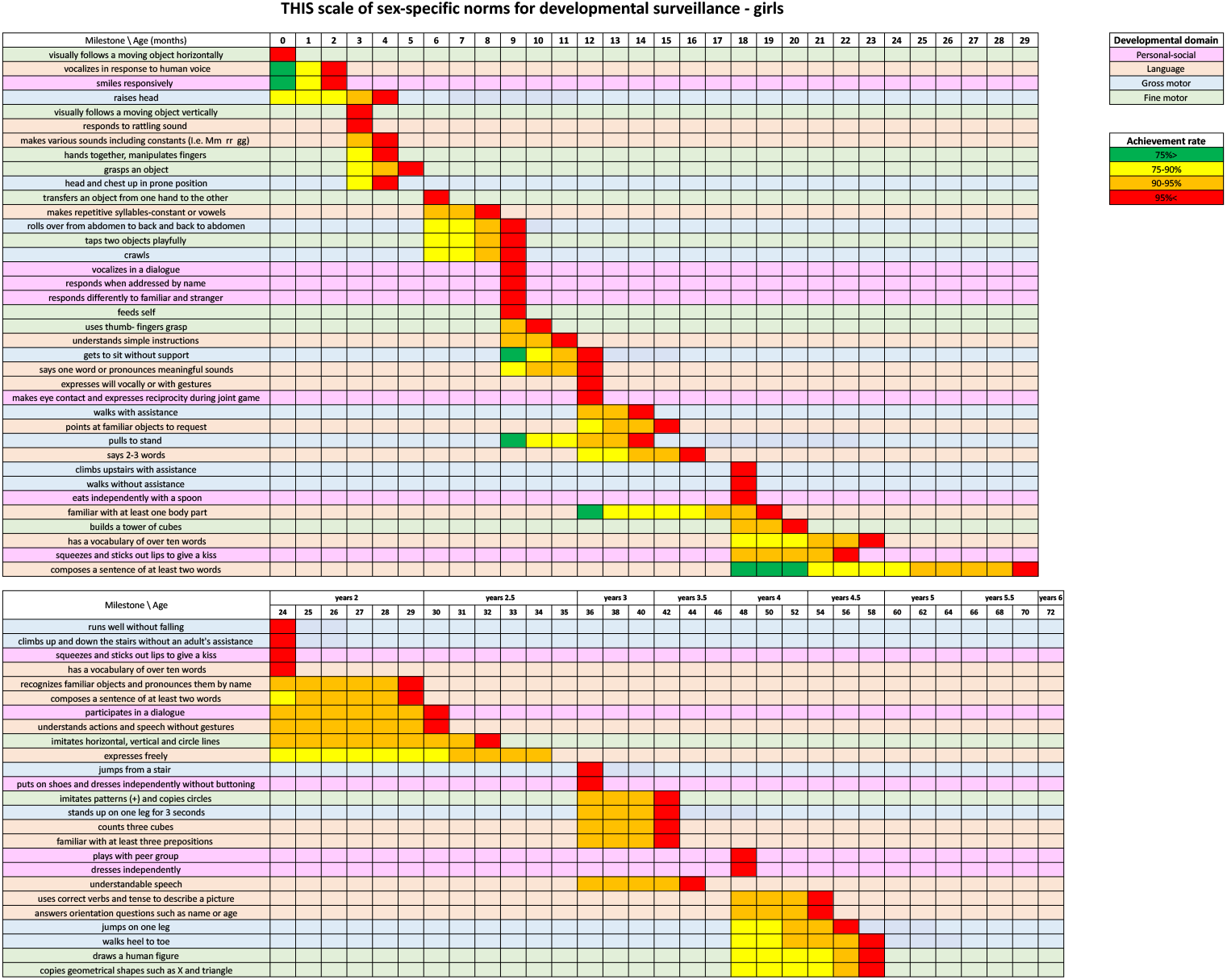
THIS scale of sex-specific norms for developmental surveillance. The achievement age of 59 milestones is presented in sex-specific scales. The columns indicate the age (by month) and the rows present the evaluated milestone. Each milestone is colored according to the relevant developmental domain (personal-social, language, fine and gross motor) and the attainment age of less than 75%, 75%-90%, 90%, and 95% is presented. For example - the milestone ‘smiles responsively’ is achieved by less than 75% of children aged less than 1 month, 75%-90% of children aged 1 month, and 95% of children aged 2 months. The presented norms are based on data analysis of the main cohort, which includes 3,774,517 developmental assessment visits of 643,958 children (319,562 girls, 49.6%).

The attainment age by 90% or 95% of the children according to the sex-specific scales was different from the reference attainment age by the unified THIS scale in 32/59 milestones for the boys (54%), and in 28/59 milestones for the girls (47%).

### Disagreement between unified and sex-specific scales

As shown in Table 2, in the main cohort, among visits with at least one failure in milestone attainment by girls (30.4% of all visits by girls), 18.7% of the visits that were labeled as ‘suspected delay’ (attainment age> 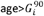) by the sex-specific scale would have been considered ‘no suspected delay’ by the THIS unified scale (false negative rate). On the other hand, among boys, visits with unattained millstones (34.4% of all visits by boys), 5.5% of the visits labeled ‘no suspected delay’ according to the sex-specific scale (attainment 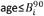) would have been considered ‘suspected delay’ by the unified THIS scale, thus triggering a false-alarm (false positive rate). These trends were evident in the social, language, and fine motor domains. Similar results were obtained by repeating the analysis on the validation cohort, with a false negative rate of 19.3% among girls and a false positive rate of 5.9% among boys.

**Table 2.** Disagreement between unified and sex-adjusted developmental scales.

| Milestone type | Total visits | Visits with unattained milestones | % visits with unattained milestones | Suspected delay by unified scale (≥90%) | No suspected delay by unified scale (<90%) | Suspected delay by sex-adjusted scale (≥90%) | No suspected delay by sex-adjusted scale (<90%) | FNR of unified scale | FPR of unified scale |
| --- | --- | --- | --- | --- | --- | --- | --- | --- | --- |
| **Boys – main cohort** |  |  |  |  |  |  |  |  |  |
| Any | 1880809 | 646182 | 34.4% | 235721 | 410461 | 211738 | 434444 | 0.0% | 5.5% |
| Personal - social |  | 114557 | 6.1% | 50622 | 63935 | 43211 | 71346 | 0.0% | 10.4% |
| Language |  | 387311 | 20.6% | 101738 | 285573 | 86518 | 300793 | 0.0% | 5.1% |
| Gross-motor |  | 224740 | 11.9% | 60256 | 164484 | 53960 | 170780 | 0.0% | 3.7% |
| Fine-motor |  | 168615 | 9.0% | 86477 | 82138 | 79959 | 88656 | 0.0% | 7.4% |
| **Girls – main cohort** |  |  |  |  |  |  |  |  |  |
| Any | 1844562 | 560483 | 30.4% | 196129 | 364354 | 241196 | 319287 | 18.7% | 0.0% |
| Personal - social |  | 80164 | 4.3% | 33199 | 46965 | 55840 | 24324 | 40.5% | 0.0% |
| Language |  | 302993 | 16.4% | 77375 | 225618 | 114176 | 188817 | 32.2% | 0.0% |
| Gross-motor |  | 224087 | 12.1% | 58572 | 165515 | 58679 | 165408 | 0.2% | 0.0% |
| Fine-motor |  | 143395 | 7.8% | 69900 | 73495 | 83725 | 59670 | 16.5% | 0.0% |
| **Boys – validation cohort** |  |  |  |  |  |  |  |  |  |
| Any | 411331 | 161161 | 39.2% | 65599 | 95562 | 59644 | 101517 | 0.0% | 5.9% |
| Personal - social |  | 35405 | 8.6% | 17047 | 18358 | 15038 | 20367 | 0.0% | 9.9% |
| Language |  | 103051 | 25.1% | 31223 | 71828 | 27220 | 75831 | 0.0% | 5.3% |
| Gross-motor |  | 54587 | 13.3% | 17414 | 37173 | 15757 | 38830 | 0.0% | 4.3% |
| Fine-motor |  | 47028 | 11.4% | 24198 | 22830 | 22463 | 24565 | 0.0% | 7.1% |
| **Girls – validation cohort** |  |  |  |  |  |  |  |  |  |
| Any | 385449 | 131046 | 34.0% | 50042 | 81004 | 62045 | 69001 | 19.3% | 0.0% |
| Personal - social |  | 23546 | 6.1% | 10638 | 12908 | 17342 | 6204 | 38.7% | 0.0% |
| Language |  | 77065 | 20.0% | 22134 | 54931 | 31921 | 45144 | 30.7% | 0.0% |
| Gross-motor |  | 49957 | 13.0% | 15132 | 34825 | 15262 | 34695 | 0.9% | 0.0% |
| Fine-motor |  | 36116 | 9.4% | 17978 | 18138 | 21817 | 14299 | 17.6% | 0.0% |

Visits with unattained milestones were labeled as ‘suspected delay’ or ‘no suspected delay’ according to the age norms of unified scale and sex-adjusted scales. The rate of false-negative and false-positive errors (FNR, FPR) by the unified scale was calculated, using the sex-adjusted scale as ground truth. The main cohort included all visits between January 2014 and September 2020, and the validation cohort included all visits from October 2020 to October 2021.

### Association between sex and failure in milestone attainment

We conducted regression analysis in order to assess the association between sex and failure to attain developmental milestones. The unadjusted and adjusted odds ratios for failing to attain each of the milestones by boys, compared to girls, are shown in supplementary eFigure 11, along with 95% confidence intervals. In 45/59 (76%) of the milestones, boys had significantly higher odds for failing (OR and 95% CI > 1), with adjusted OR ranging from 1.03 to 2.63 (median 1.50, IQR [1.26,1.70]). In 11 milestones (19%), boys had significantly lower odds for failing (OR and 95% CI < 1), with OR ranging from 0.81 to 0.98 (median 0.93, IQR [0.89,0.95]). In the remaining 3 milestones there were no significant differences between sex groups (CI overlapping 1). The higher odds of failure by boys were more accentuated at an older age (all 29 milestones evaluated at age>18 months) and in language and personal-social domains (27/31 milestones). Evidently, the changes in the odds ratios following adjustments for key socio-demographic variables were negligible.

## Discussion

Developmental scales, comprising norms of milestone attainment ages, are the cornerstone of routine developmental surveillance conducted by clinicians and parents globally^1^. These scales assist to identify children’s weaknesses and strengths and to recognize those who may need further follow-up. Currently, developmental surveillance is performed worldwide using unified scales, which may introduce a potential bias.

The current study evaluated the differences in milestone attainment rate between boys and girls during early childhood years, using two large cohorts consisting of more than 4.3 million developmental assessments. Analysis of these developmental assessments indicated that girls precede boys in attaining most milestones of all developmental domains. These differences were more apparent at older ages and were most significant in the personal-social domain, followed by language skills, fine motor, and, to a lesser degree, gross motor.

Unlike developmental screening, which utilizes tools with known predicted performance that were validated against a future outcome, developmental surveillance is based solely on norms of developmental milestones attainment age. Hence, the utility of developmental surveillance cannot be assessed by evaluating long-term outcomes or psychometric properties. To assess the potential benefit of sex-specific developmental surveillance we have calculated the projected error rates of a unified developmental scale when the sex-specific scales are considered the ground truth. Our results show that when a single scale is used for both sexes, there is a substantial subgroup of girls whose failure to achieve a milestone may be considered insignificant (age below the populations’ 90% threshold age), although it is interpreted as a suspected delay (age≥90%) by the sex-specific scale (18.7% and 19.3% of failed assessments in the main and validation cohorts, respectively). This underestimation may result in reduced identification of girls in need of further follow-up, preventing their timely evaluation. For boys, the unified scale gives false positive alerts for cases where the age of milestone failure is below the 90% threshold of the sex-specific scale (5.5% and 5.9% of failed assessments in the main and validation cohorts, respectively). Overestimating the significance of unattained milestones by boys may result in unnecessary parental and clinical concern.

A possible explanation for the observed difference in milestones attainment rate between sexes is the higher prevalence of conditions related to developmental delay among boys. To reduce this potential bias we initially excluded children with abnormal developmental potential from both groups (preterms, failure to thrive, microcephaly, and macrocephaly). Moreover, we conducted an additional sensitivity analysis using a subset of ‘successful’ children, who have attained all developmental milestones at age 24-36 months. Comparison of the differences between sexes in attainment age of earlier milestones demonstrated that girls precede boys in all language and personal-social milestones at ages 9-24 months (supplementary eFigure 12). This further supports our assumption that the reported results represent an actual difference in the rate of milestone attainment between normally developed children and they do not stem from a higher prevalence of developmental conditions in the boys’ group.

Consistent with our findings, previous studies have demonstrated sex-related differences in early childhood development, mainly in social, language, and fine motor skills^20–30^. Although some of these differences were evident in major studies that were fundamental in the establishment of milestone norms^9,10^, as well as in recent large cohort studies^16^, they were not referred to as clinically significant.

This study includes the largest multicultural cohort of developmental assessments analyzed on a national scale, rather than a smaller sample of the population. Furthermore, the reliability of the developmental assessments transcends those of previous studies since they are based on evaluations conducted by trained public health nurses, rather than parental reports^23,24,27,28^.

To the best of our knowledge, this study is the first to evaluate the clinical significance of separate sex-specific scales by demonstrating the potential rates of false-negative and false-positive errors when conducting unified developmental surveillance. This differs from earlier studies in which clinical significance was determined arbitrarily and inconsistently^9,10,16^.

Investigating the underlying causes of sex-related differences remains a challenge to neuroscientists worldwide, due to the difficulty in disentangling biological sex differences from possible environmental influence^15,18,22^. There are known congenital and physiological differences between girls and boys. Extensive research has established the presence of biological sex-related differences in neurodevelopment, demonstrated by variations in brain structure and function, such as total brain volume, cortical volume, gray matter density, and white matter organization. It has been observed that girls reach their peak brain volume, as well as their puberty, earlier than boys^15^. These findings are in agreement with the hypothesis that girls normally mature earlier than boys, even during the first years of life. Therefore, the observed differences in milestones attainment age merely represent different maturation rates, while the endpoint abilities are the same. Eventually, children of both sexes accomplish all milestones by the age of six years and there is no evident developmental gap. This phenomenon may be equivalent to the known physiological difference during puberty, at which girls mature earlier than boys. Caregivers are well aware of this difference and therefore do not expect the same timeline for sexual maturity of teenagers. Our observations suggest that a similar logic applies during early childhood development.

Although there are possible environmental and social factors contributing to sex-related differences in developmental milestones attainment, our results indicate that the observed differences may be primarily attributed to intrinsic biological differences for several reasons. First, the differences are significant across all developmental domains, whereas the environmental component is expected to affect only part of the developmental domains. Second, the gap is evident from a very young age for basic milestones such as lifting the head and chest, in which an environmental effect is less likely. Third, the observed differences remain significant after controlling for socio-demographic variables that may be associated with developmental outcomes. Fourth, by the age of six years, both sexes accomplish all milestones, indicating that the sex-related differences reflect variations in the maturation rate, rather than different developmental endpoints.

Conducting routine developmental surveillance based on inaccurate milestone norms may lead to an extensive unnecessary burden. False-alarming unnecessary developmental work-up for normally developed children may cause parental emotional distress, as well as faulty allocation of limited financial and medical resources. On the other hand, missing children at risk of developmental delay may result in late intervention and suboptimal outcomes. Thus, performing routine sex-specific developmental surveillance may optimize the cost-effectiveness of the evaluation.

This study has several limitations. The study population is limited to Israel, and while it is a heterogeneous, multicultural population, the presented findings require further evaluation worldwide. The use of attainment rate cutoffs of 90% and 95% represents the common practice in the Israeli developmental surveillance program. Although the recently updated checklist of CDC milestones^1^ defined threshold ages with a cutoff attainment rate of 75%, in our data the actual attainment rates of most comparable milestones were considerably higher than 75%, indicating that these milestones were achieved by most of the Israeli children at an earlier age than suggested by the CDC checklists^37^. In addition, of milestones available in the literature, only 59 major milestones were evaluated in our scale, and some of these milestones (17/59) had already exceeded the 95% threshold at their initial evaluation, thereby precluding measuring the extent of their sex-related differences. As our observation that girls preceded boys in most milestones was consistent across all developmental domains, we believe that it represents a general finding. However, further evaluation of sex-related differences in additional milestones and at earlier age thresholds is required to further validate the results.

Finally, although the study establishes the difference in milestone achievement rate between sexes, and suggests sex-specific scales for developmental surveillance, it does not evaluate how these variations affect the accuracy of developmental screening and assessment tools commonly used. Therefore, further study is required to assess the potential of establishing sex-specific screening and diagnostic tools, using longitudinal data that include reliable developmental outcomes of the evaluated children.

## Conclusions

This study demonstrates significant sex-related differences in norms of milestone attainment as part of developmental surveillance. These differences are of fundamental clinical and social importance, indicating a possible bias in the currently used developmental scales. Our findings suggest the clinical need for utilizing sex-specific scales in early childhood developmental surveillance.

## Acknowledgments

We thank Dr. Chen Yanover and Dr. Maytal Bivas-Benita for their valuable comments to this manuscript.

The authors received no specific funding for this work.

## Author contribution

T.S, G.A, and Y.S conceptualized and designed the study, carried out the data analysis, drafted the initial manuscript, and prepared the revised manuscript. T.S, G.A, and Y.S directly accessed and verified the underlying data reported in the manuscript.

M.A.T, P.A, and E.B participated in conceptualizing and designing the study and critically reviewed the manuscript for important intellectual content.

D.R.Z, H.Y, R.B, and D.B.M designed the data collection instruments, collected data, facilitated access to the data and critically reviewed the manuscript.

The corresponding author attests that all listed authors meet authorship criteria and that no others meeting the criteria have been omitted

