## Supplementary data for "Sex-specific developmental scales improve early childhood developmental surveillance"

**Online Supplements**

1. eFigure 1 -Notation’s illustration of comparison of milestone attainment between sex groups
2. eFigure 2 - Task performance results over time according to sex at the age 0-3 months
3. eFigure 3 - Task performance results over time according to sex at the age 3-6 months
4. eFigure 4 - Task performance results over time according to sex at the age 6-9 months
5. eFigure 5 - Task performance results over time according to sex at the age 9-12 months
6. eFigure 6 - Task performance results over time according to sex at the age 12-18 months
7. eFigure 7 - Task performance results over time according to sex at the age 18-24 months
8. eFigure 8 - Task performance results over time according to sex at the age 2-3 years (24-36 months)
9. eFigure 9 - Task performance results over time according to sex at the age 3-4 years (36-48 months)
10. eFigure 10 - Task performance results over time according to sex at the age 4-6 years (48-72 months)
11. eFigure 11 – Odds ratio for unattained developmental milestones by sex
12. eFigure 12 – Comparison of milestones attainment rate between sexes of ‘successful’ children at the age of two years
13. eTable 1 - Israeli Ministry of Health - A guide to performing developmental assessments for infants and toddlers up to the age of six years
14. eTable 2 – A comparison of milestones attainment age according to sex
15. eTable 3 – Milestones names legend for Figure 2

**eFigure 1 – Notation’s illustration of comparison of milestone attainment between sex groups**

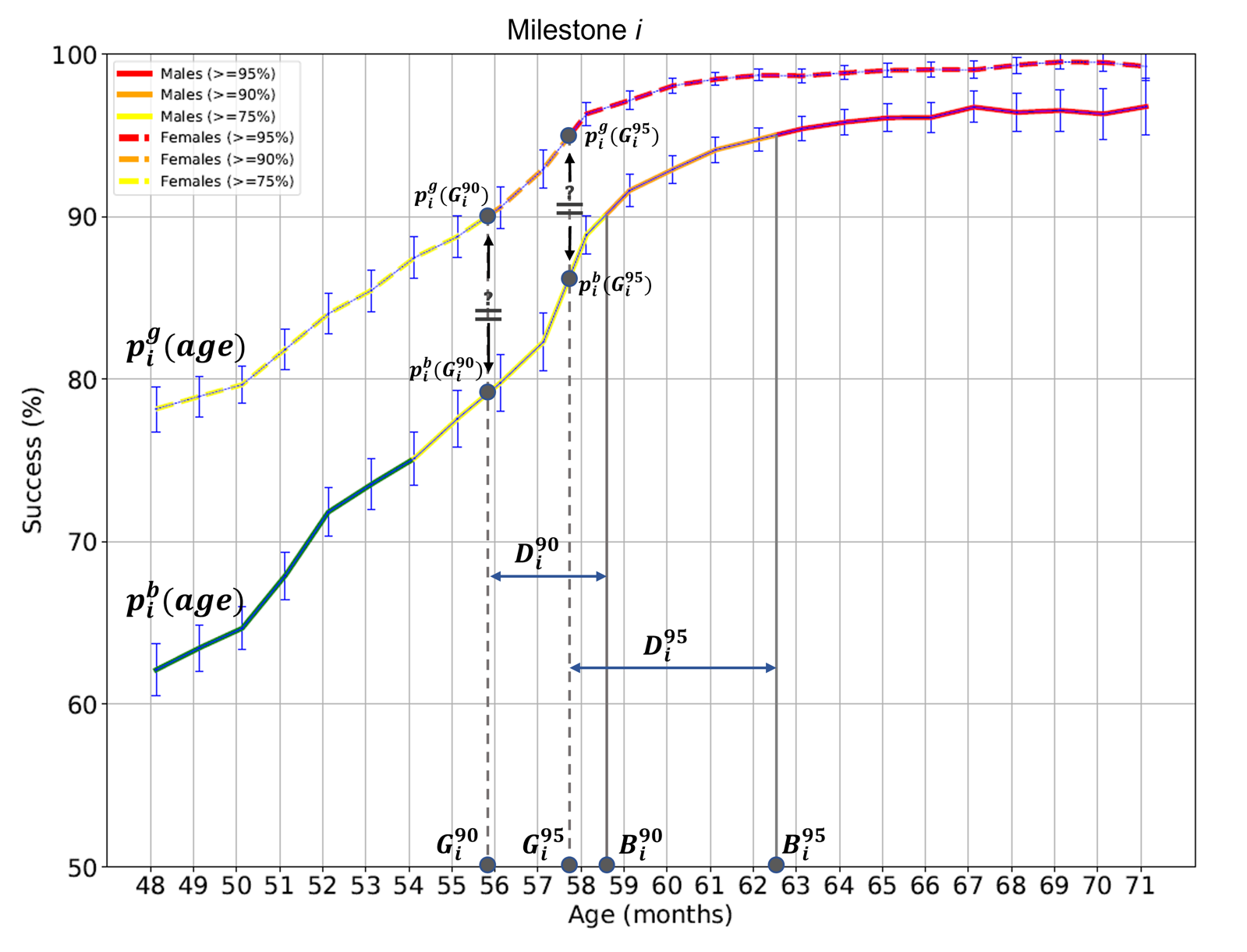

eFigure 1: Trends of the proportion attaining milestone *i* as a function of age by boys ($p_{i}^{b}(age)$) and girls ($p_{i}^{g}(age)$). $B_{i}^{90}$ *,* $B_{i}^{95}$ are the threshold age for attaining the milestone by 90% and 95% of the boys, respectively. Similarly, $G_{i}^{90}$ *,* $G_{i}^{95}$ are the threshold ages for attaining the milestone by 90% and 95% of the girls, respectively. $D_{i}^{90} , D_{i}^{95}$are the age gaps between boys and girls in attaining the milestone by 90% and 95% of the group, respectively. Assuming $G_{i}^{95}$*<*$B_{i}^{95}$*,* We assess the significance of the difference between the success rates of boys and girls by testing the null hypothesis that $p_{i}^{g}(G_{i}^{95})=$ $p_{i}^{b}(G_{i}^{95})$*.* Similarly, we test the hypothesis that $p_{i}^{g}(G_{i}^{90})=$ $p_{i}^{b}(G_{i}^{90})$*.*

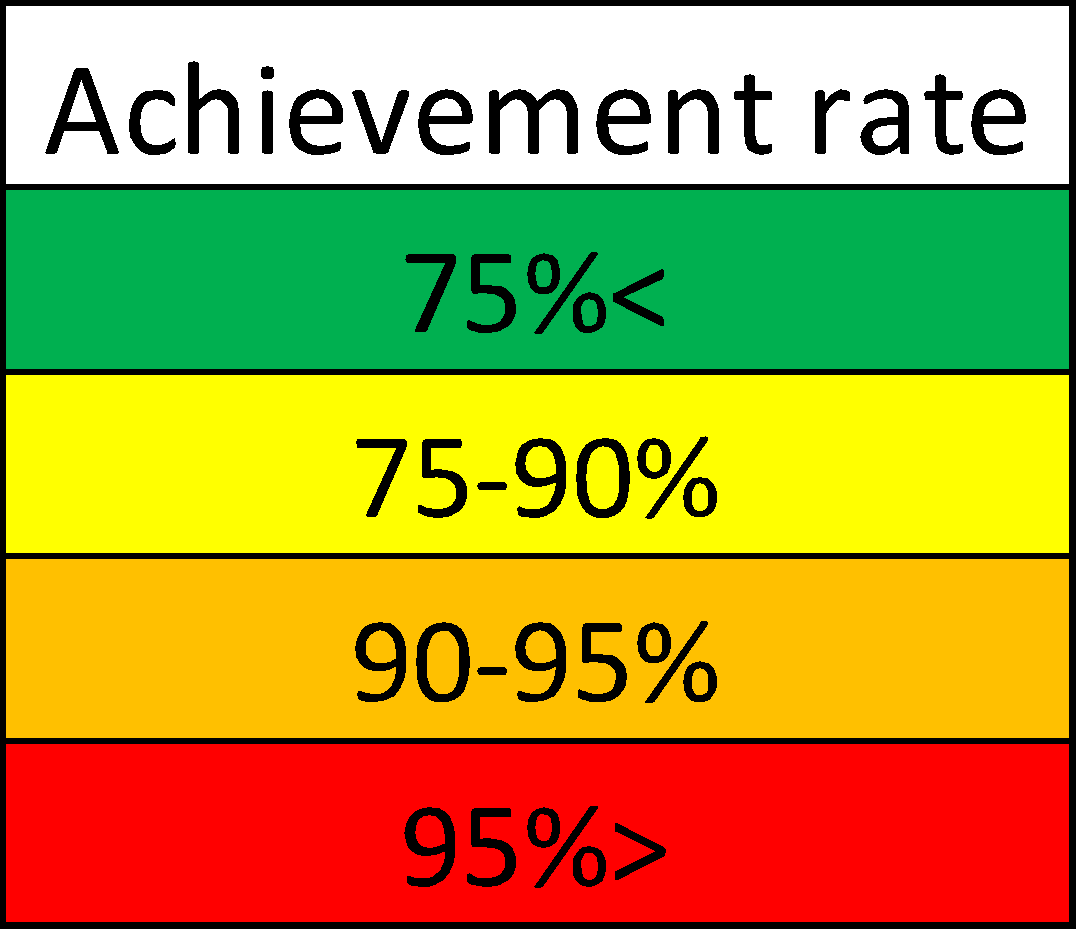
**eFigure 2 - Task performance results over time according to sex at the age 0-3 months**

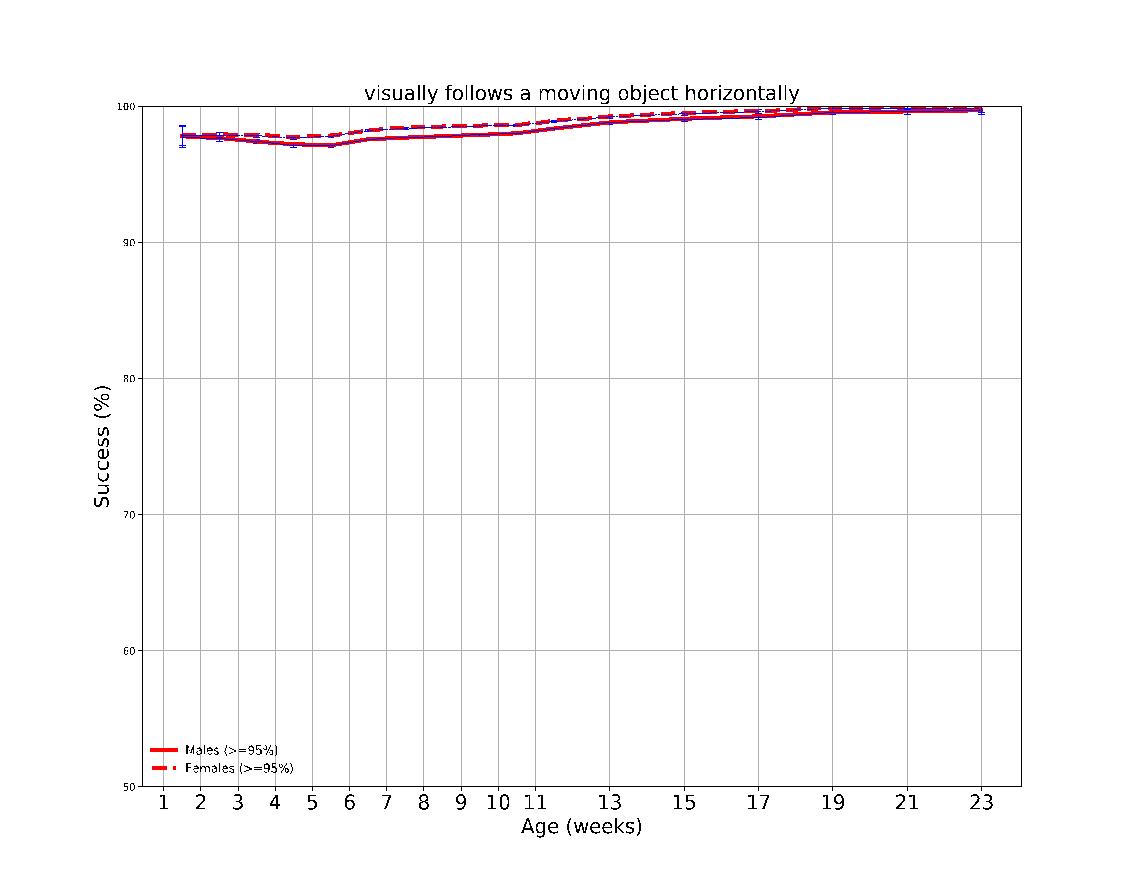

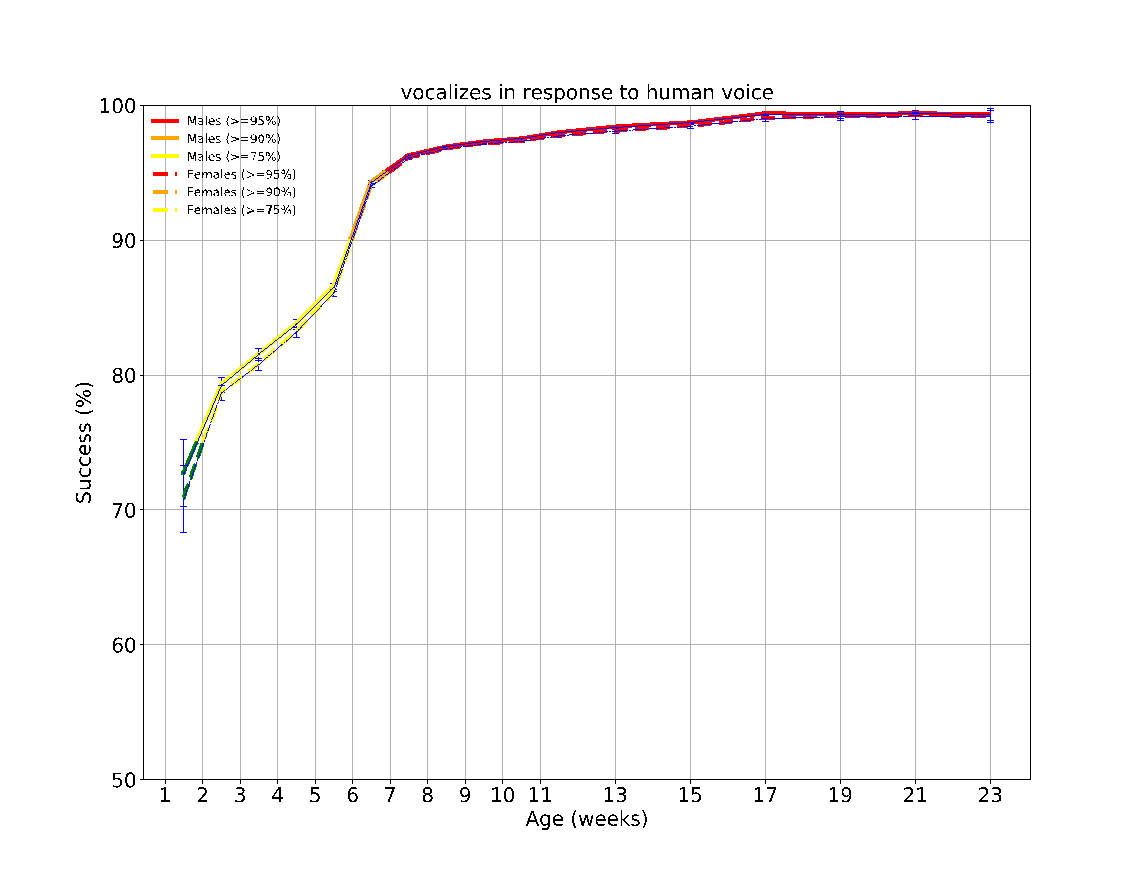

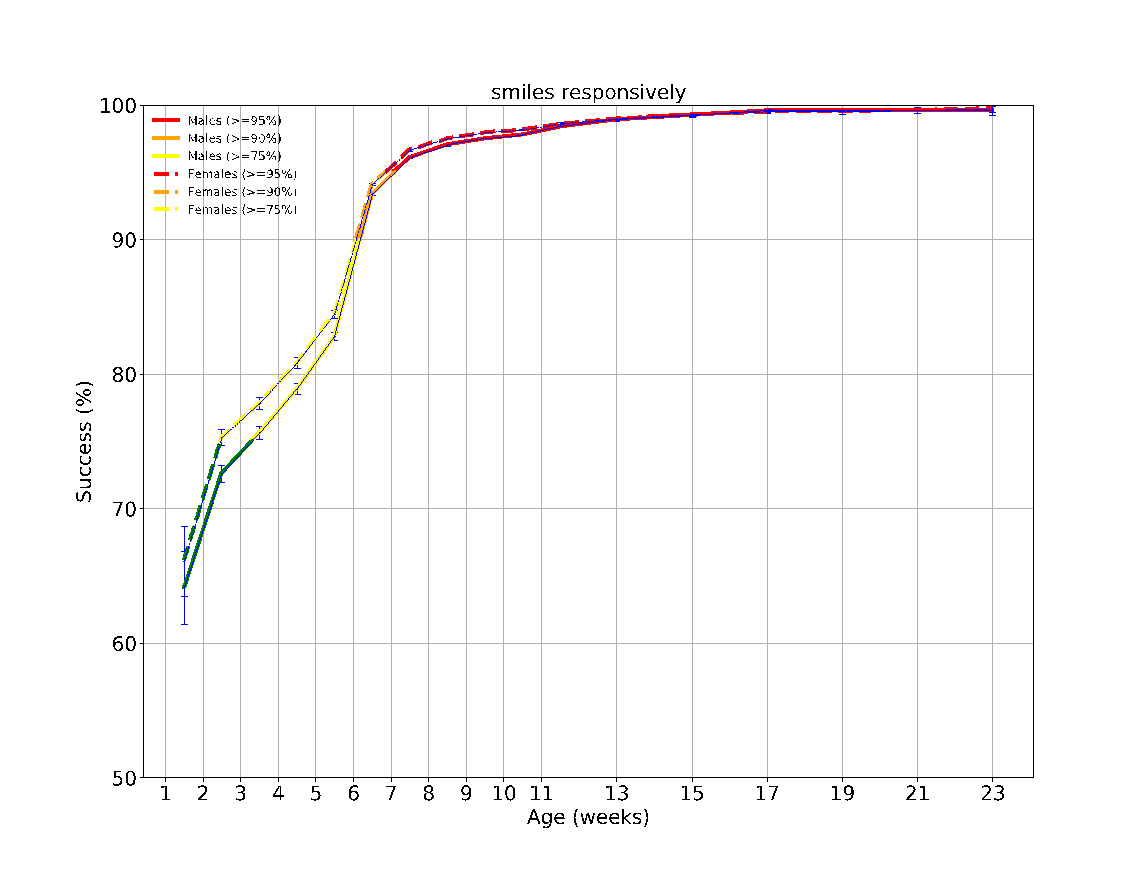

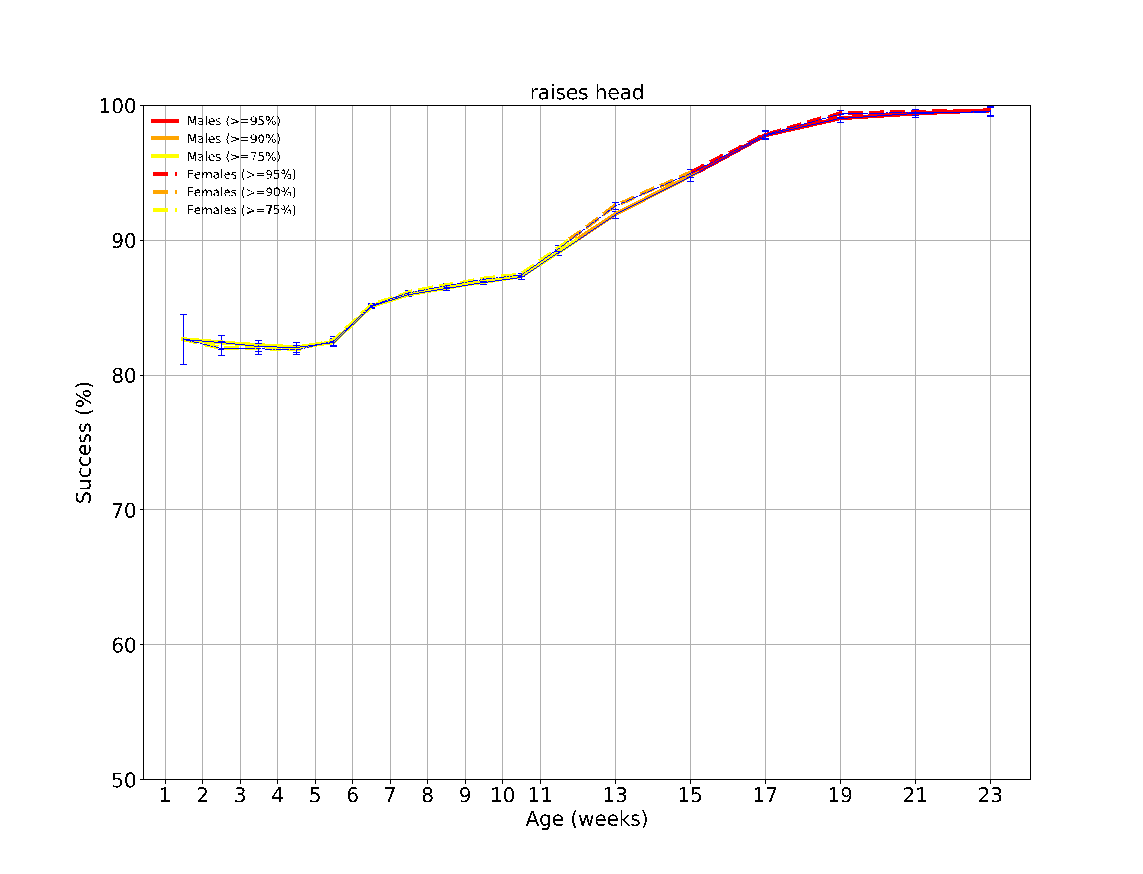

eFigure 2: The success rates over time of the different milestones evaluated between the ages 0-3 months. The achievement rates of < 75%, 75-90%, 90-95% and > 95% of children are indicated by the colors green, yellow, orange and red, respectively. The error bars are 95% confidence intervals of a binomial proportion, where larger bars indicate smaller sample size.

**eFigure 3 - Task performance results over time according to sex at the age 3-6 months**
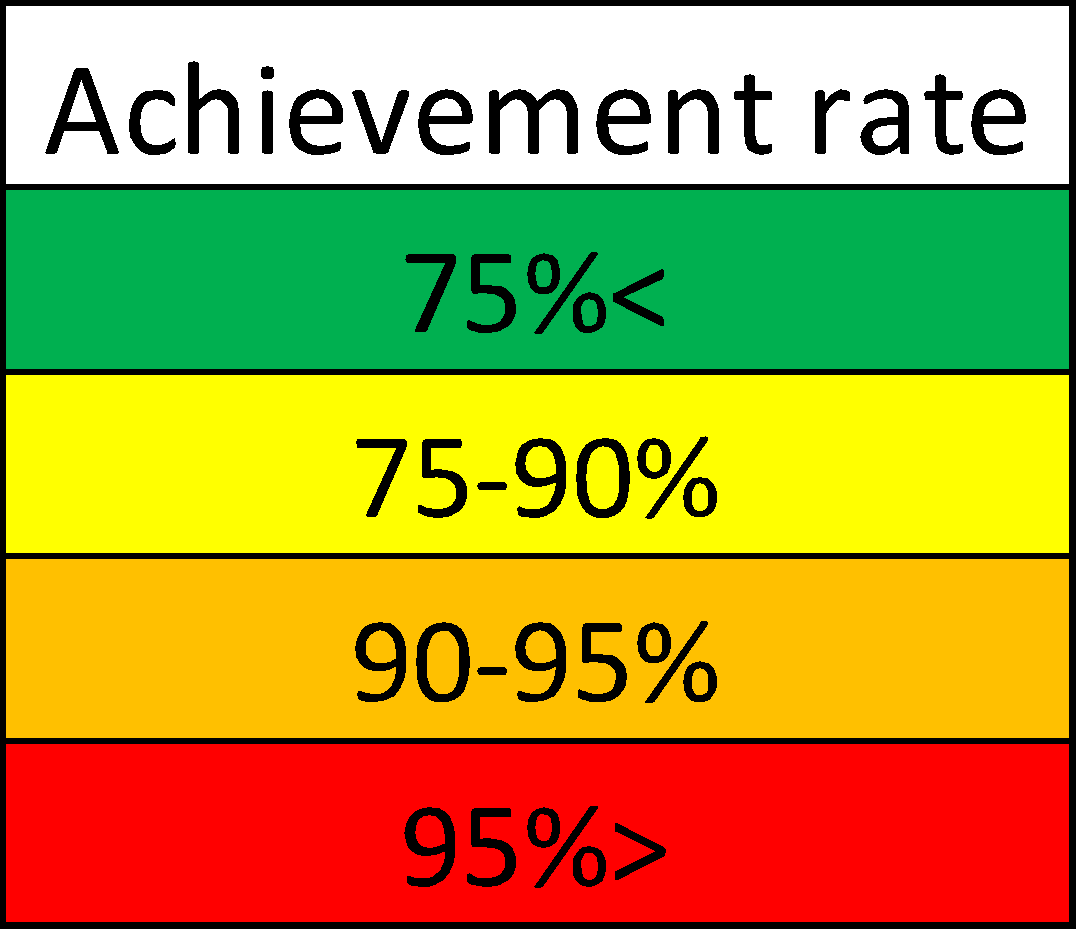

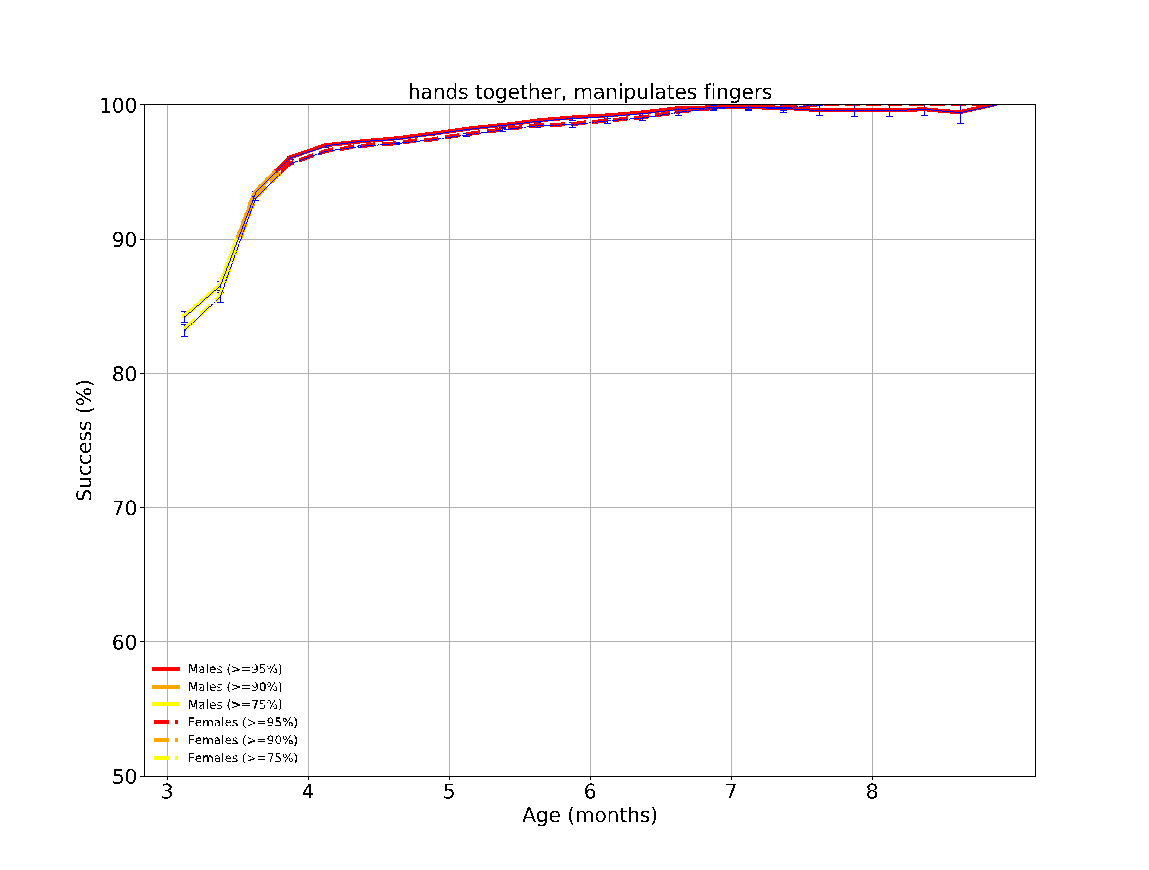

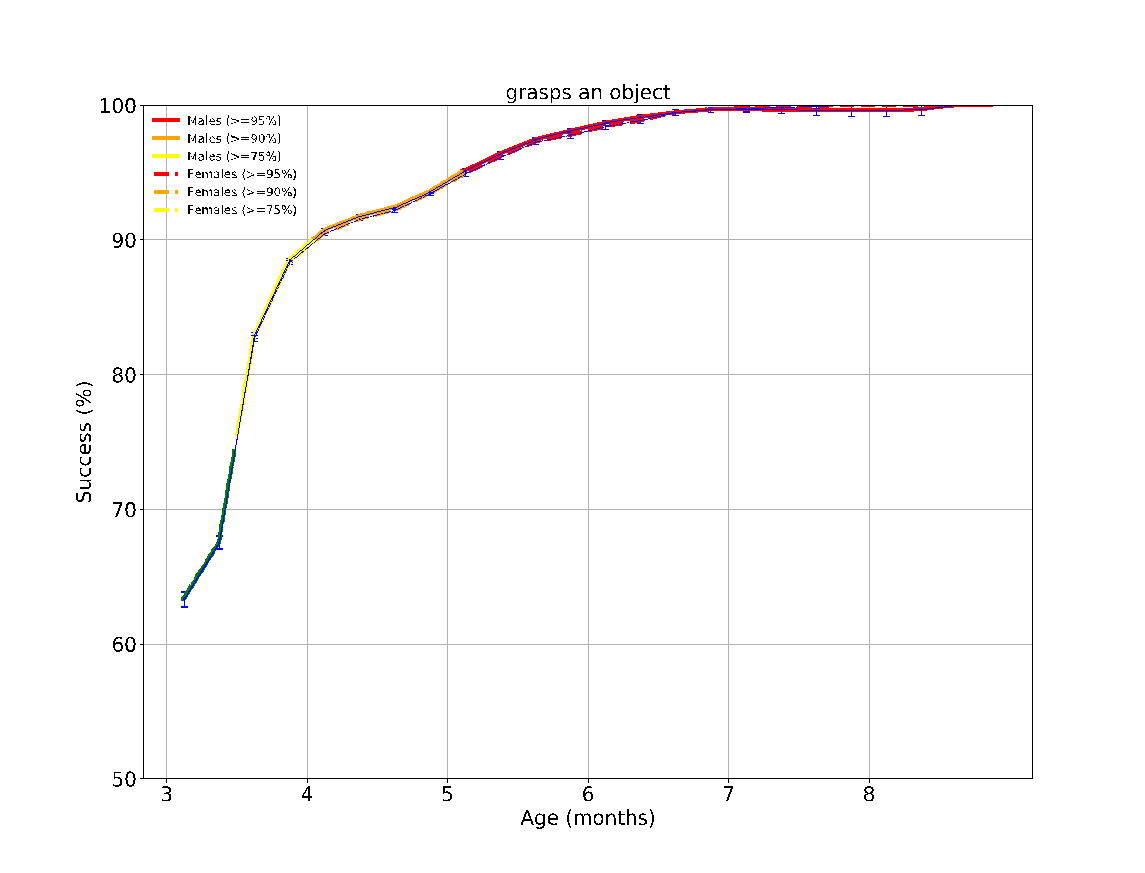

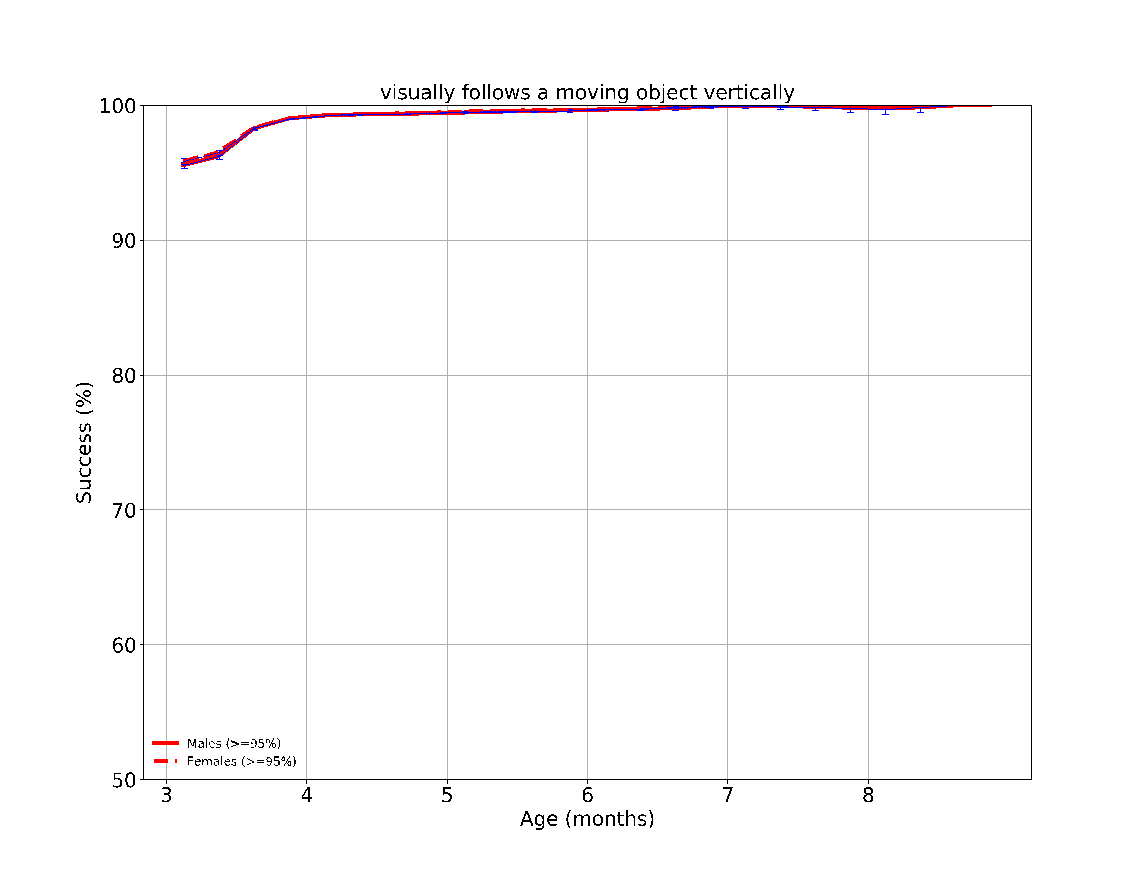

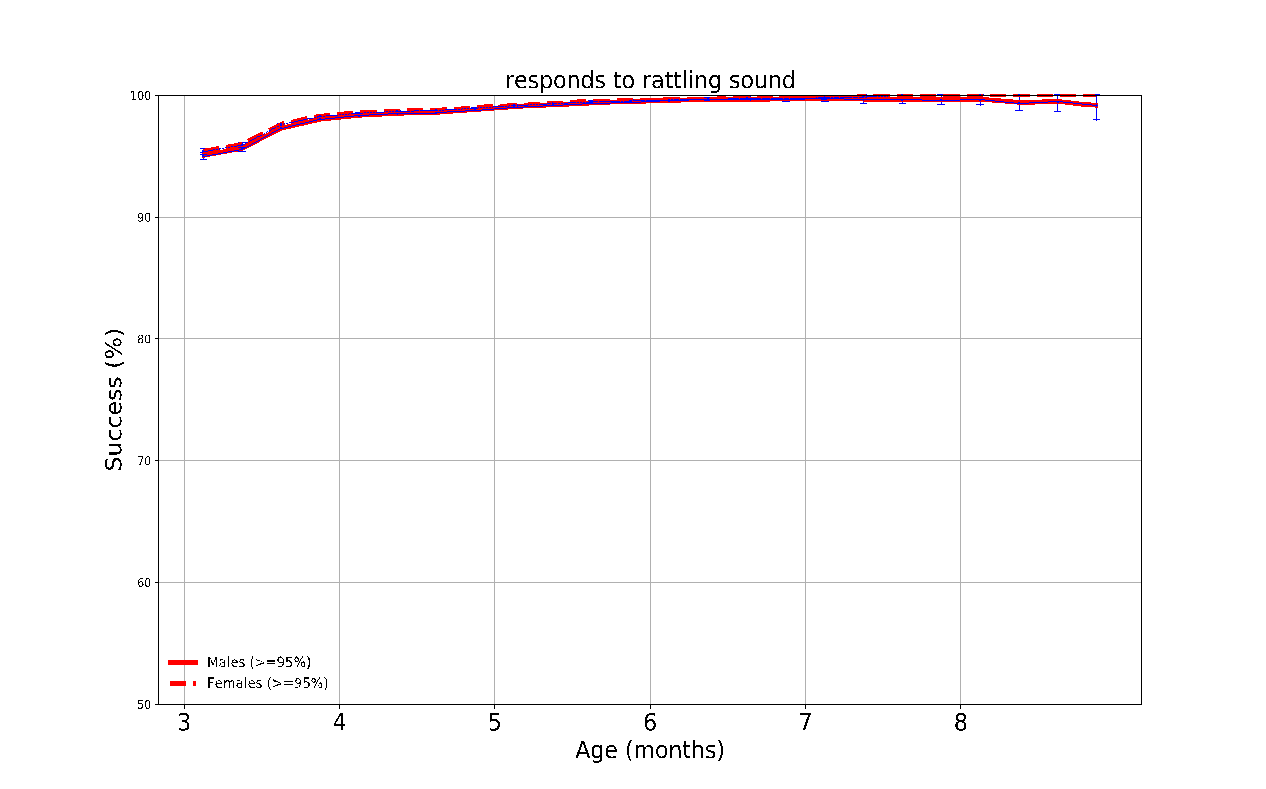

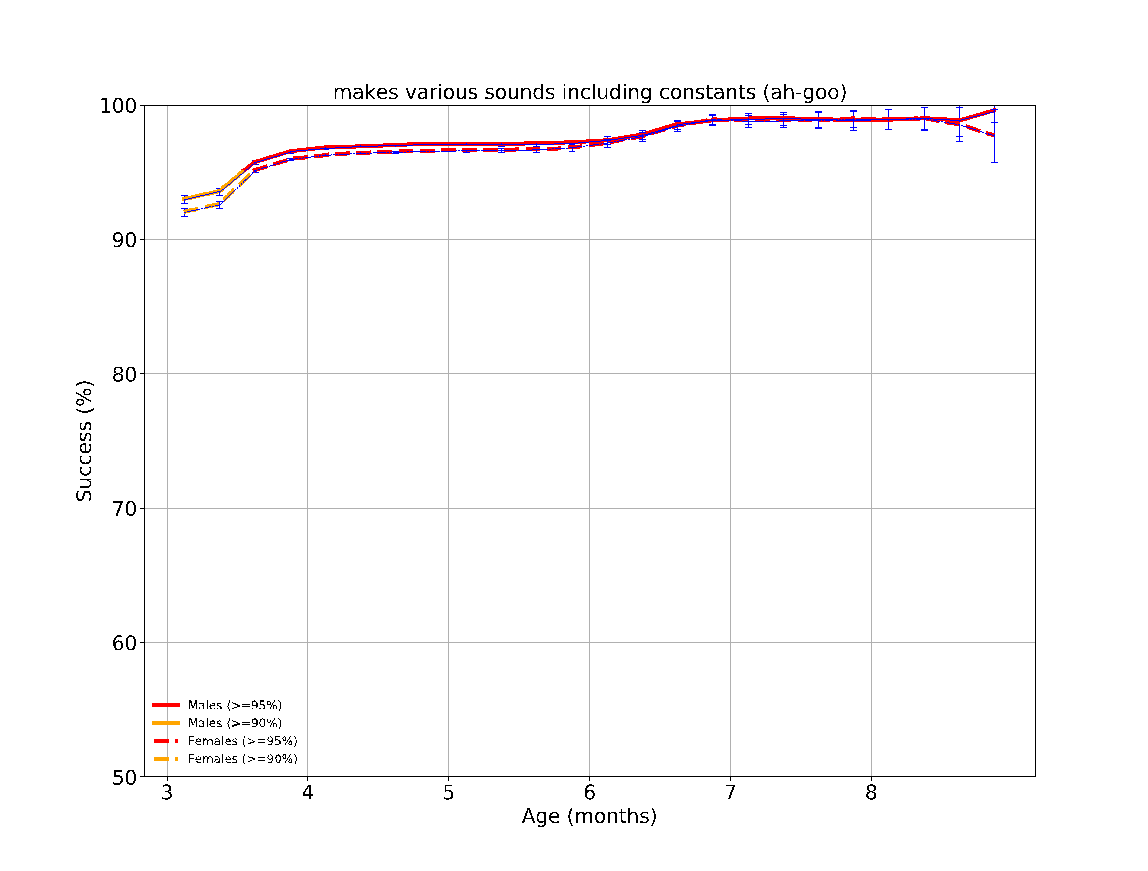

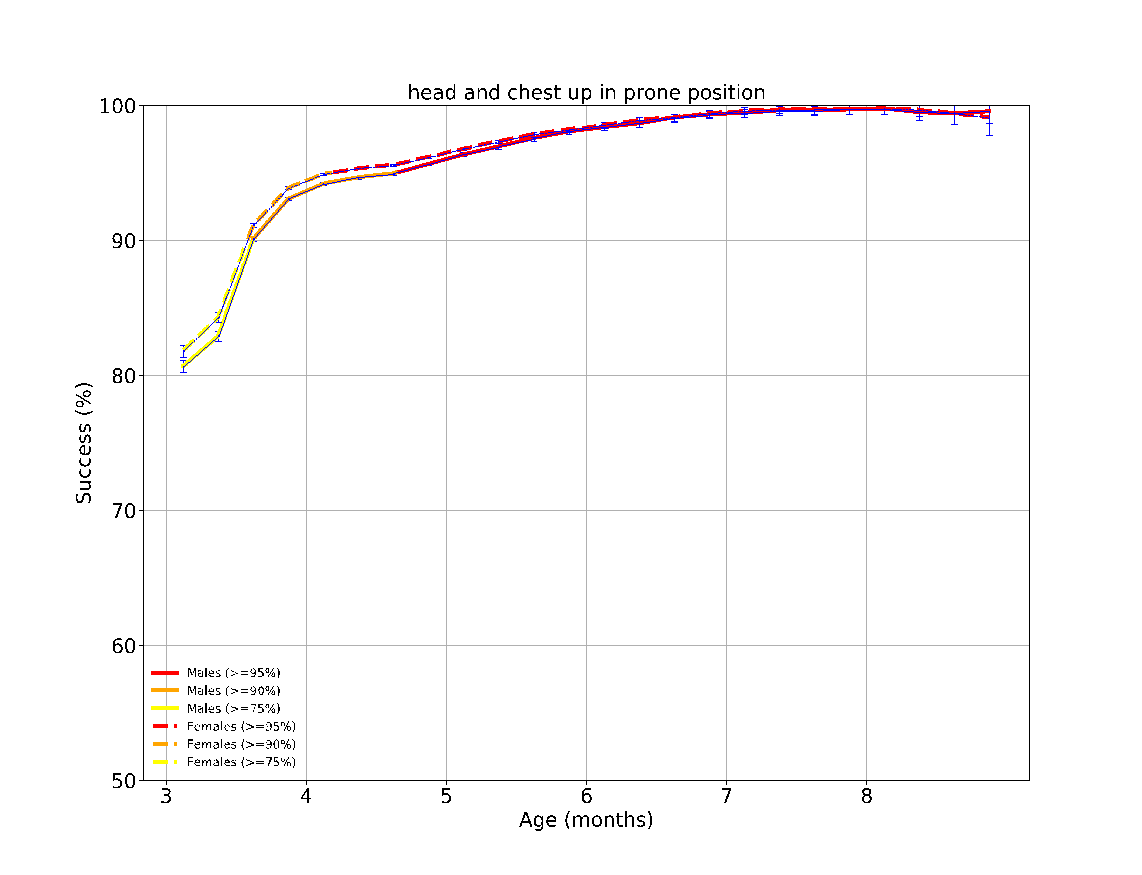

eFigure 3: The success rates over time of the different milestones evaluated between the ages 3-6 months. The achievement rates of < 75%, 75-90%, 90-95% and > 95% of children are indicated by the colors green, yellow, orange and red, respectively. The error bars are 95% confidence intervals of a binomial proportion, where larger bars indicate smaller sample size.

**eFigure 4 - Task performance results over time according to sex at the age 6-9 months**
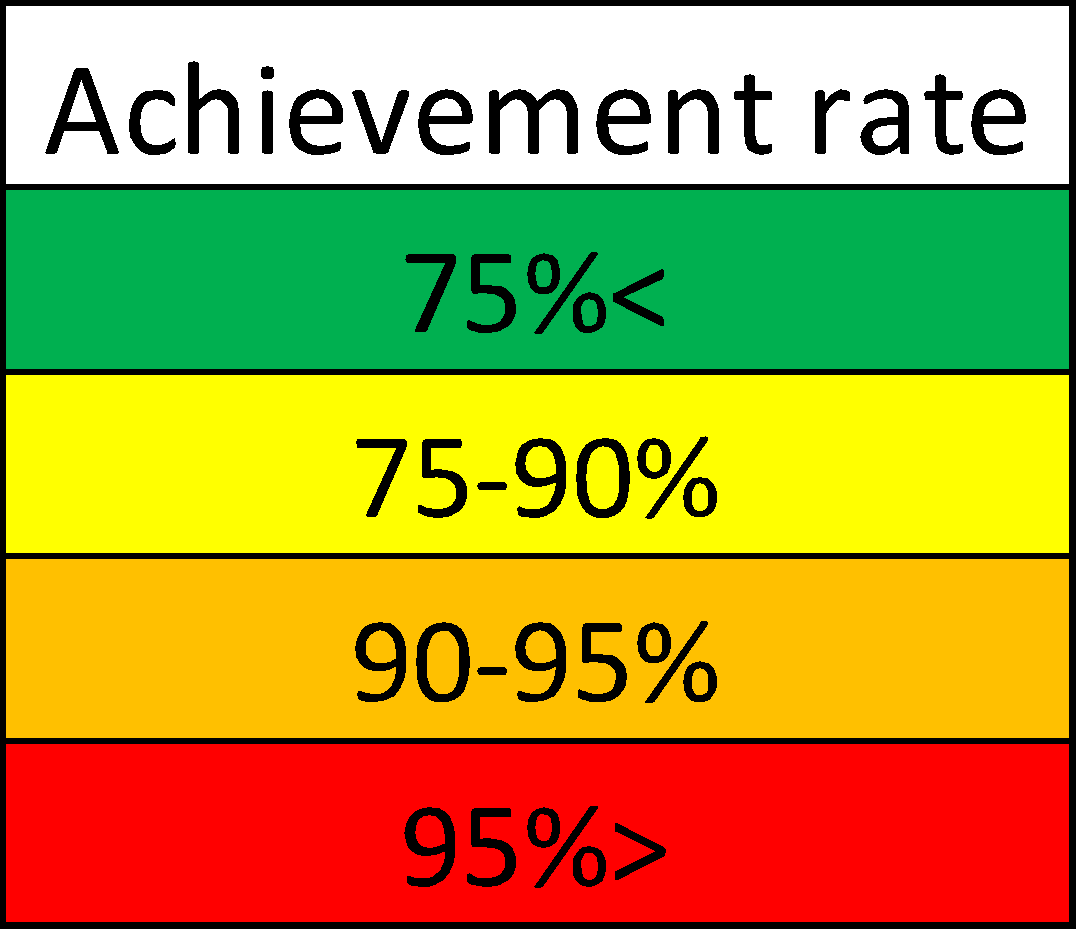

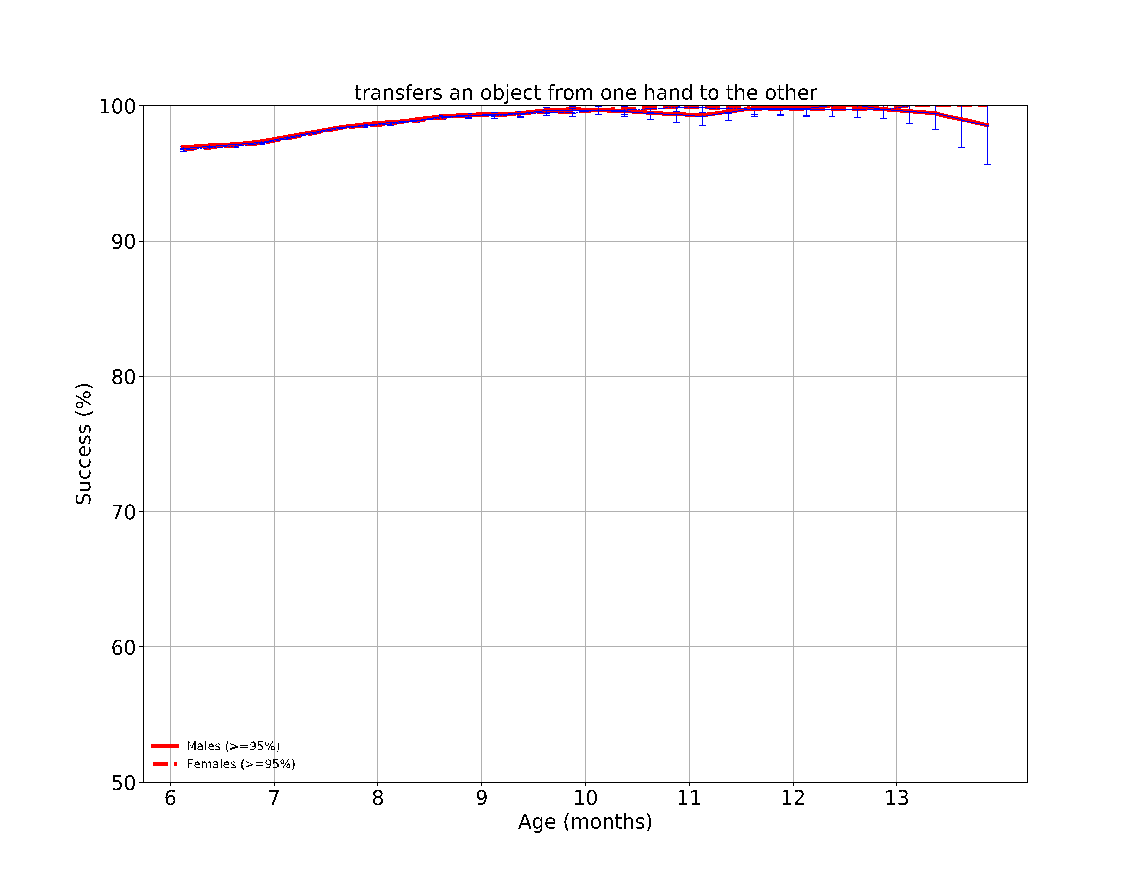

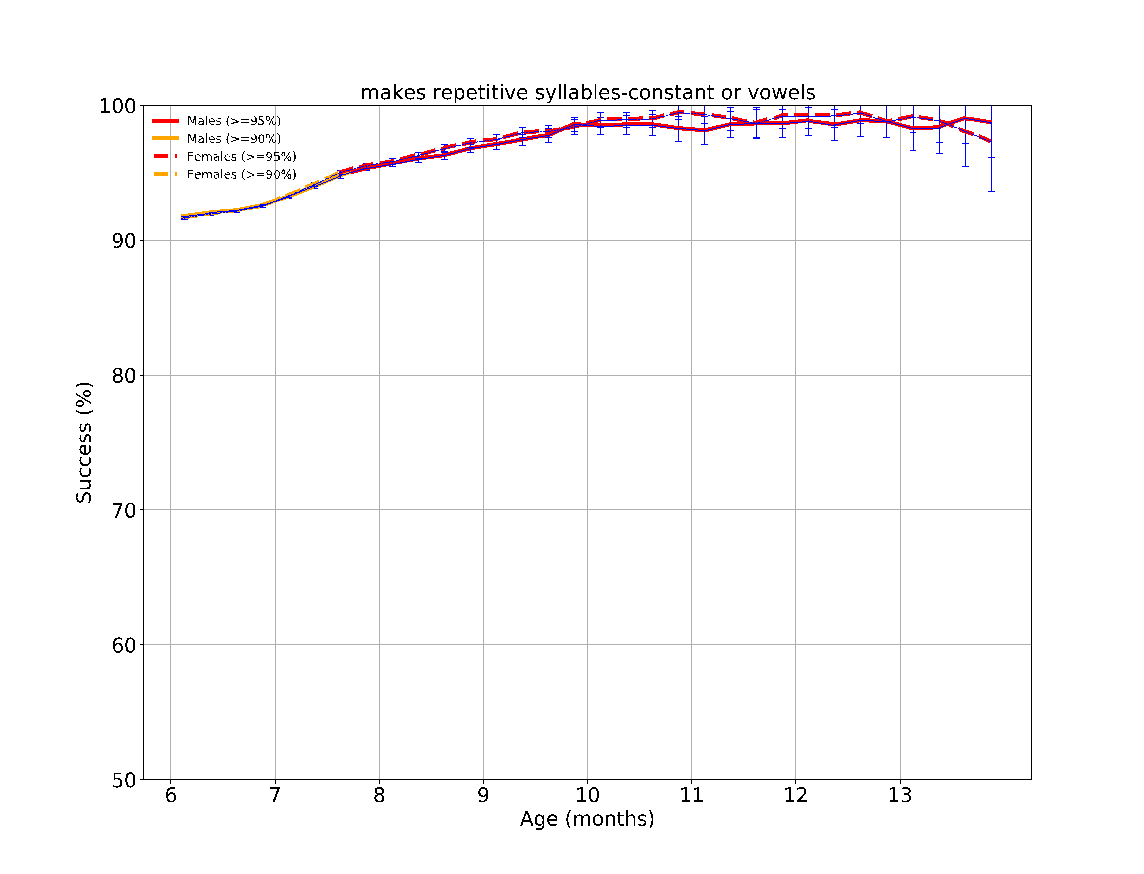

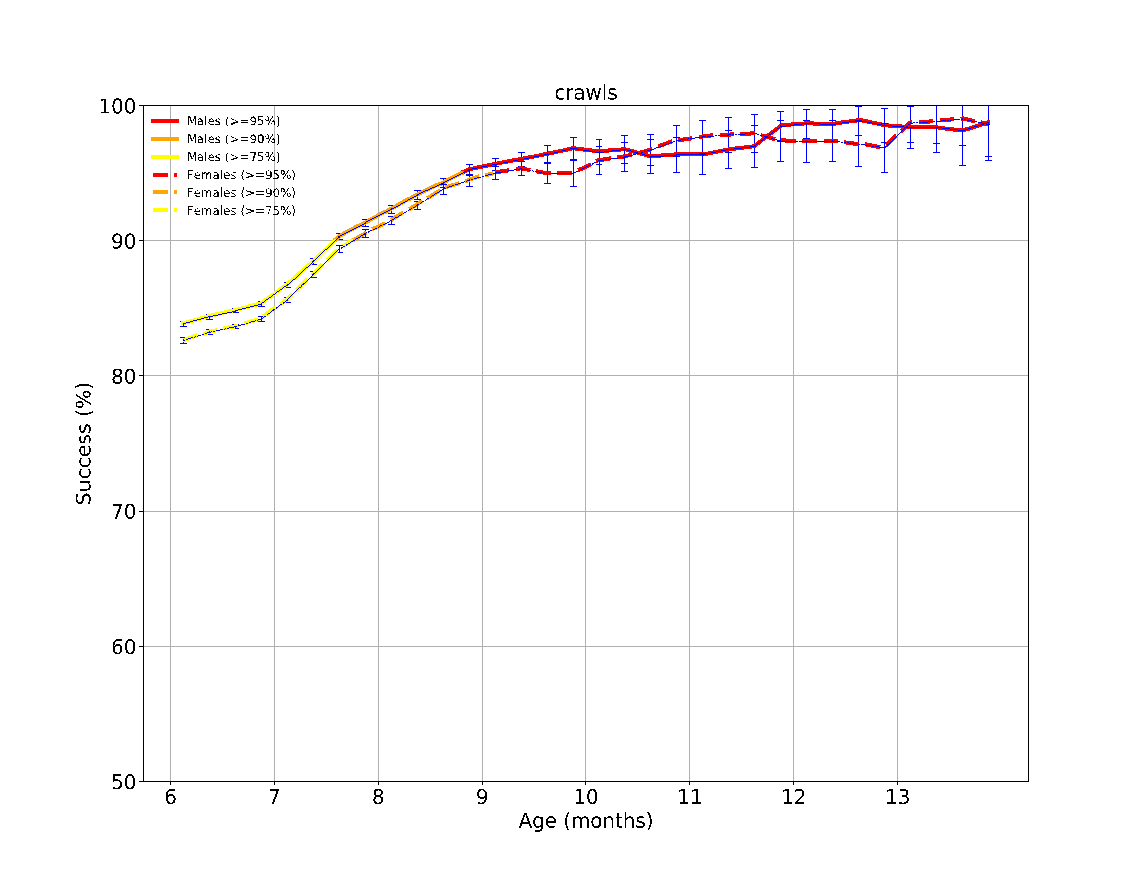

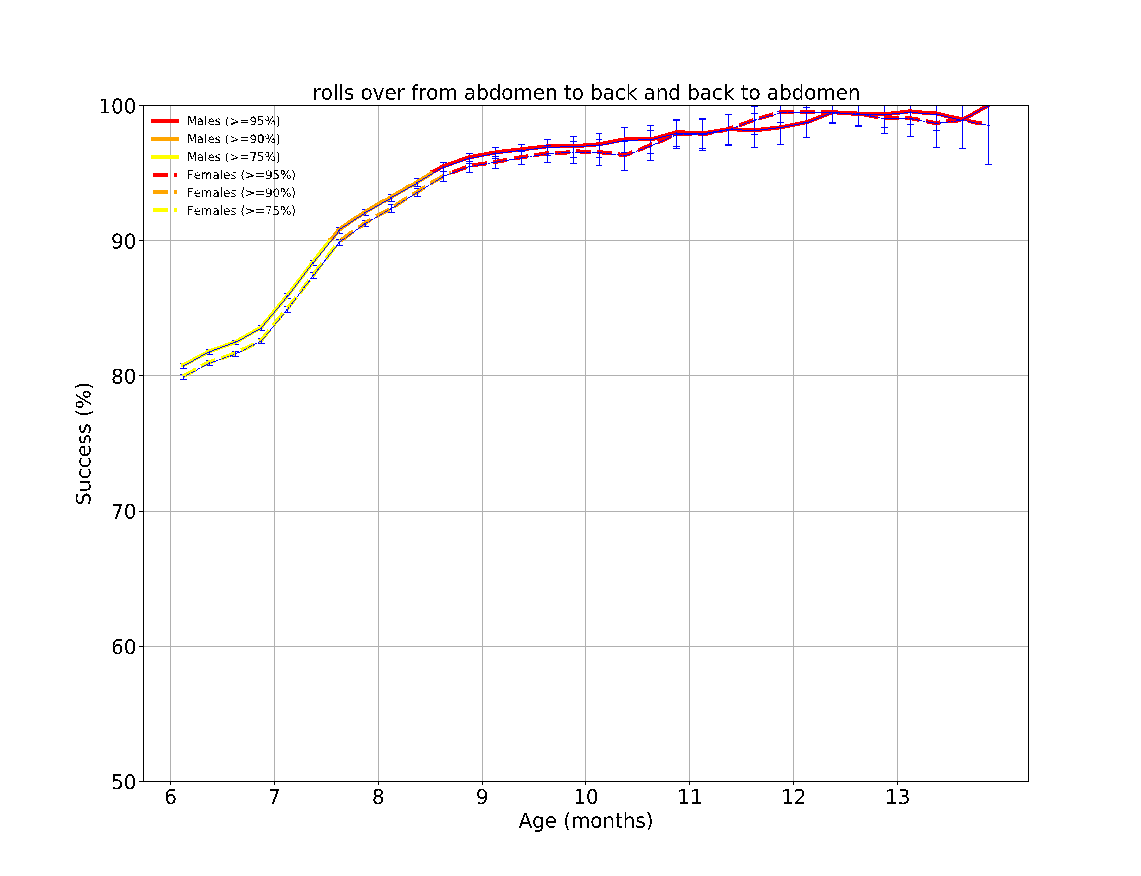

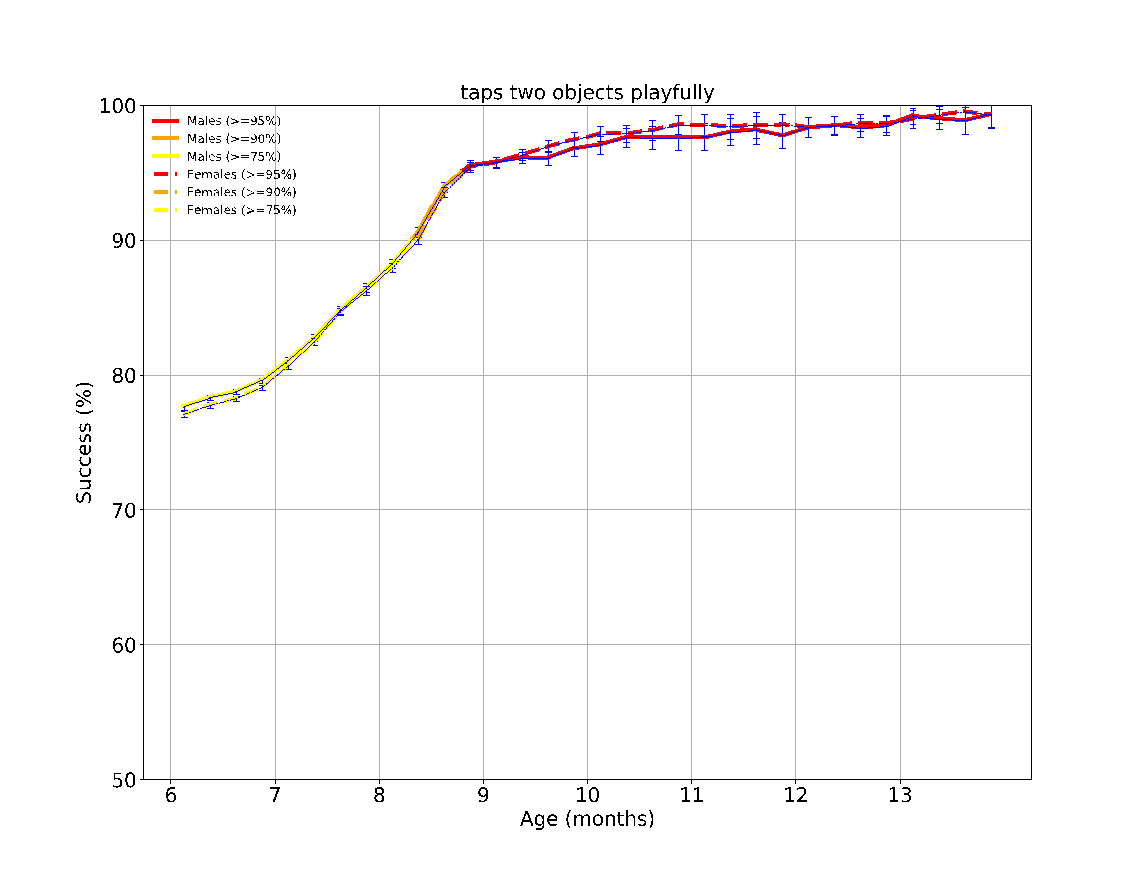

eFigure 4: The success rates over time of the different milestones evaluated between the ages 6-9 months. The achievement rates of < 75%, 75-90%, 90-95% and > 95% of children are indicated by the colors green, yellow, orange, and red, respectively. The error bars are 95% confidence intervals of a binomial proportion, where larger bars indicate smaller sample size.

**eFigure 5 - Task performance results over time according to sex at the age 9-12 months**
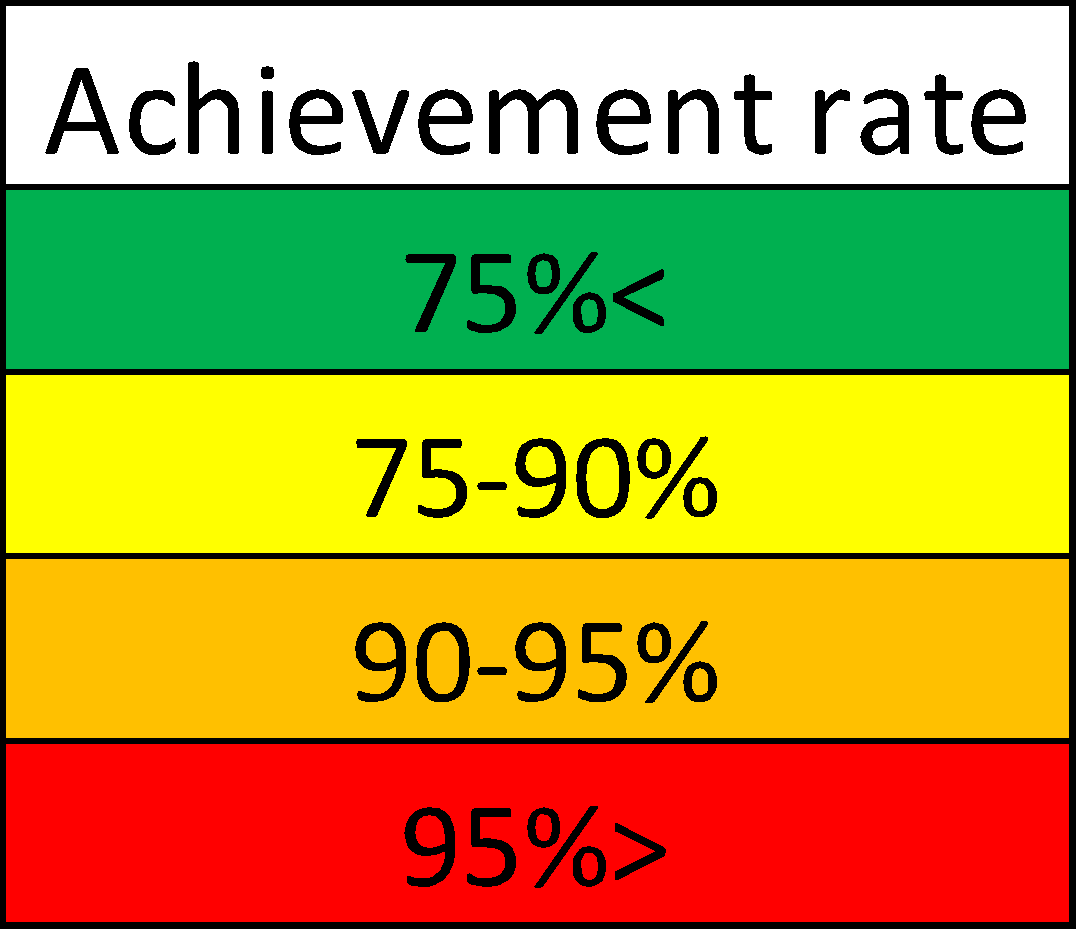

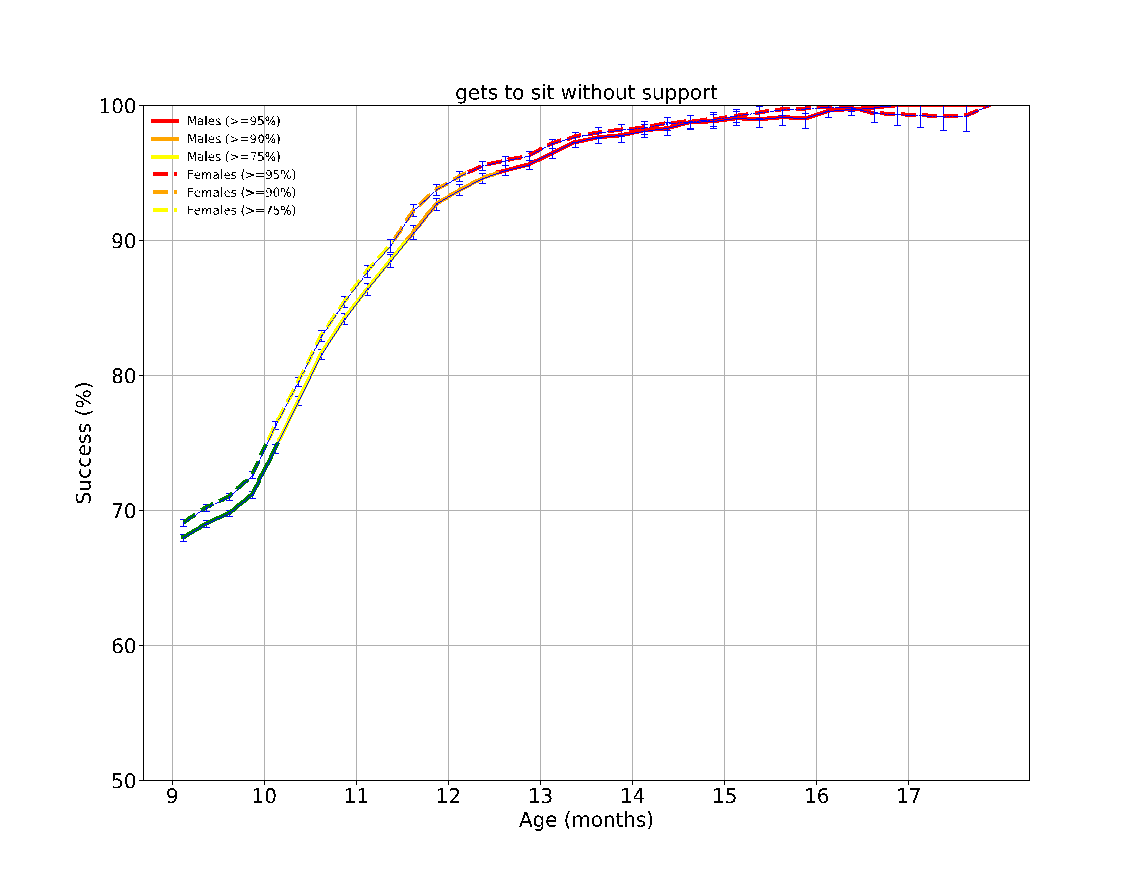

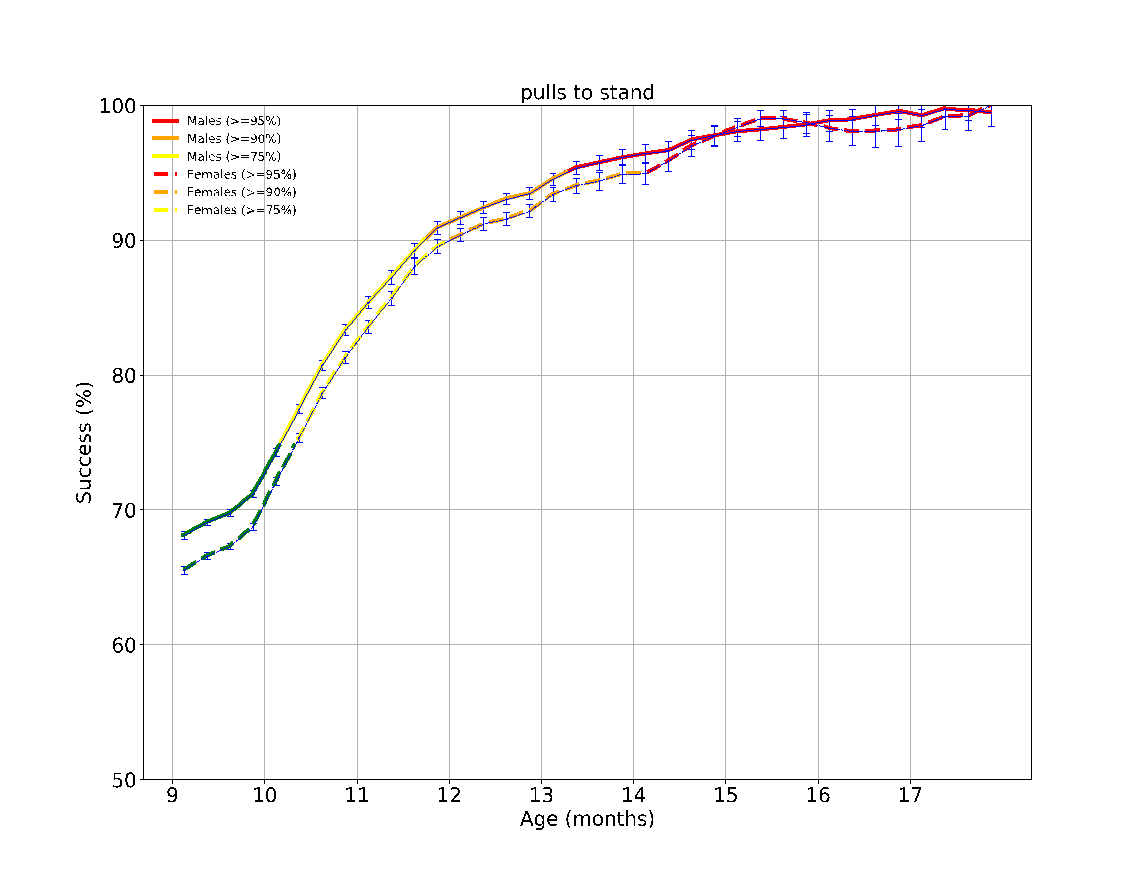

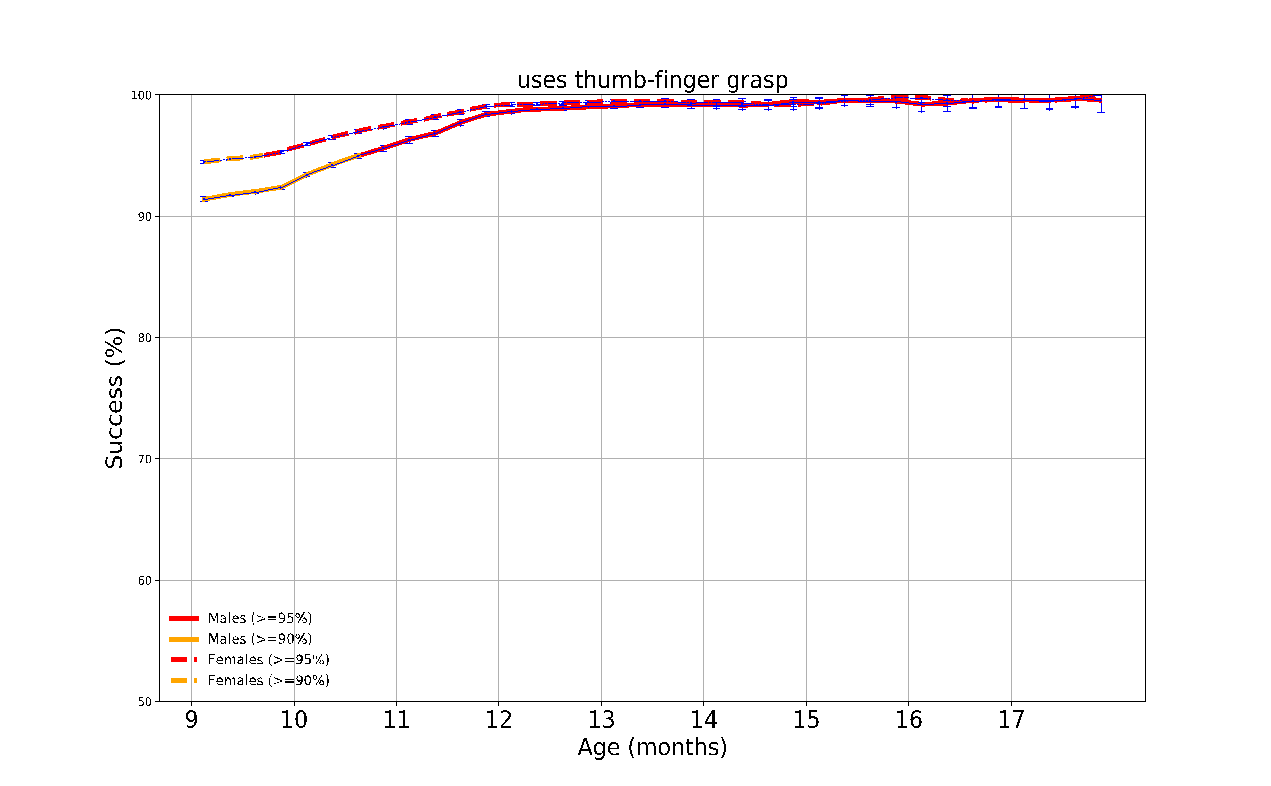

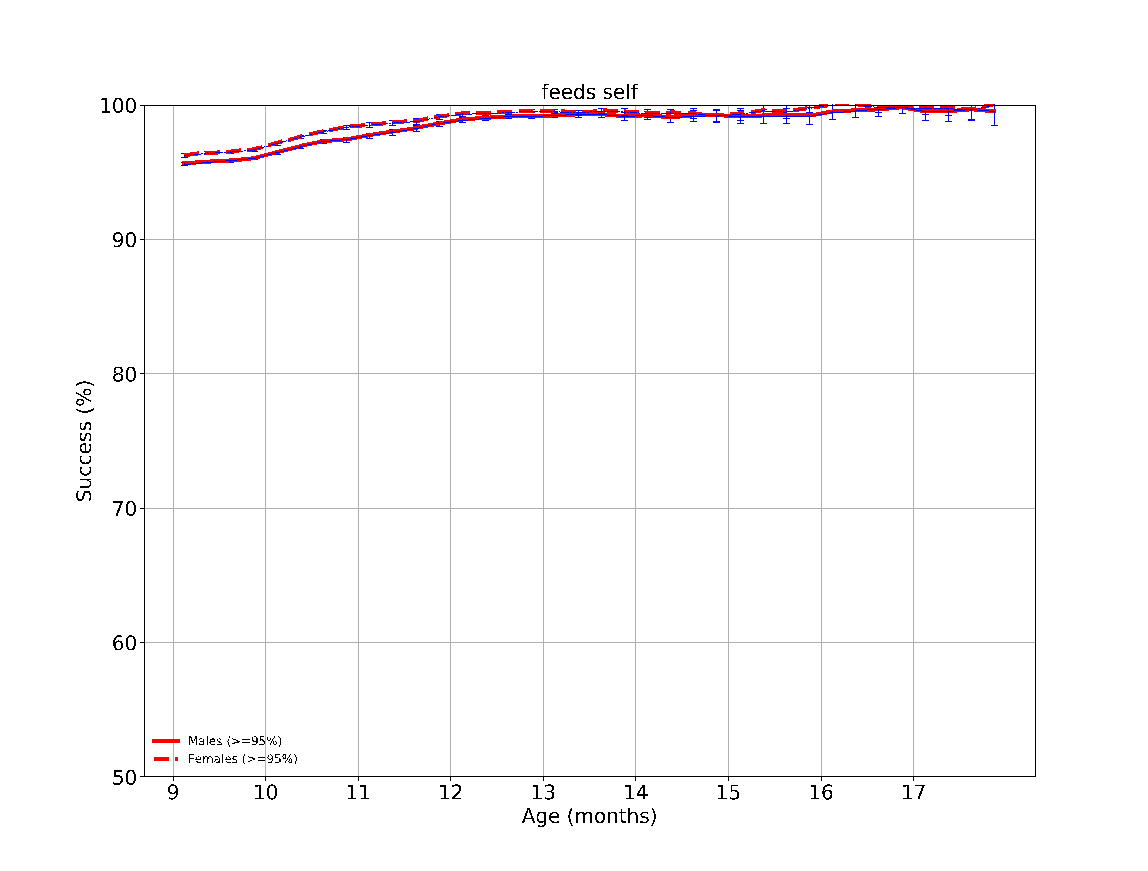

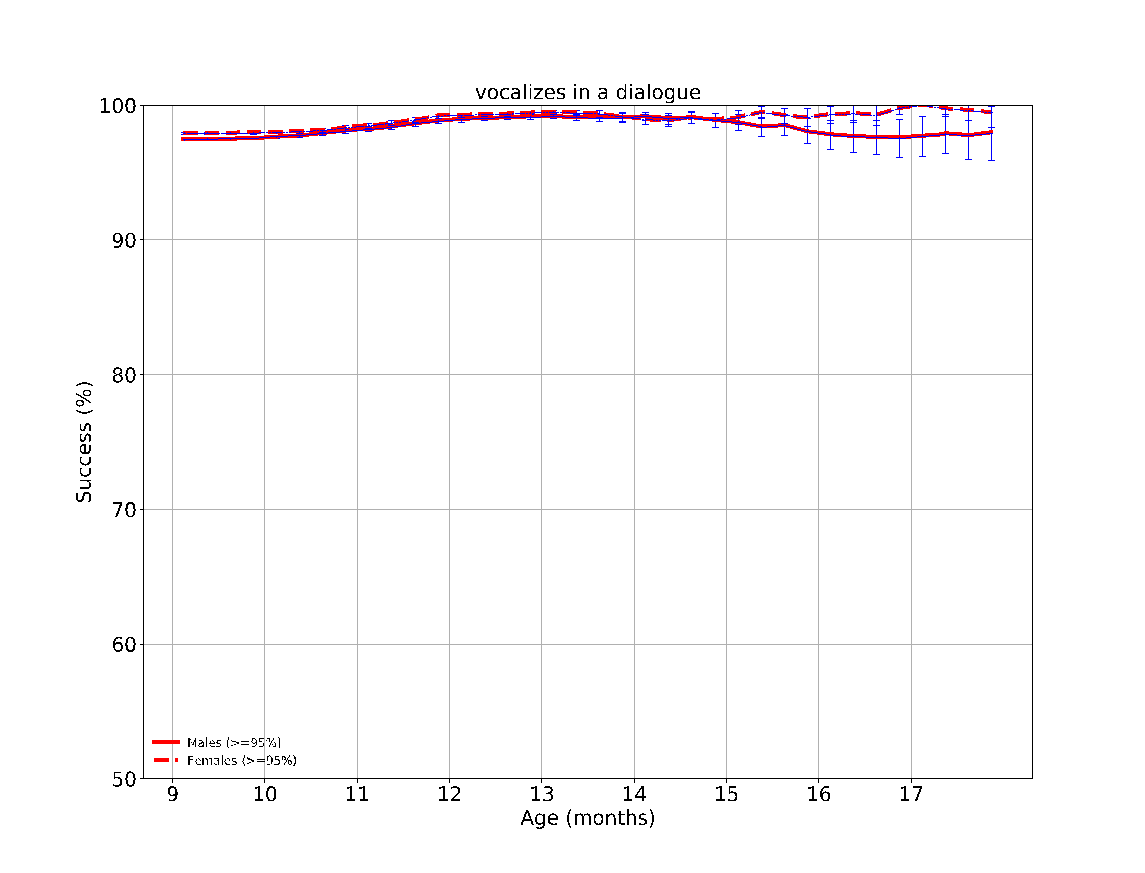

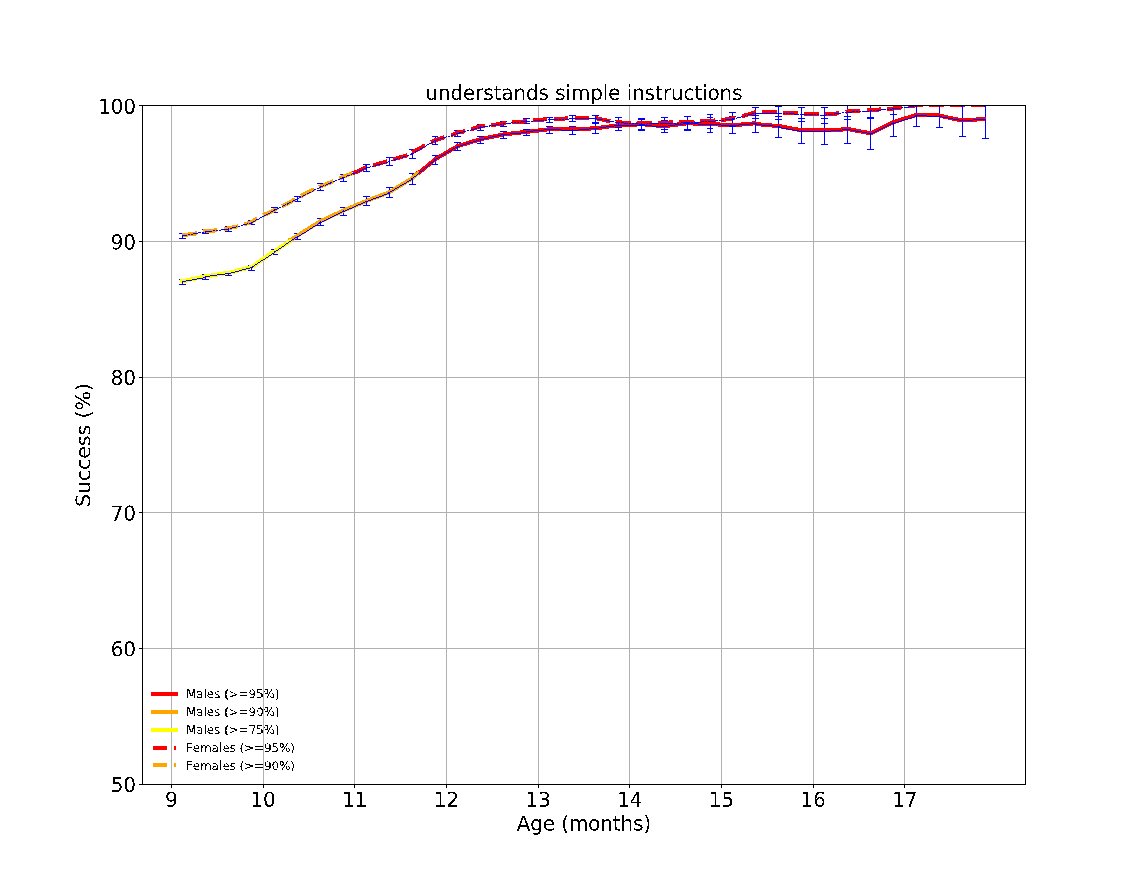

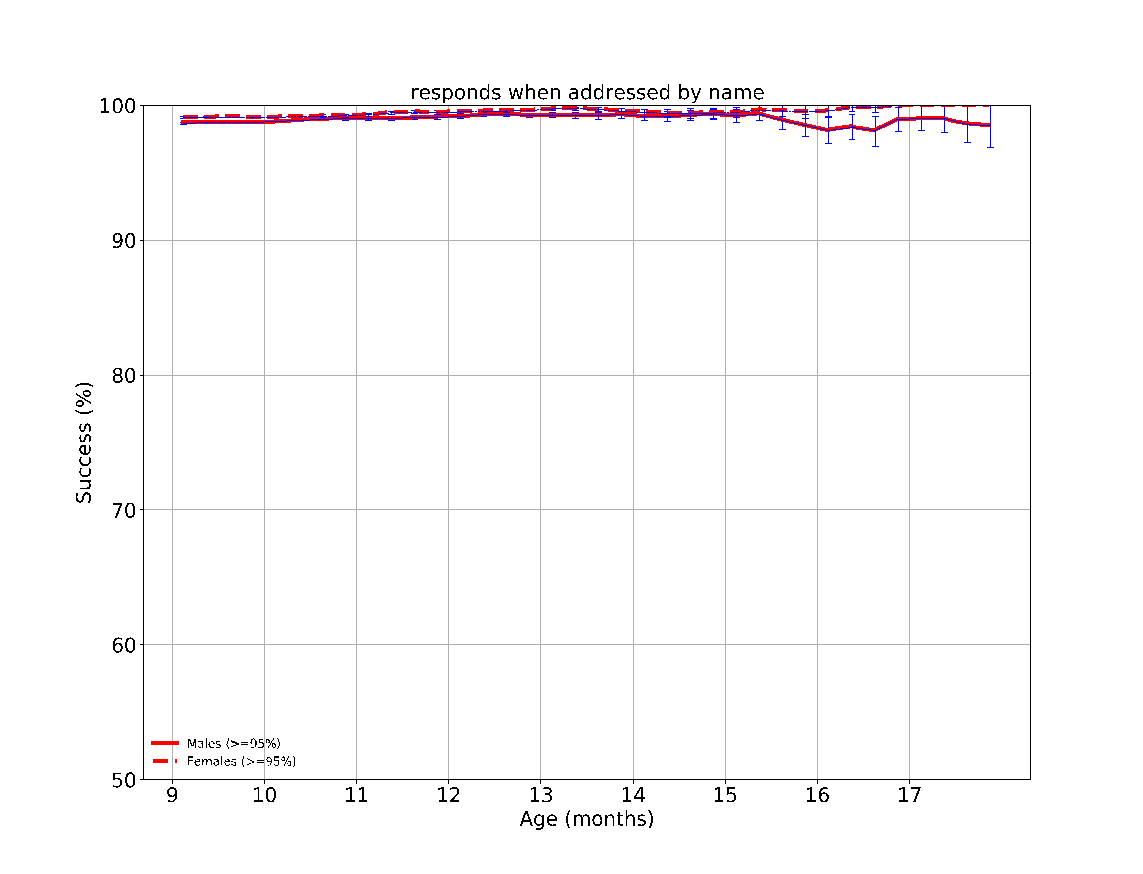

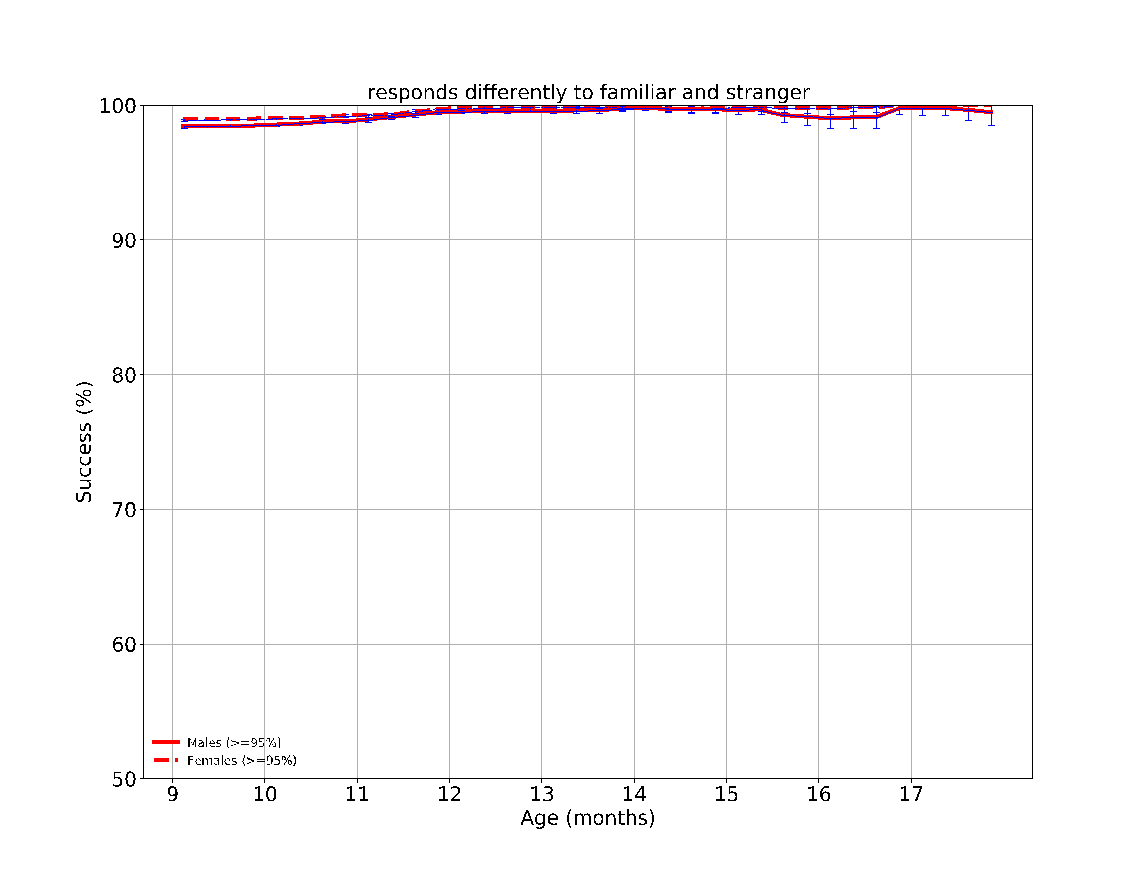

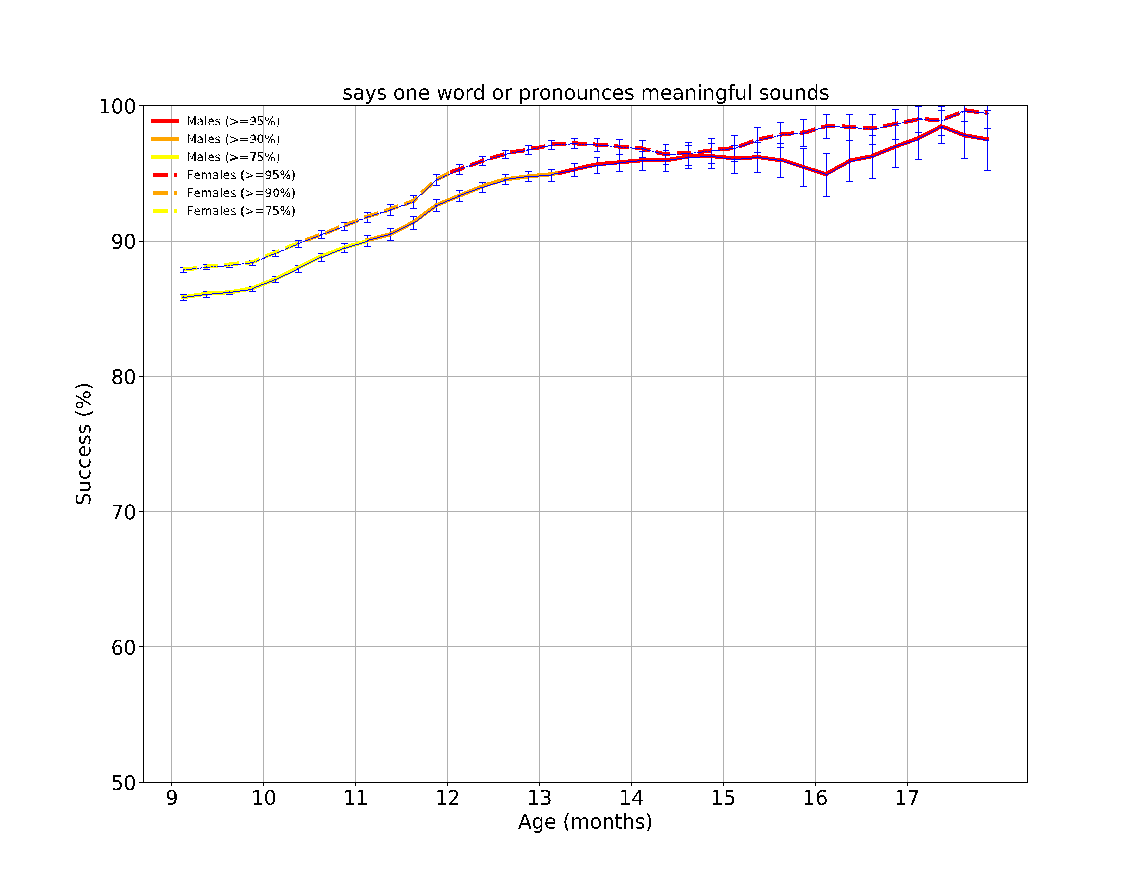

eFigure 5: The success rates over time of the different milestones evaluated between the ages 9-12 months. The achievement rates of < 75%, 75-90%, 90-95% and > 95% of children are indicated by the colors green, yellow, orange and red, respectively. The error bars are 95% confidence intervals of a binomial proportion, where larger bars indicate smaller sample size.

**eFigure 6 - Task performance results over time according to sex at the age 12-18 months**
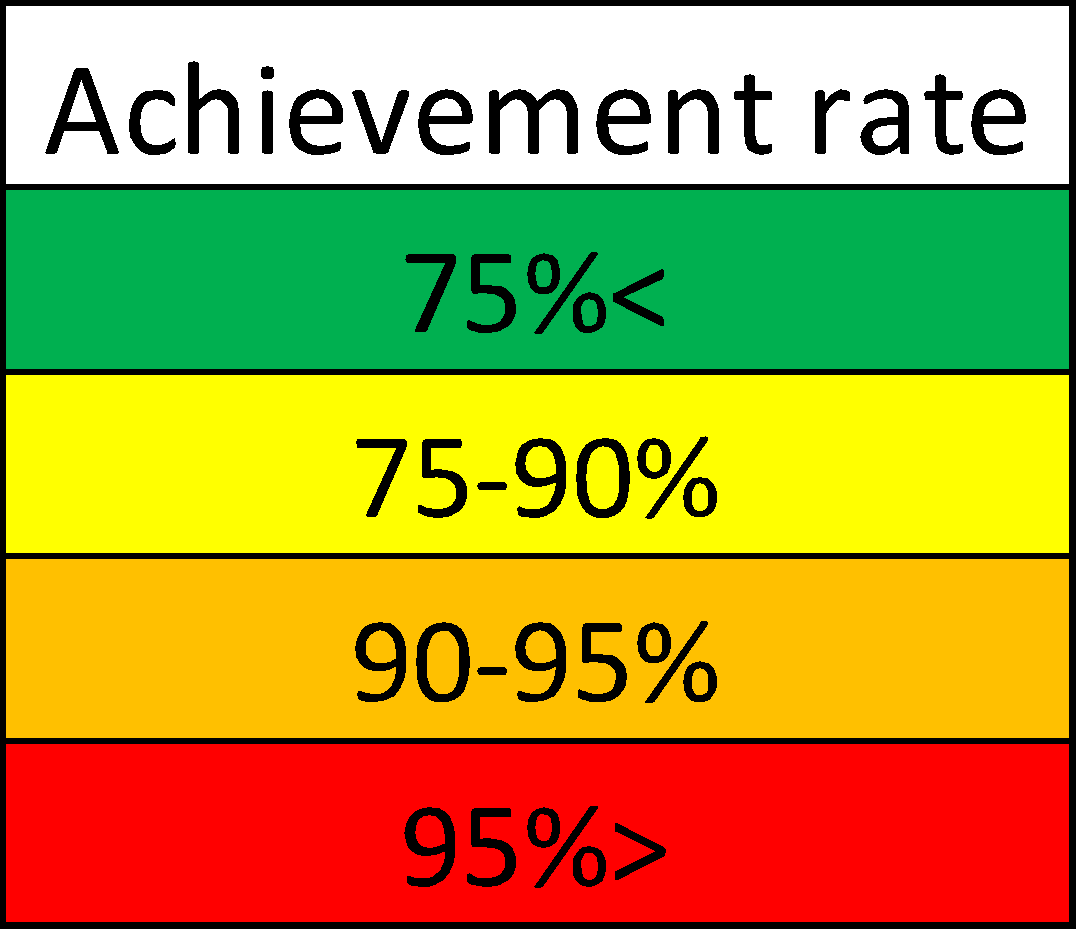

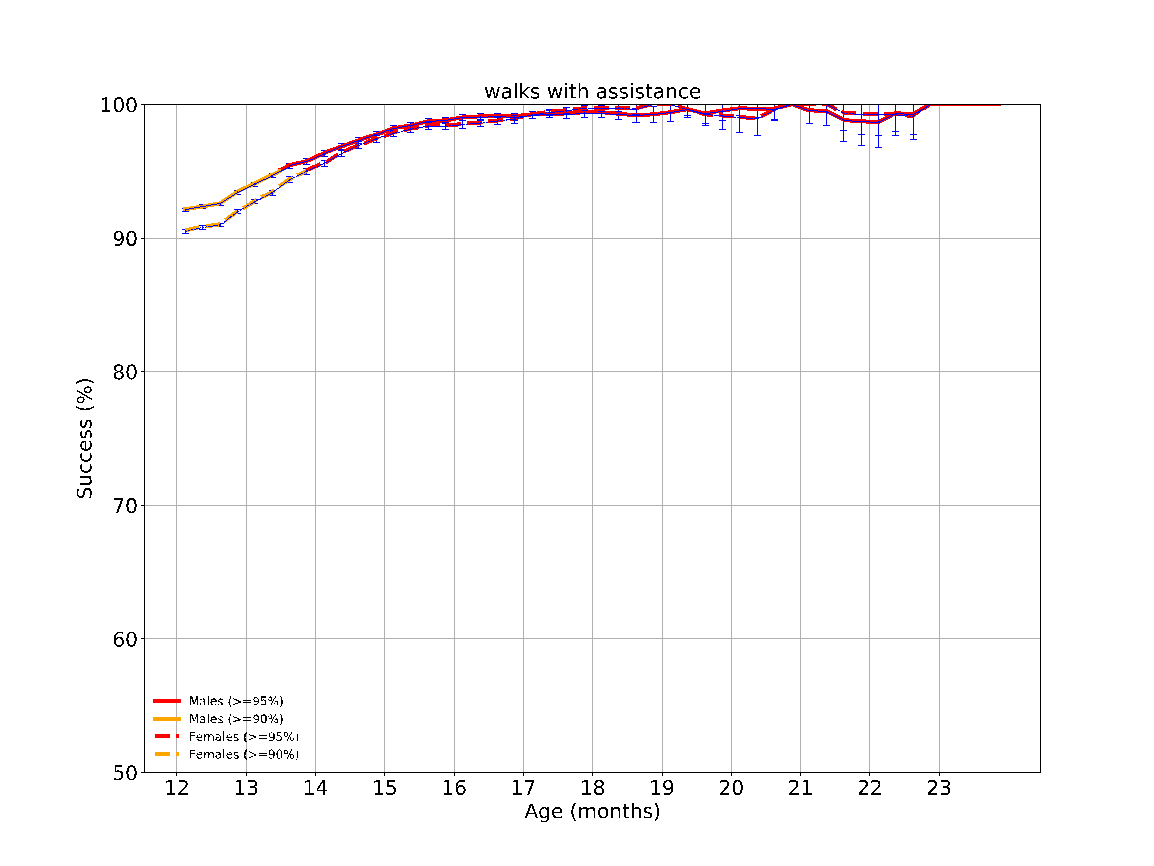

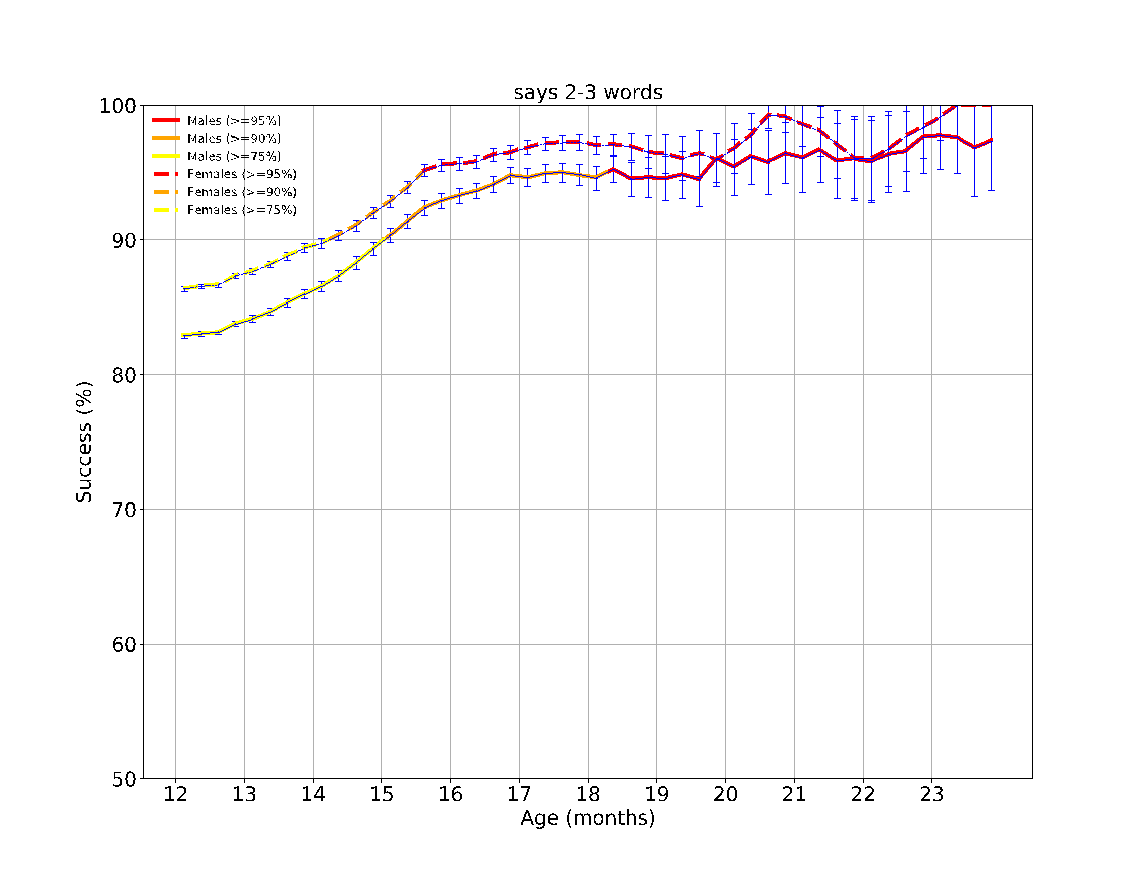

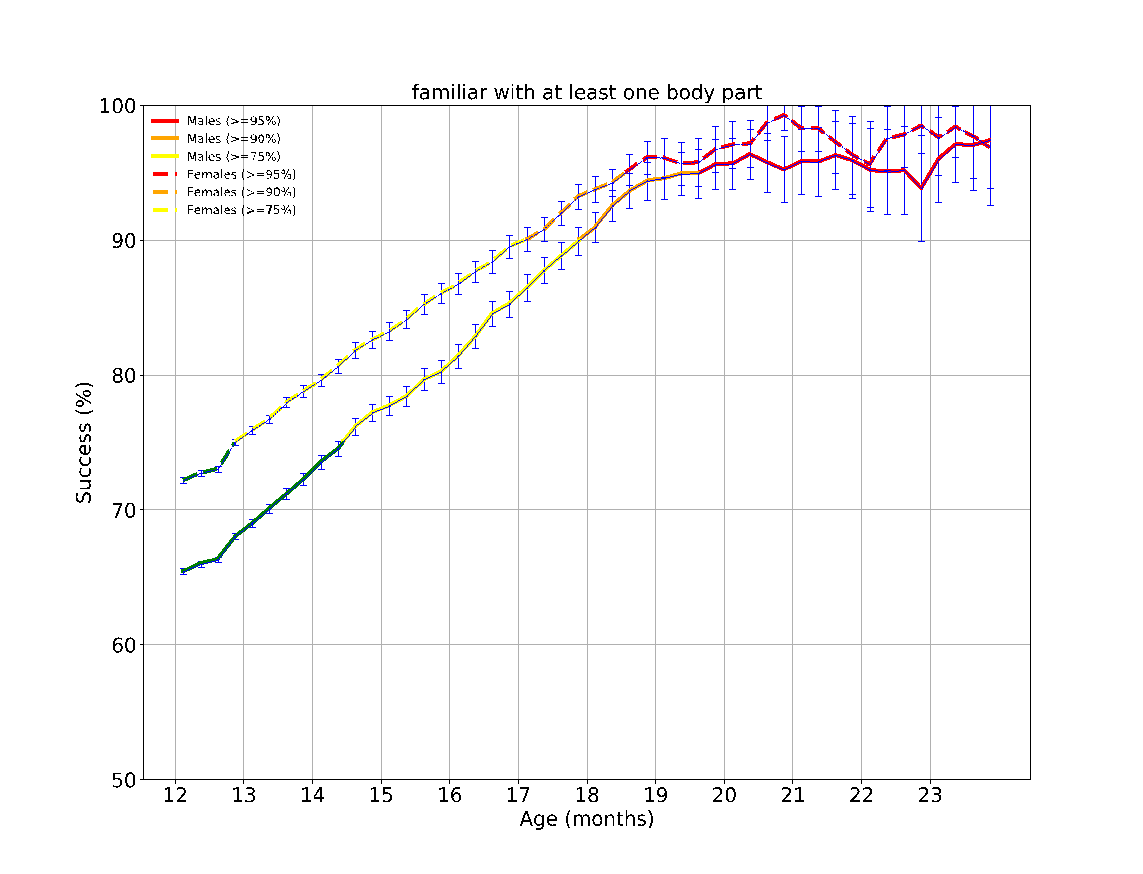

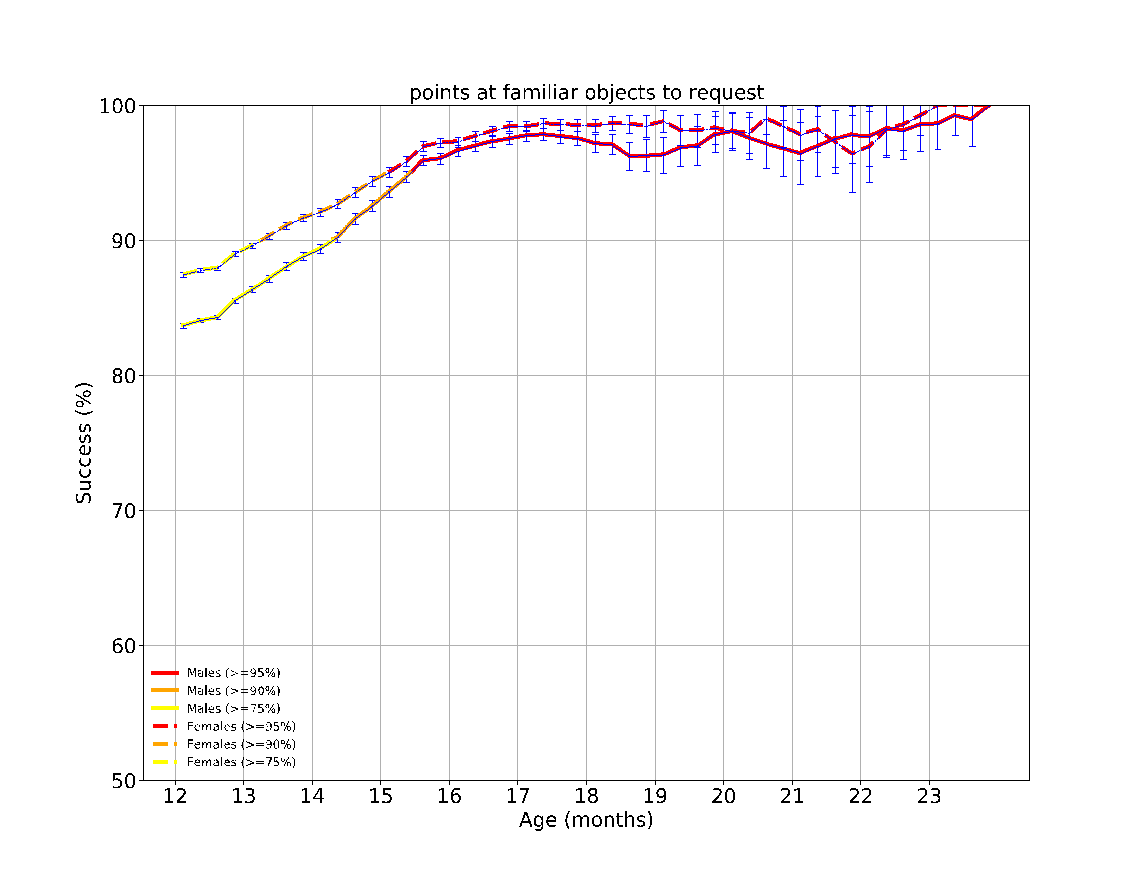

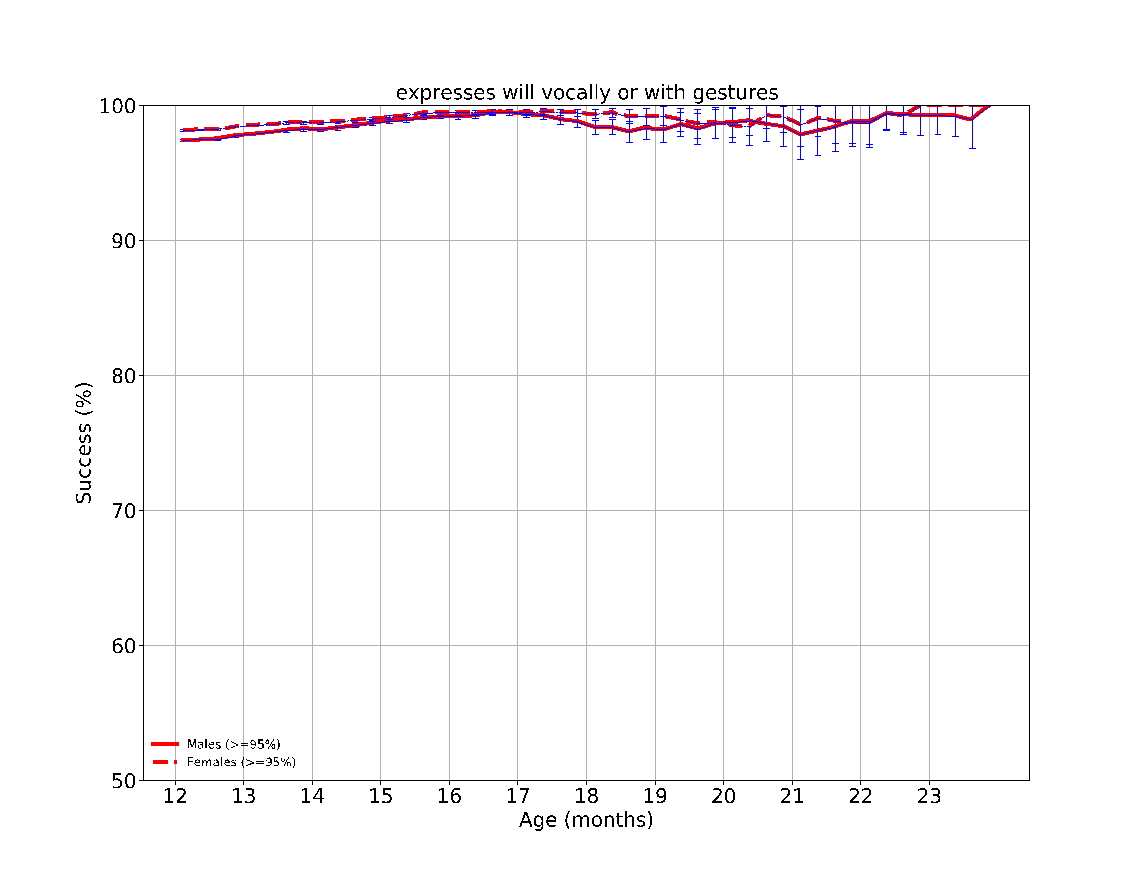

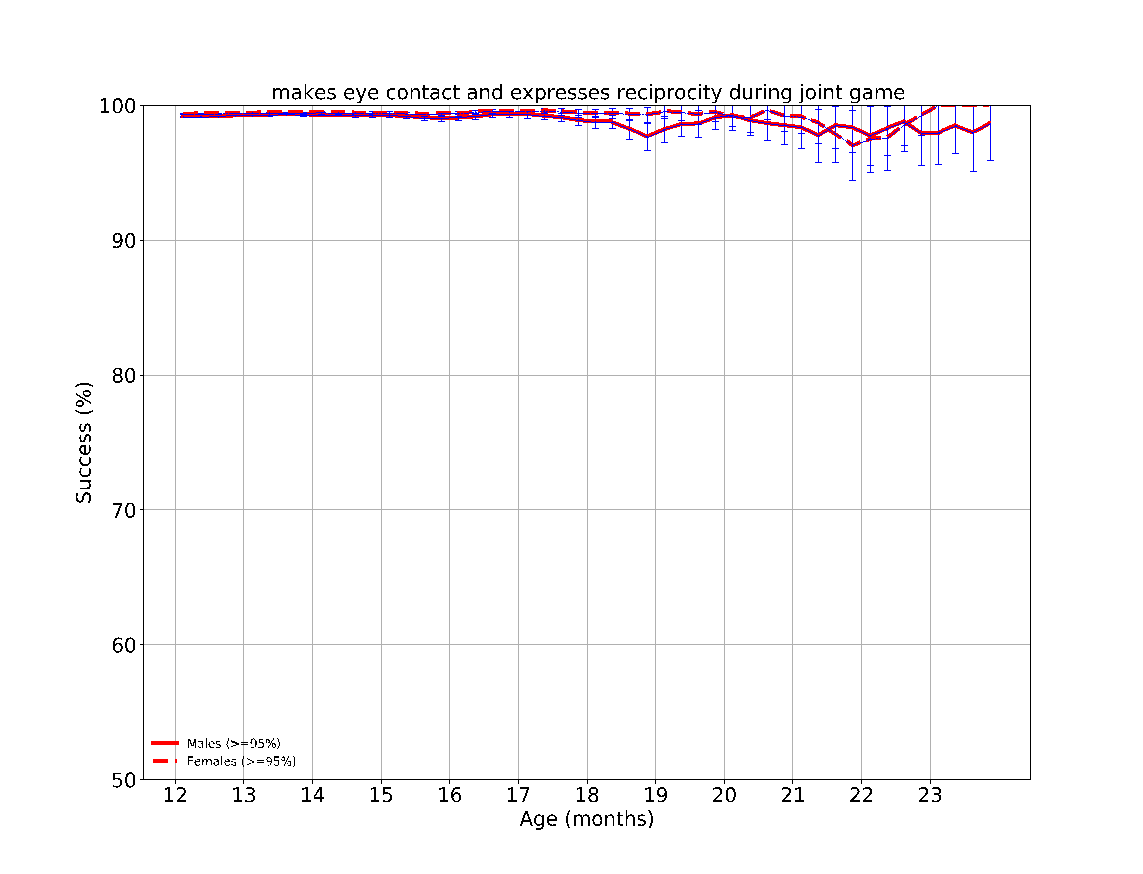

eFigure 6: The success rates over time of the different milestones evaluated between the ages 12-18 months. The achievement rates of < 75%, 75-90%, 90-95% and > 95% of children are indicated by the colors green, yellow, orange and red, respectively. The error bars are 95% confidence intervals of a binomial proportion, where larger bars indicate smaller sample size.

**eFigure 7 - Task performance results over time according to sex at the age 18-24 months**
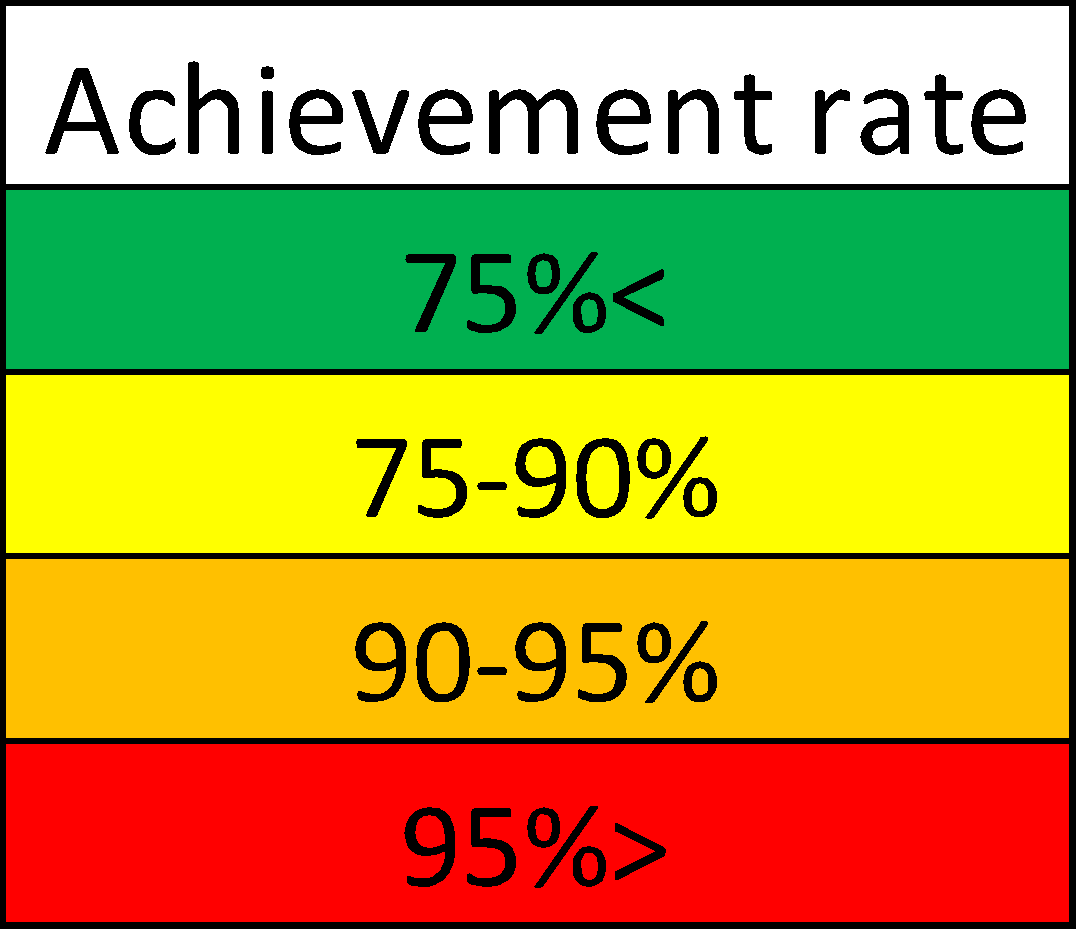

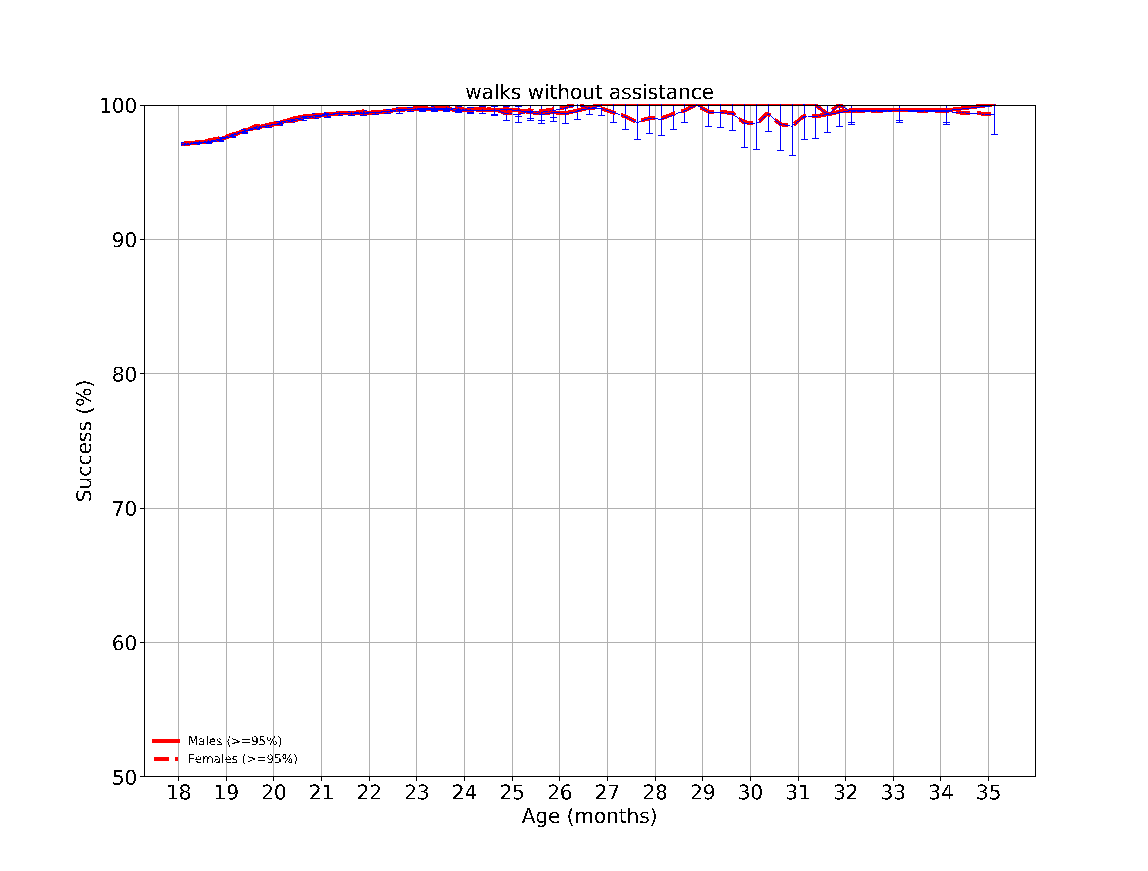

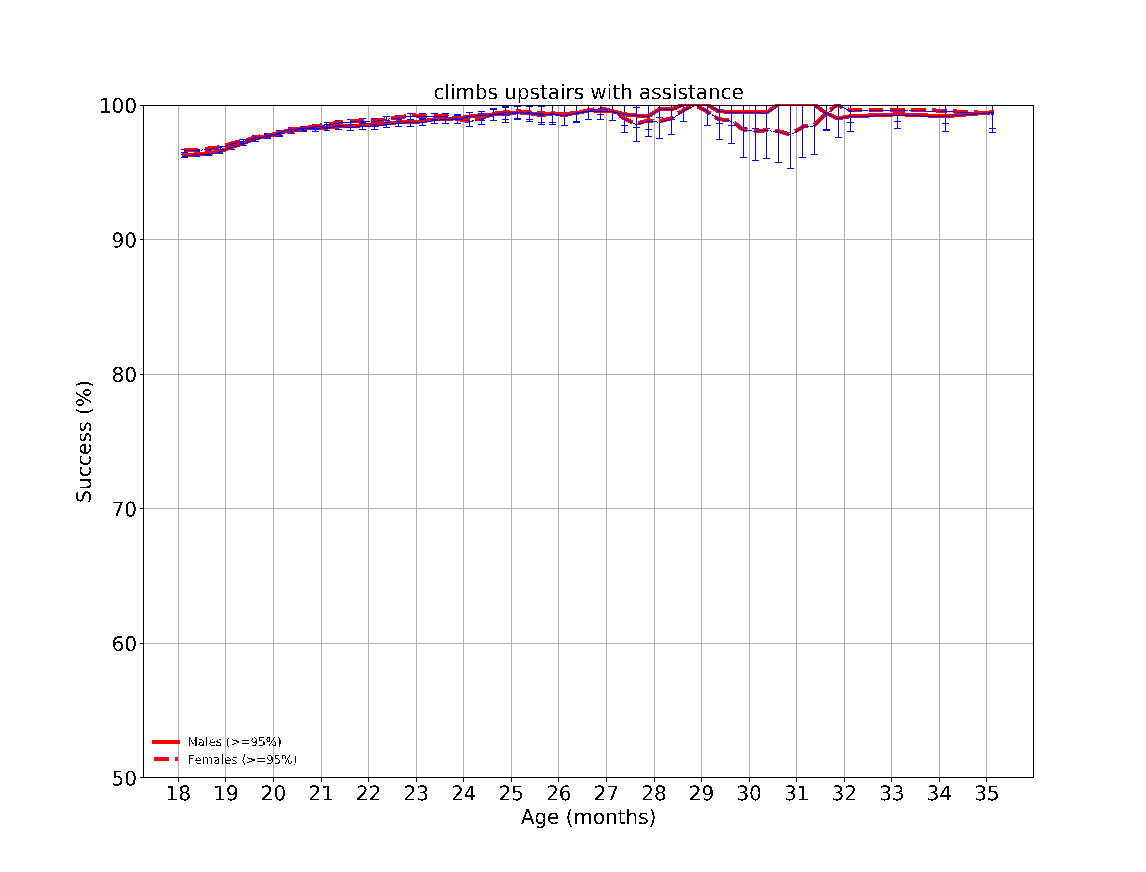

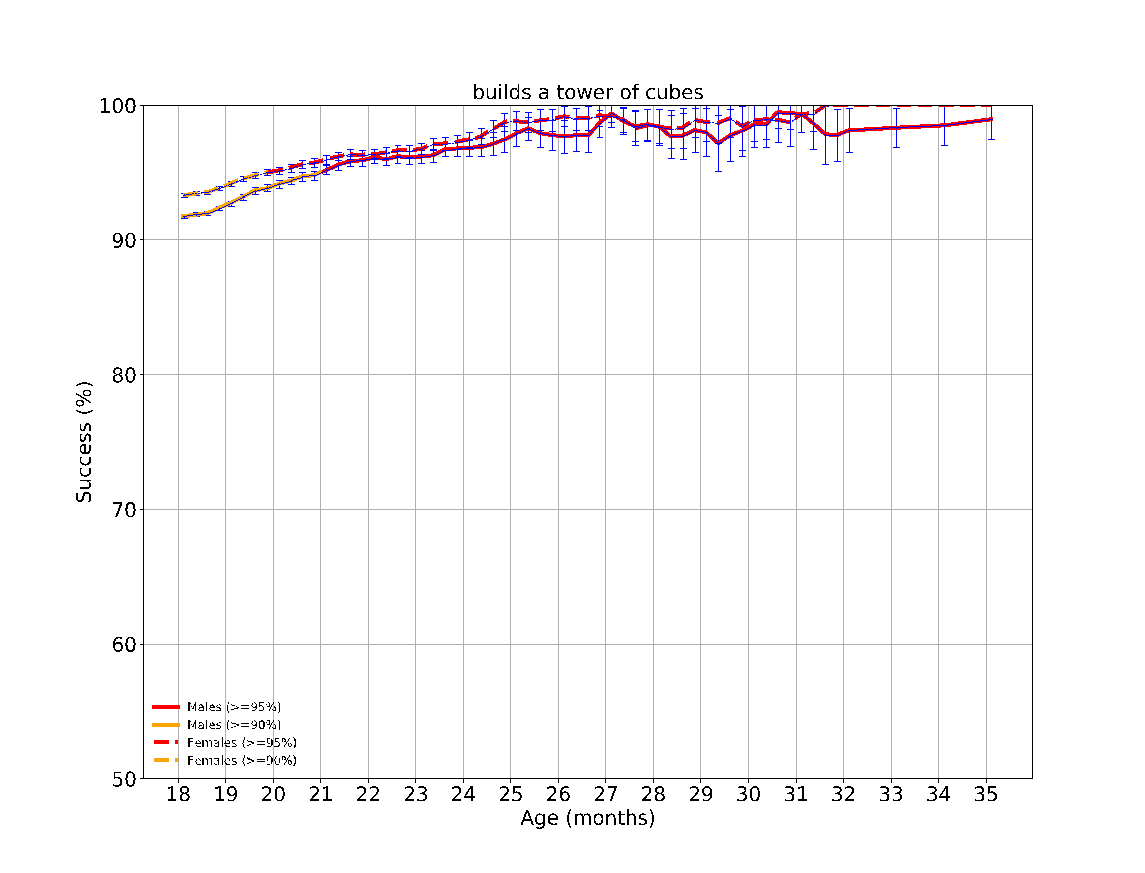

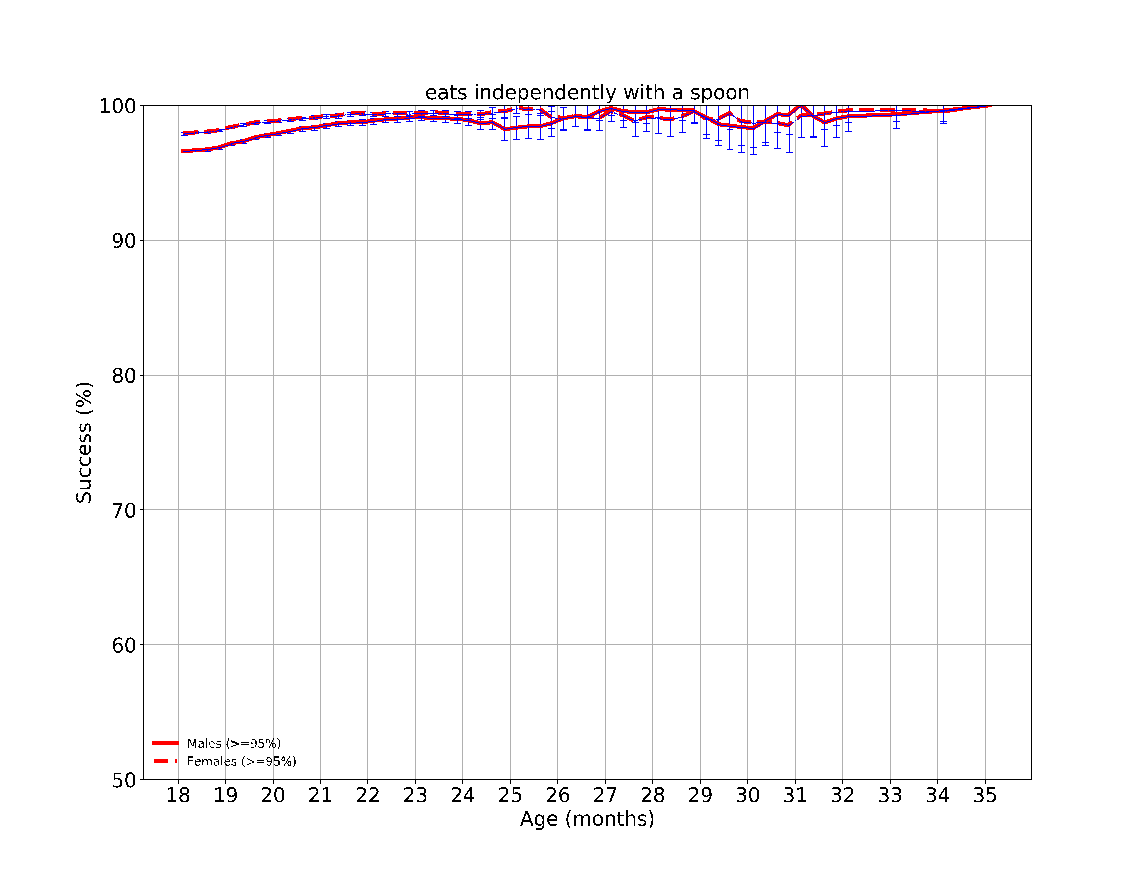

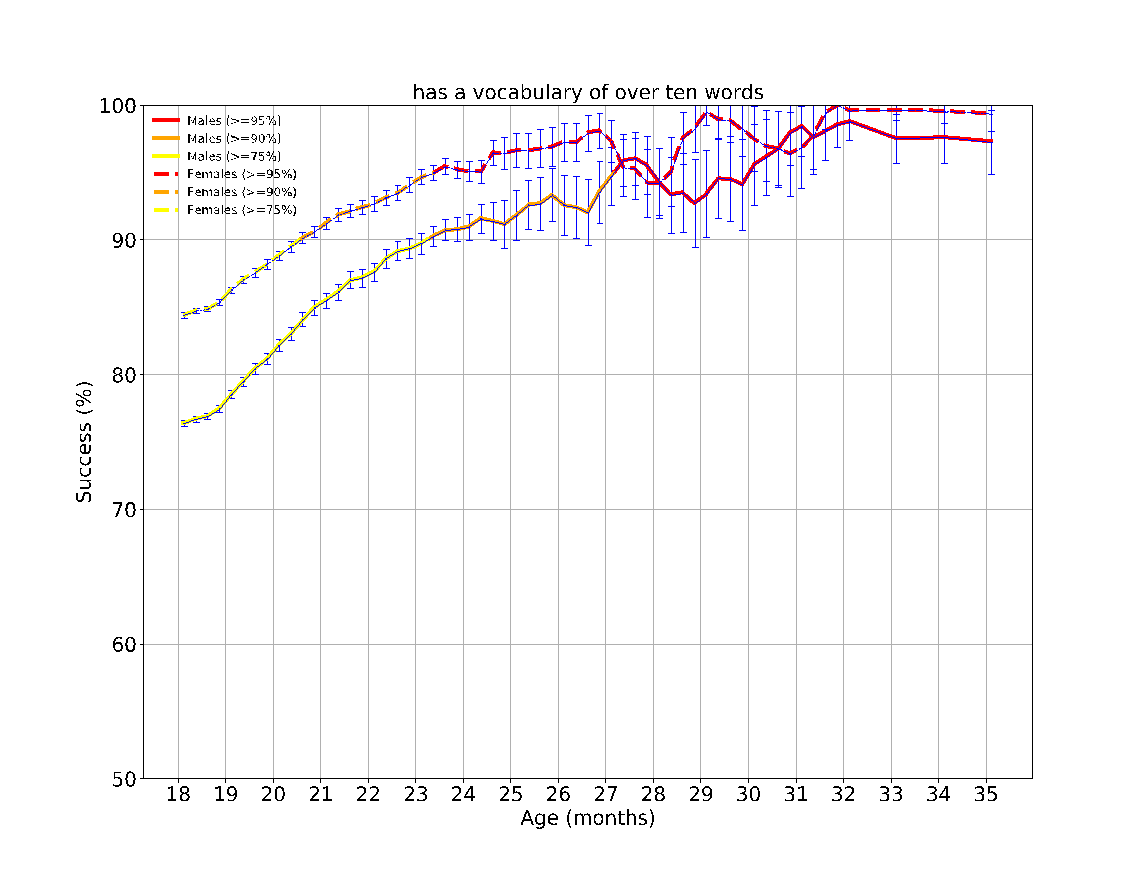

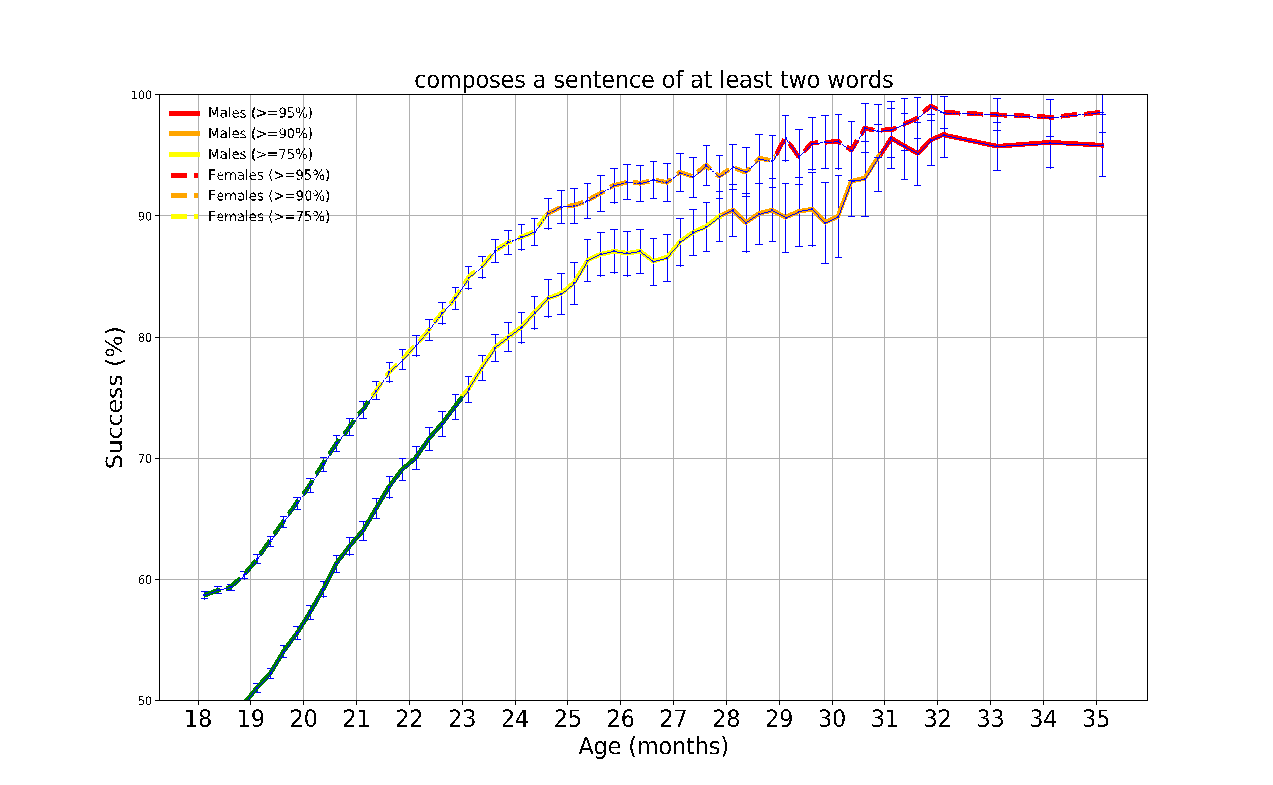

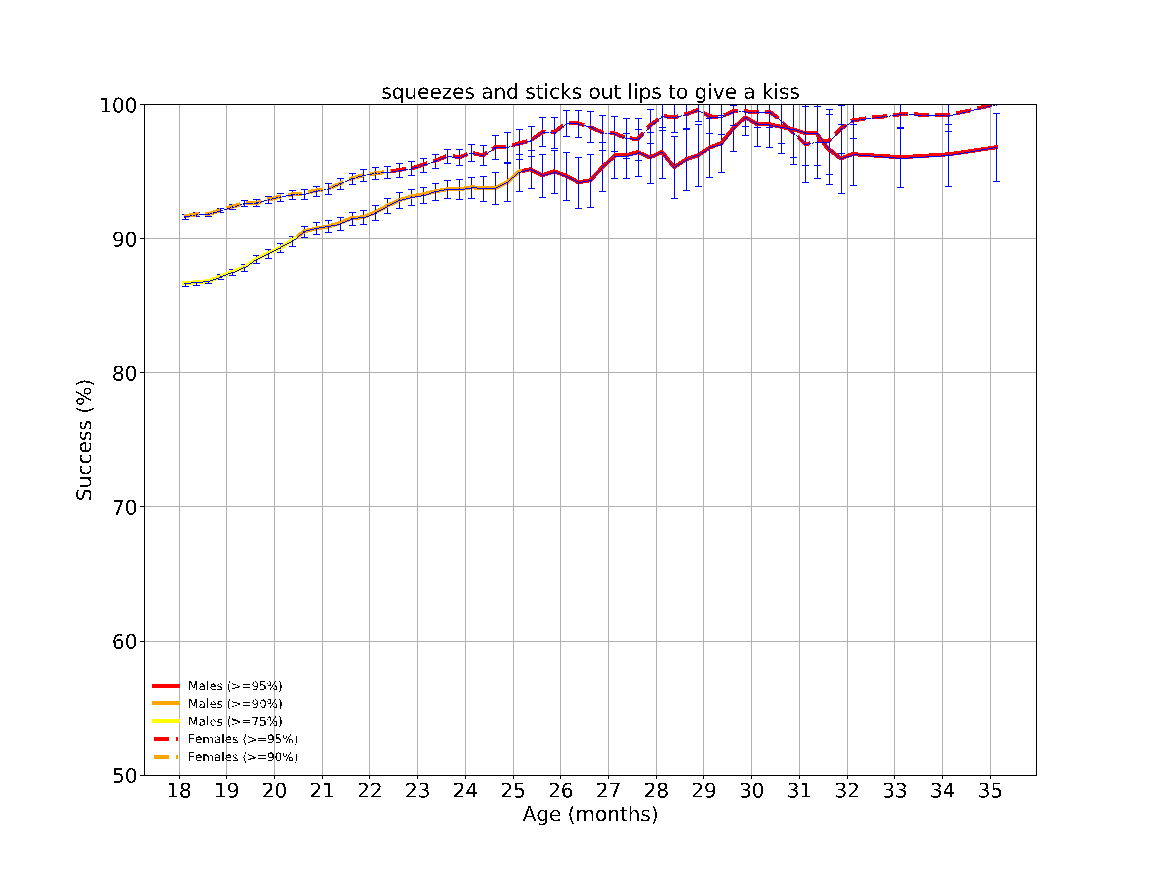

eFigure 7: The success rates over time of the different milestones evaluated between the ages 18-24 months. The achievement rates of < 75%, 75-90%, 90-95% and > 95% of children are indicated by the colors green, yellow, orange and red, respectively. The error bars are 95% confidence intervals of a binomial proportion, where larger bars indicate smaller sample size.

**eFigure 8 - Task performance results over time according to sex at the age 2-3 years (24-36 months)**
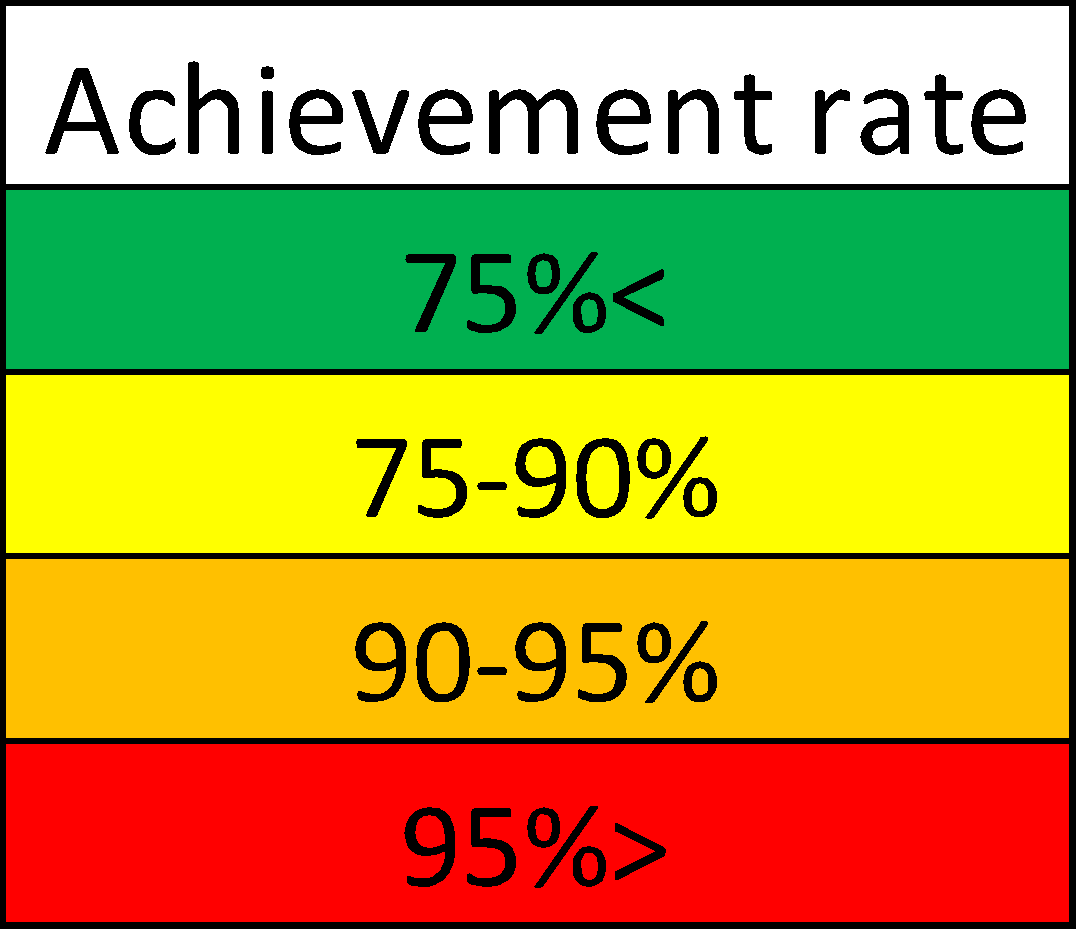

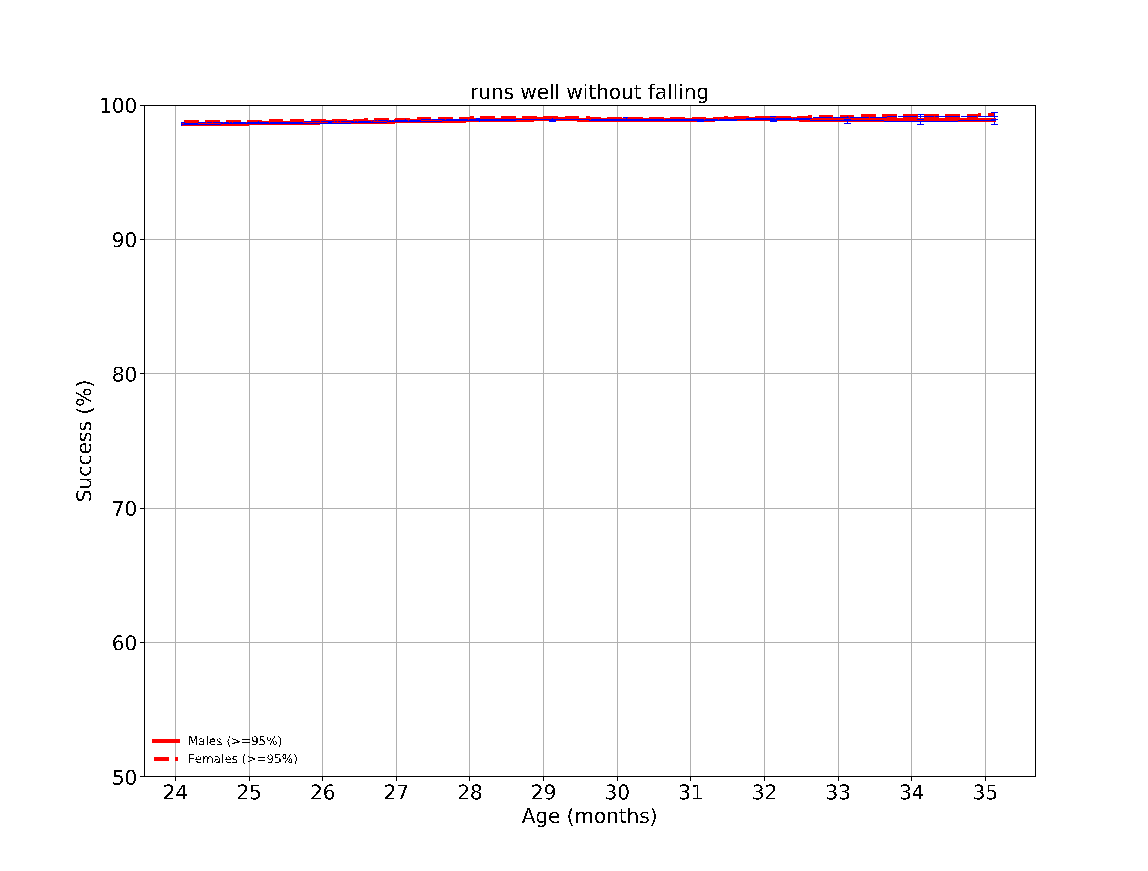

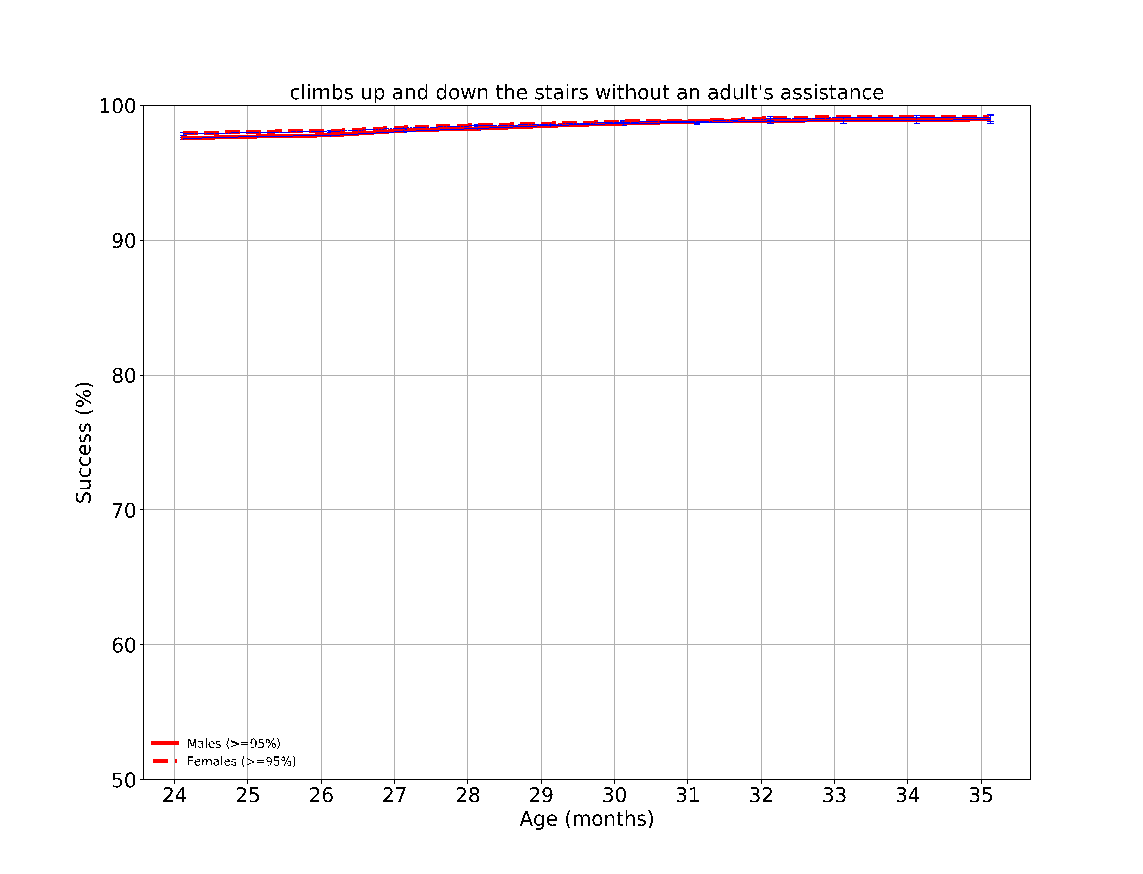

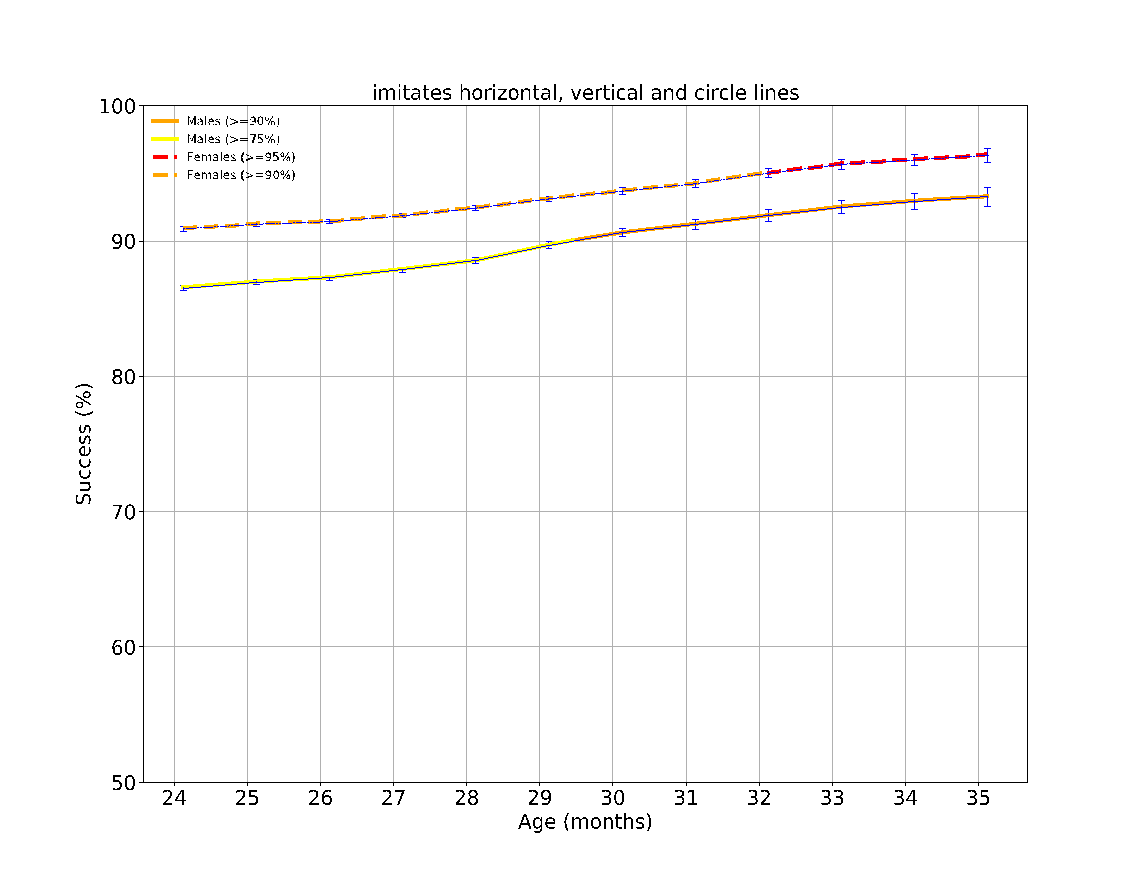

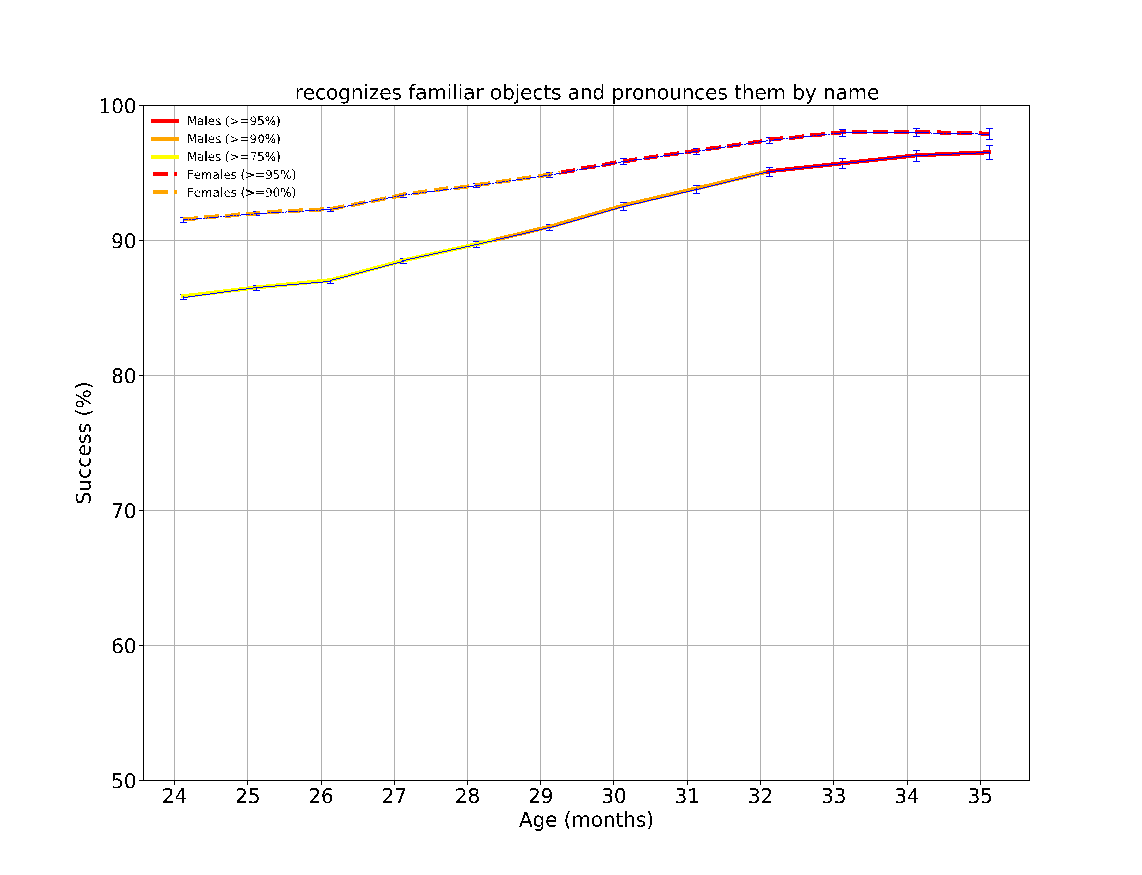

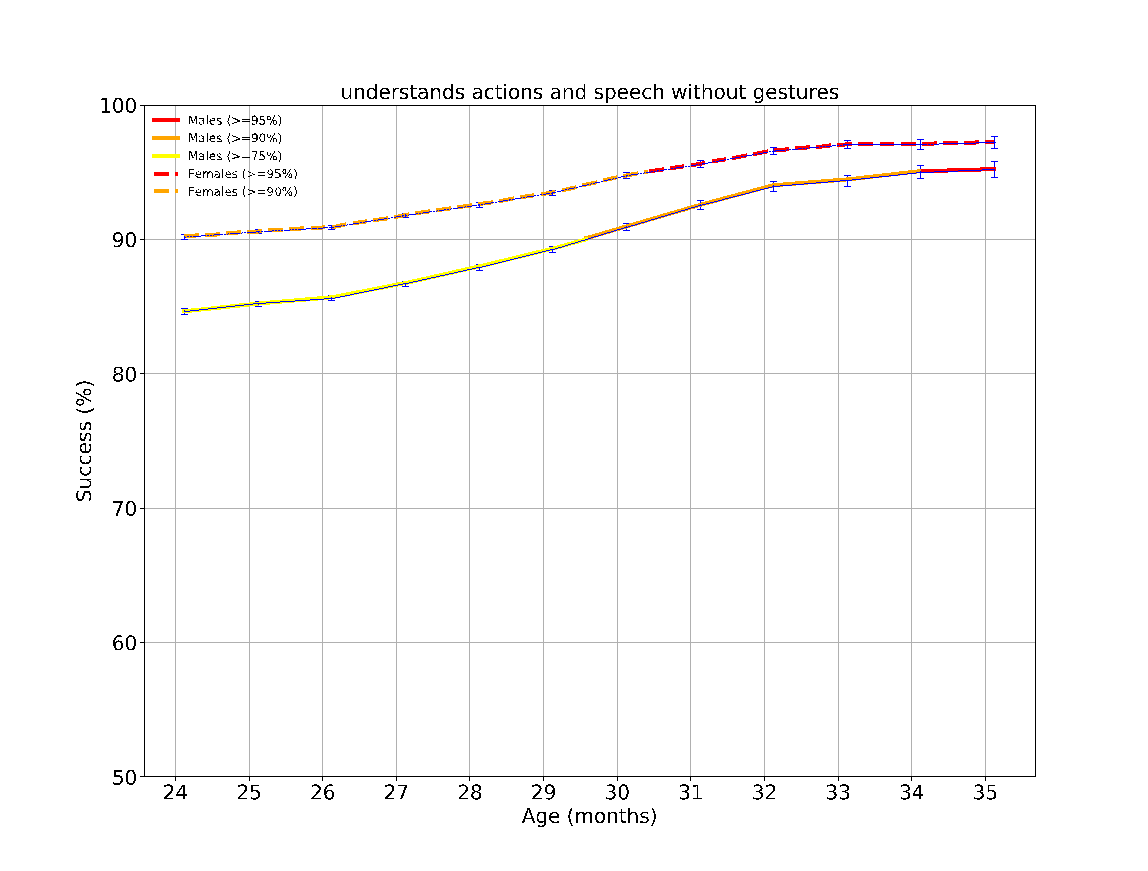

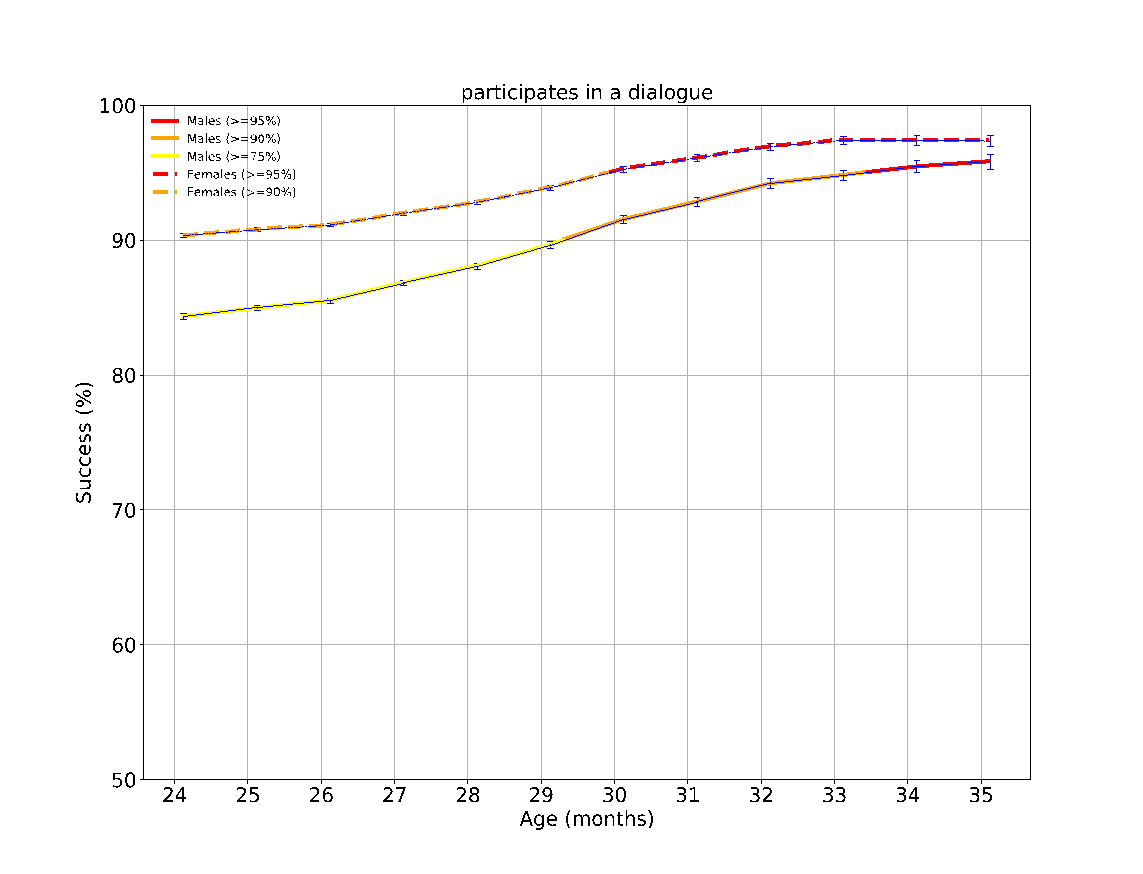

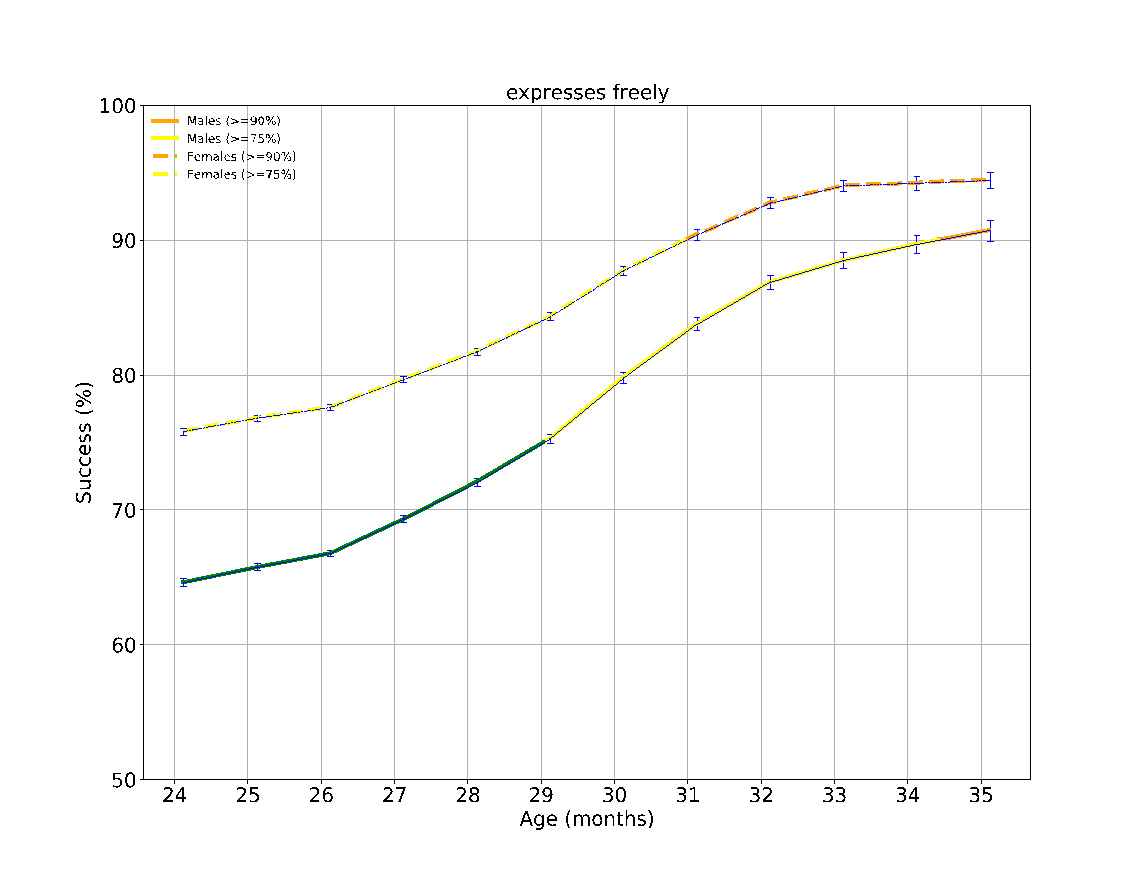

eFigure 8: The success rates over time of the different milestones evaluated between the ages 24-36 months. The achievement rates of < 75%, 75-90%, 90-95% and > 95% of children are indicated by the colors green, yellow, orange and red, respectively. The error bars are 95% confidence intervals of a binomial proportion, where larger bars indicate smaller sample size.

**eFigure 9 - Task performance results over time according to sex at the age 3-4 years (36-48 months)**
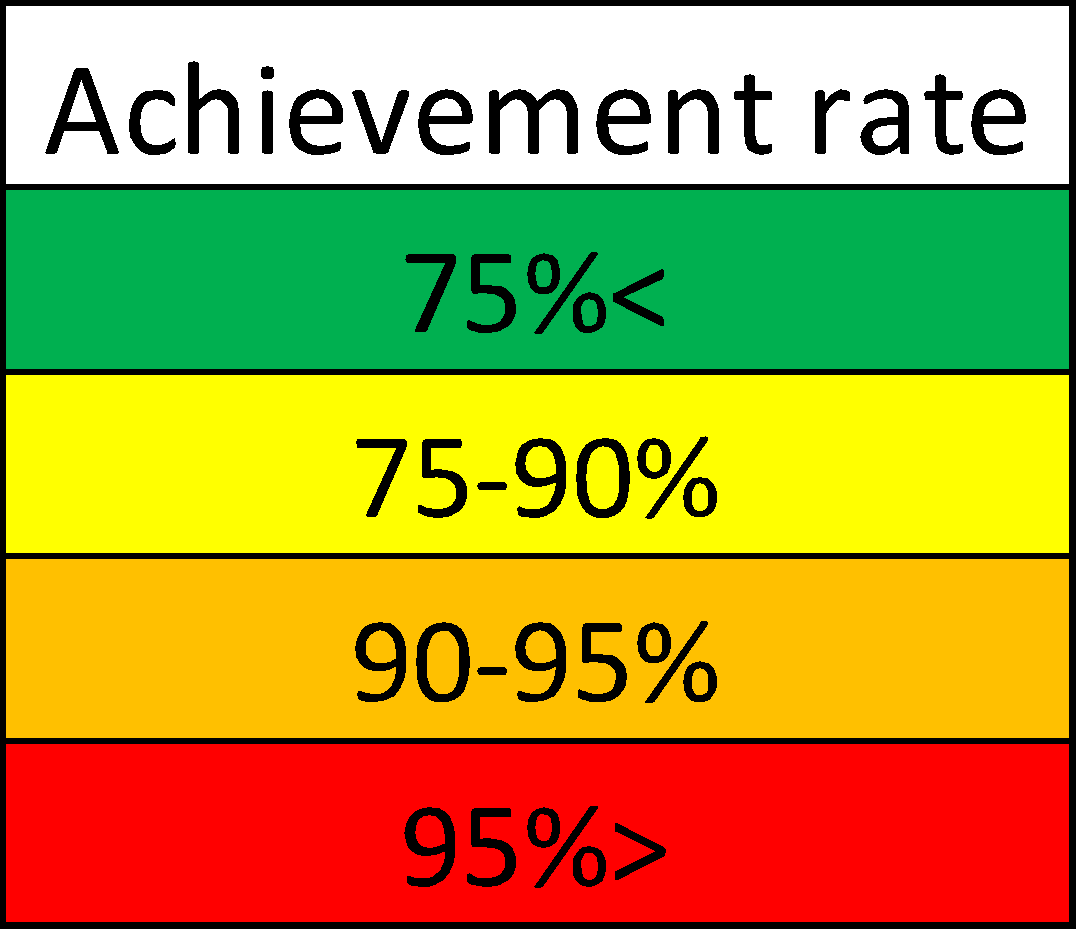

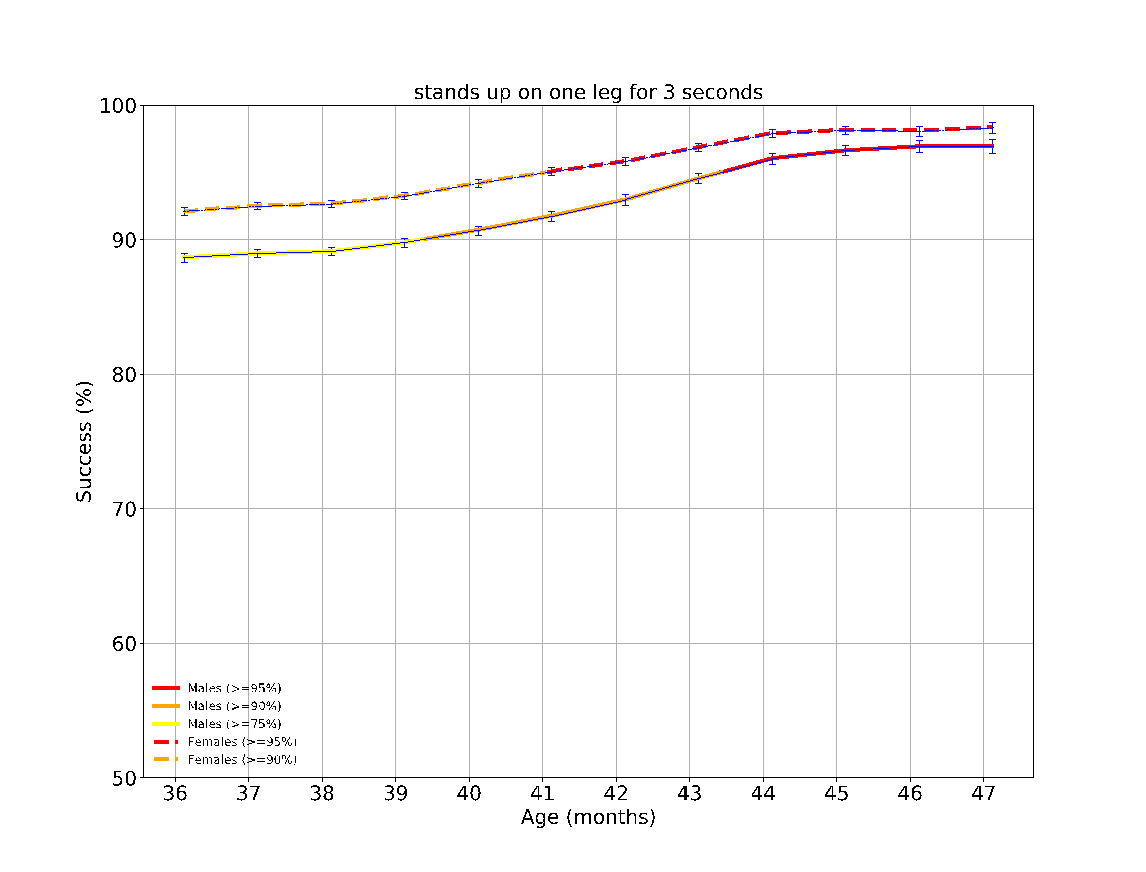

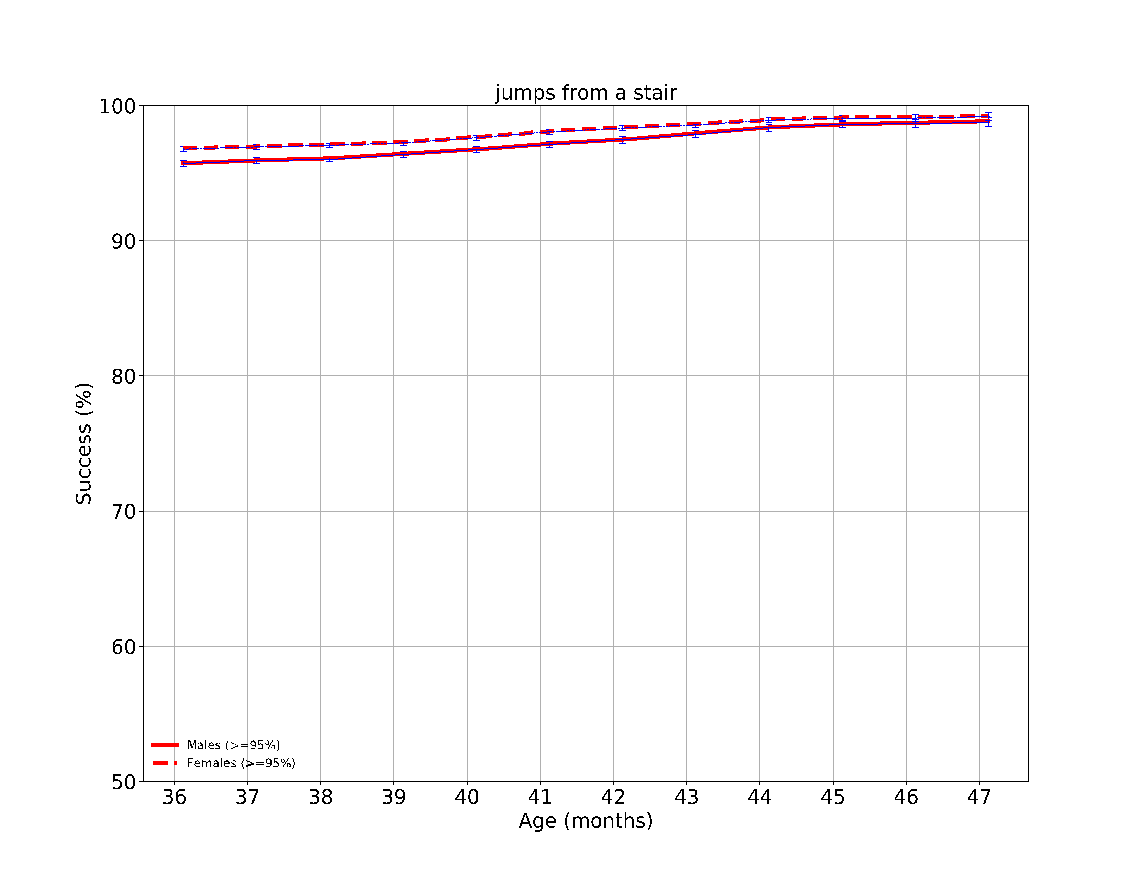

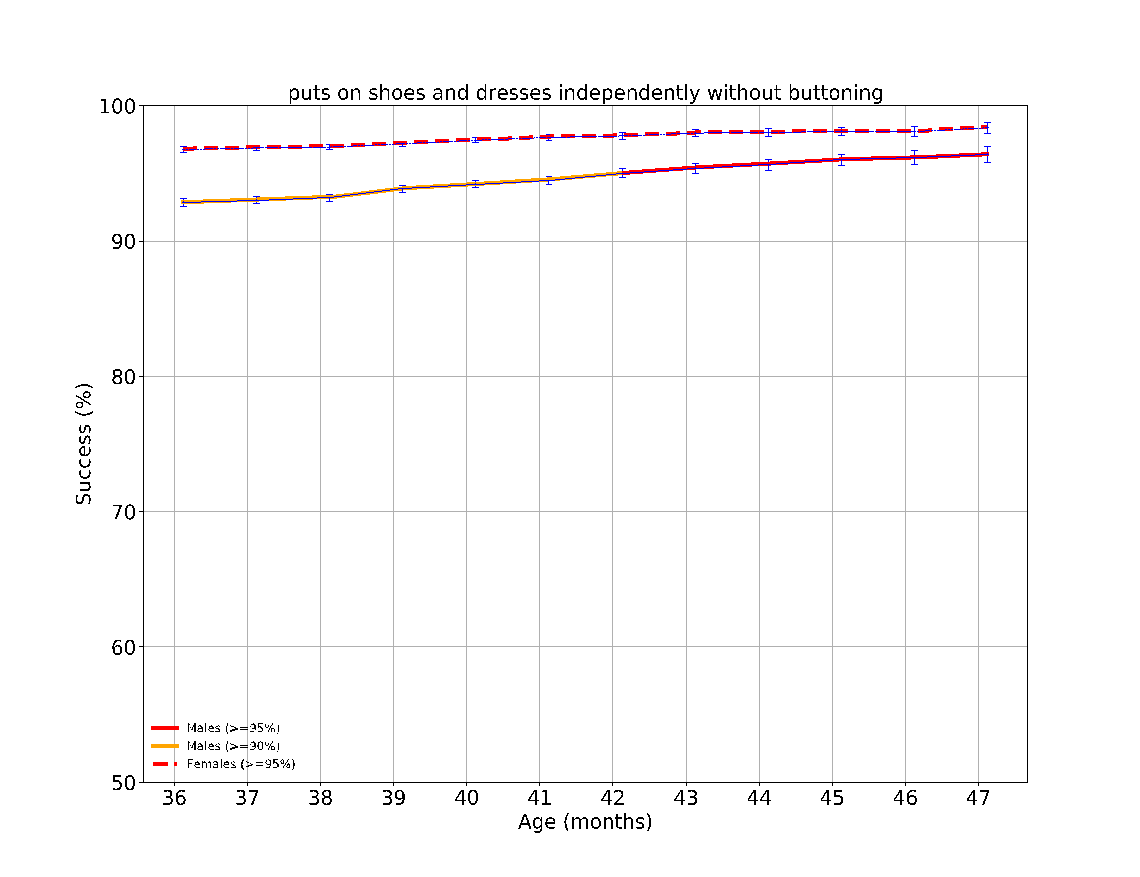

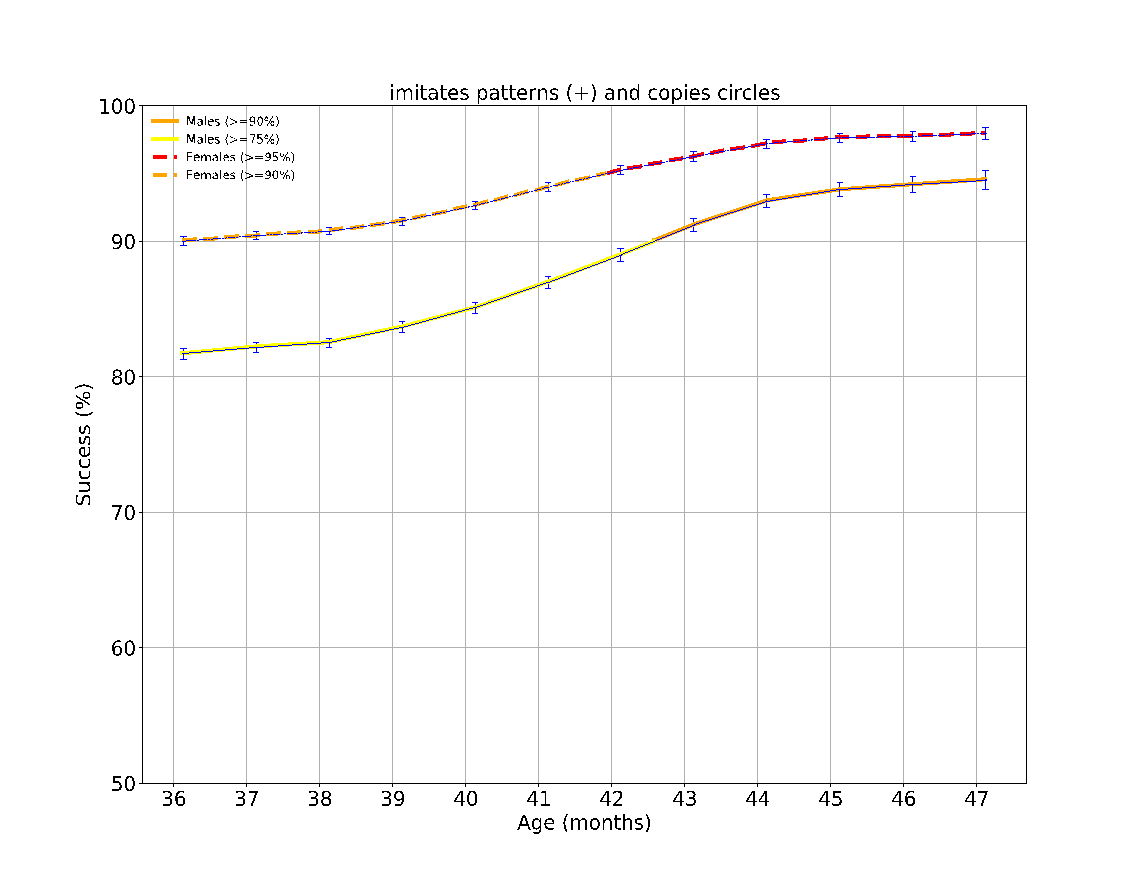

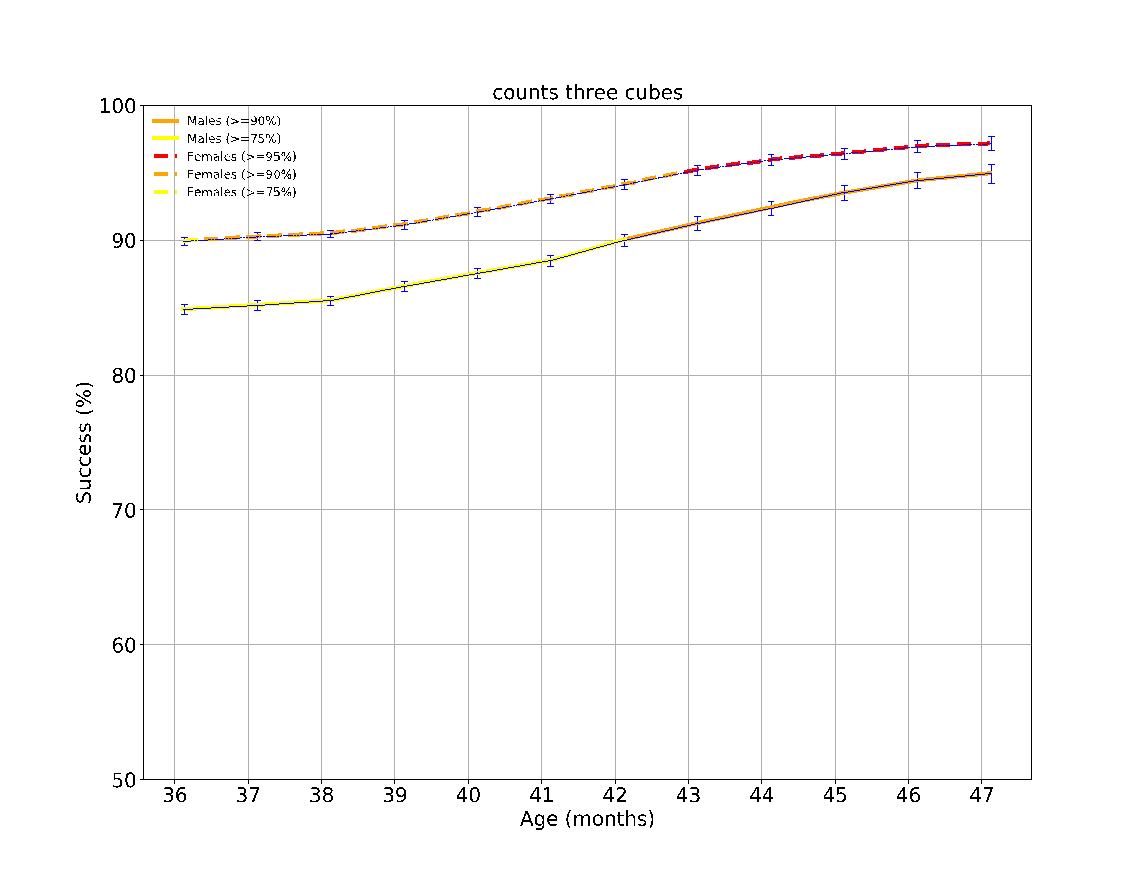

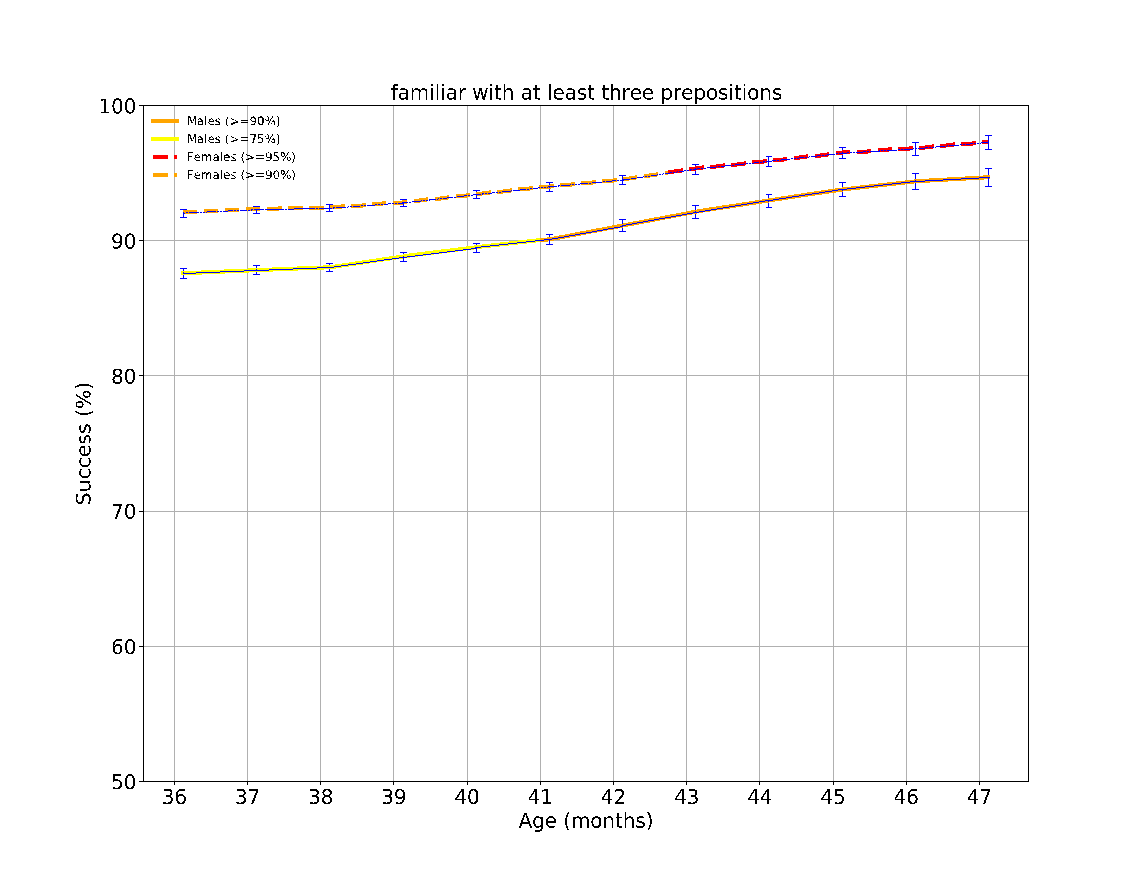

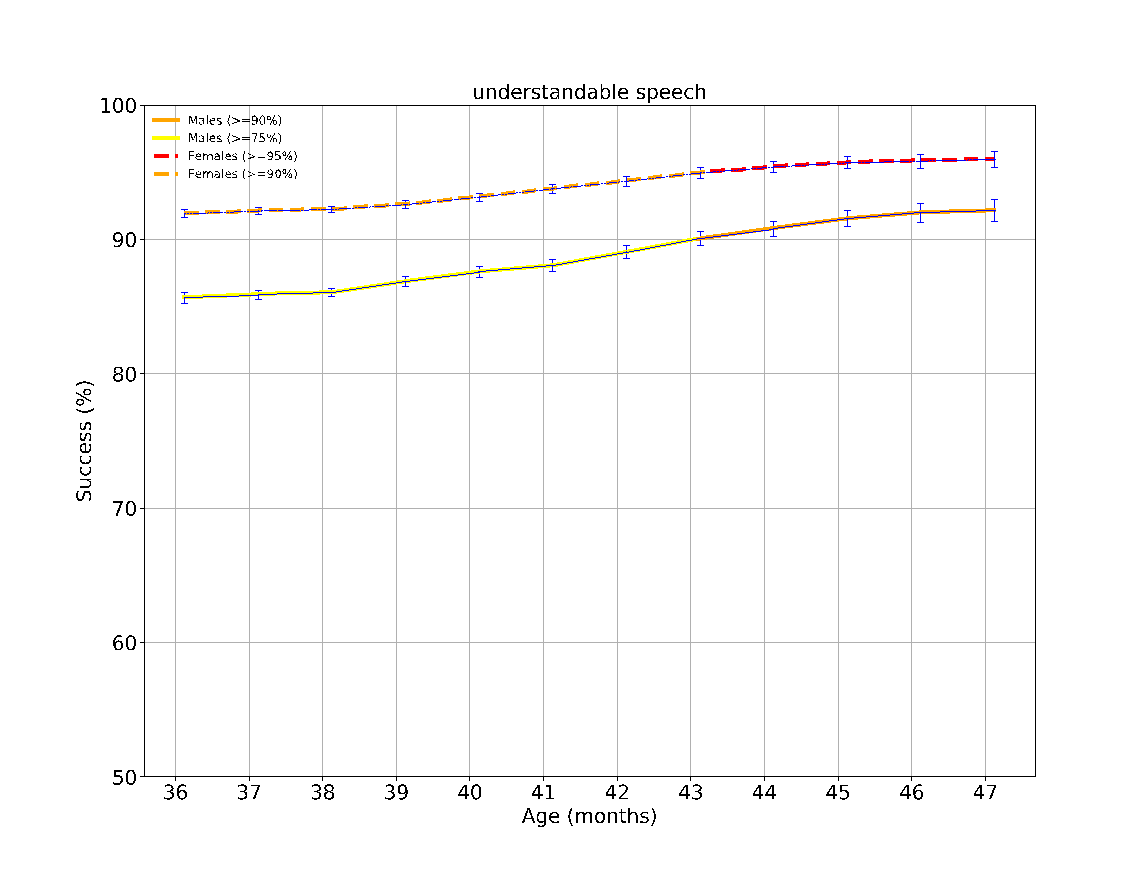

eFigure 9: The success rates over time of the different milestones evaluated between the ages 36-48 months. The achievement rates of < 75%, 75-90%, 90-95% and > 95% of children are indicated by the colors green, yellow, orange and red, respectively. The error bars are 95% confidence intervals of a binomial proportion, where larger bars indicate smaller sample size.

**eFigure 10 - Task performance results over time according to sex at the age 4-6 years (48-72 months)**

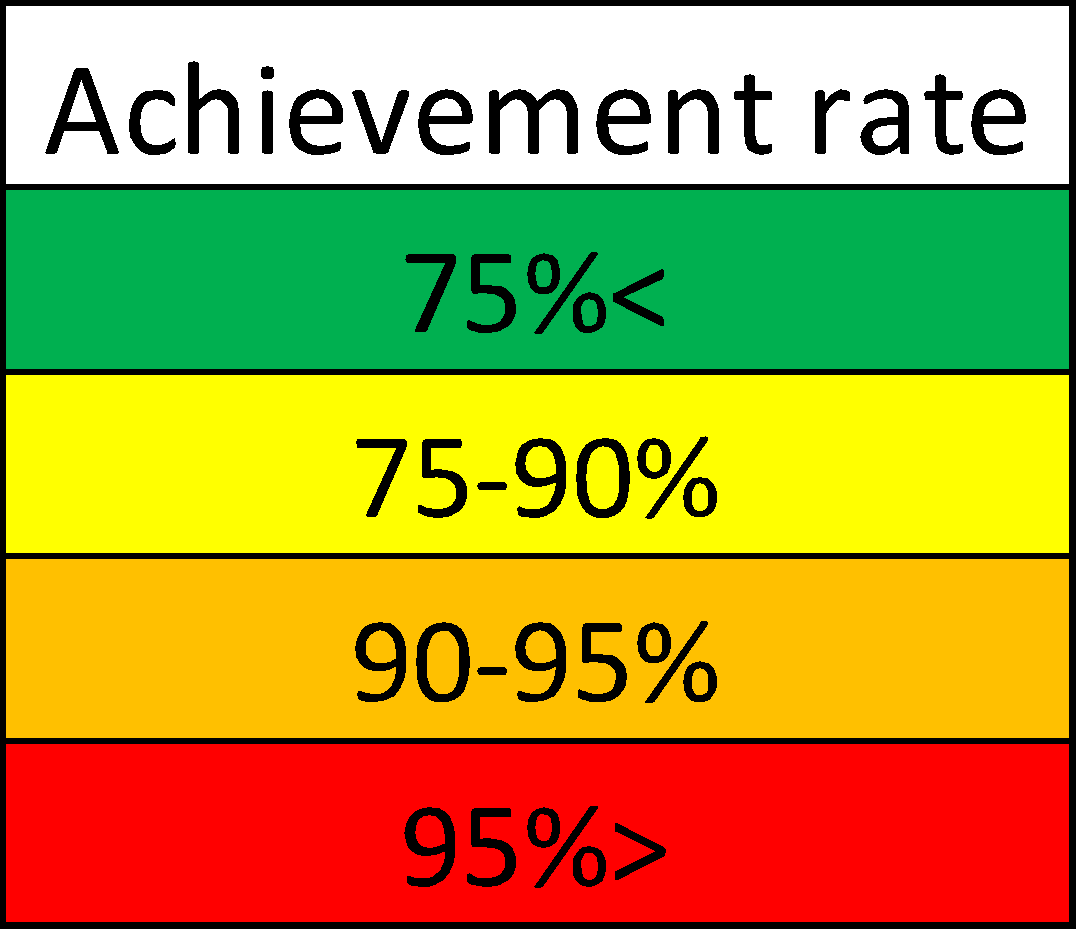

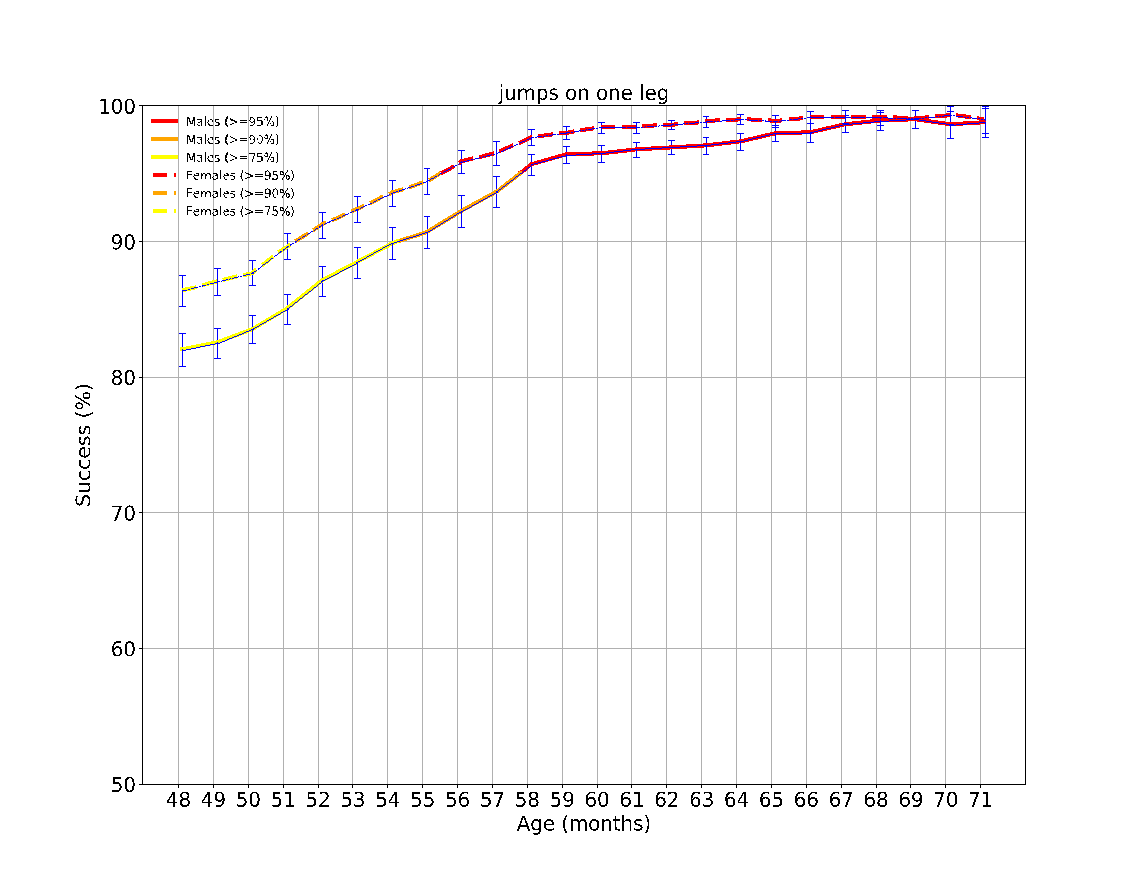

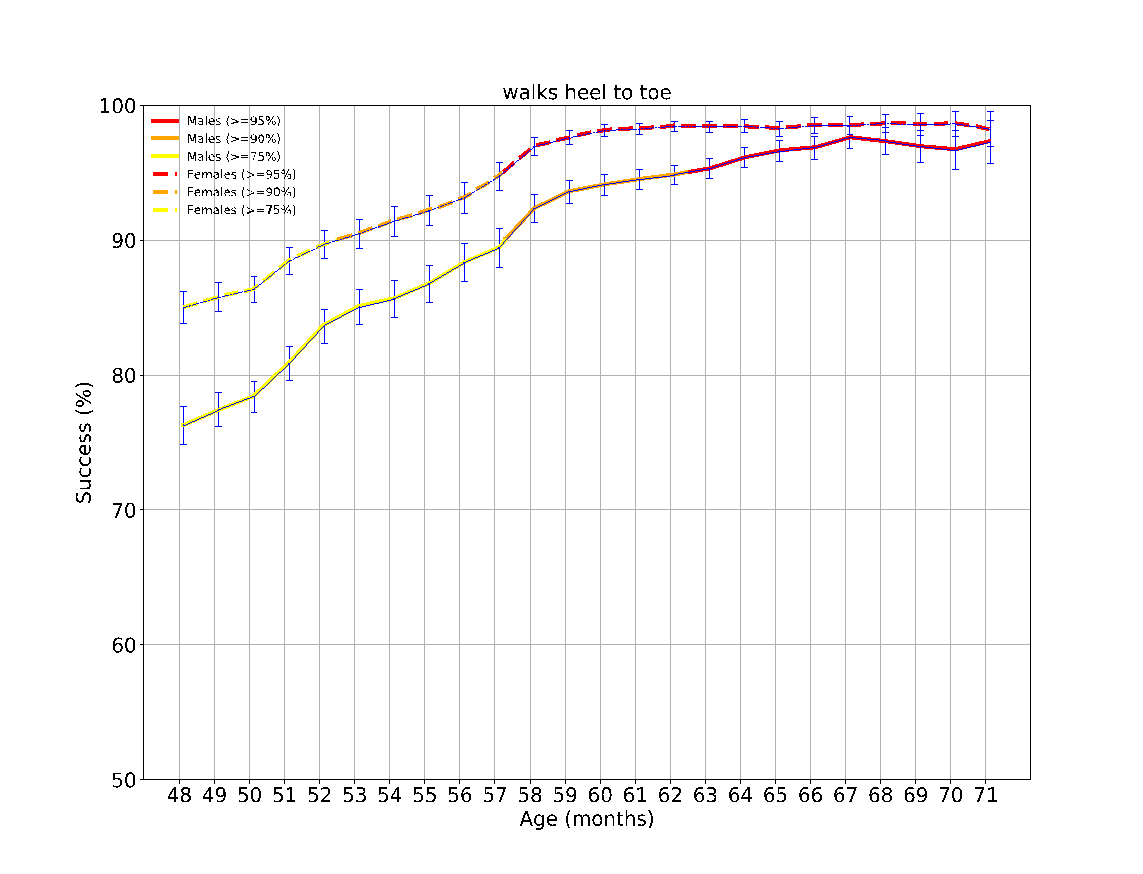

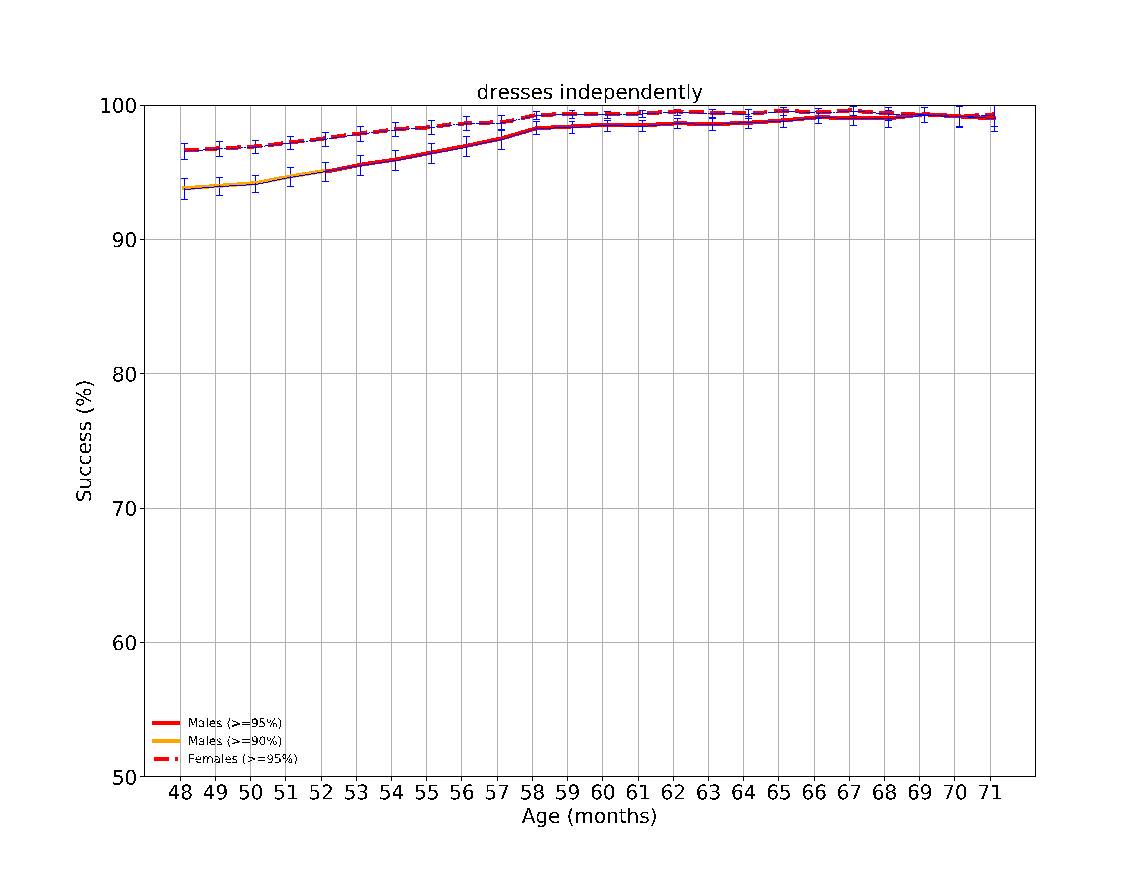

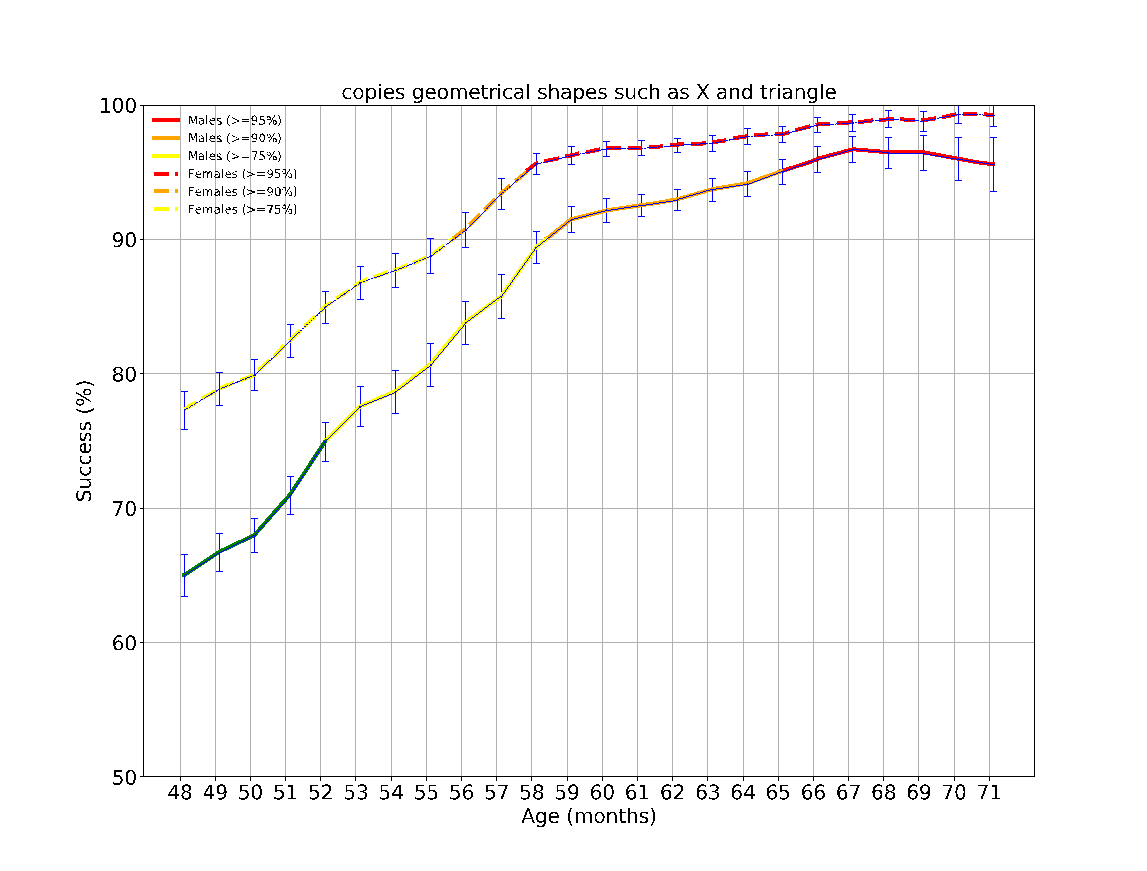

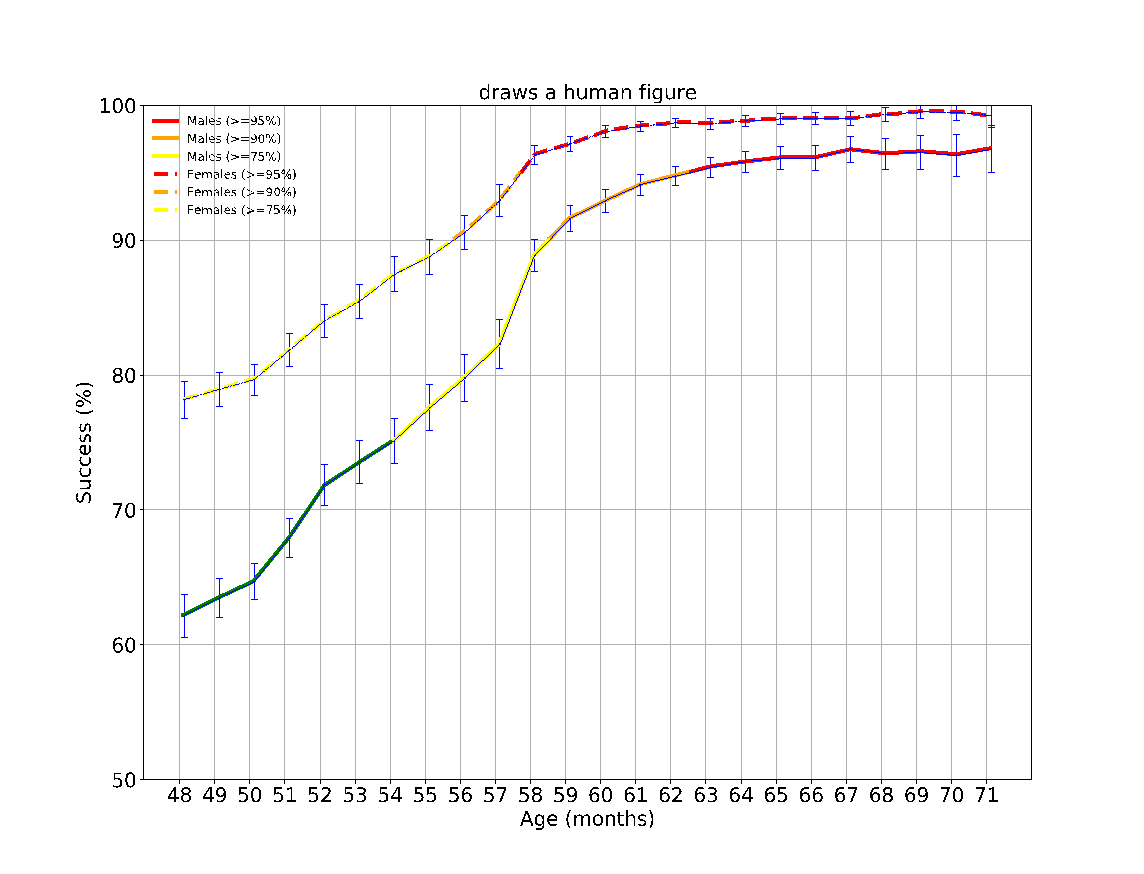

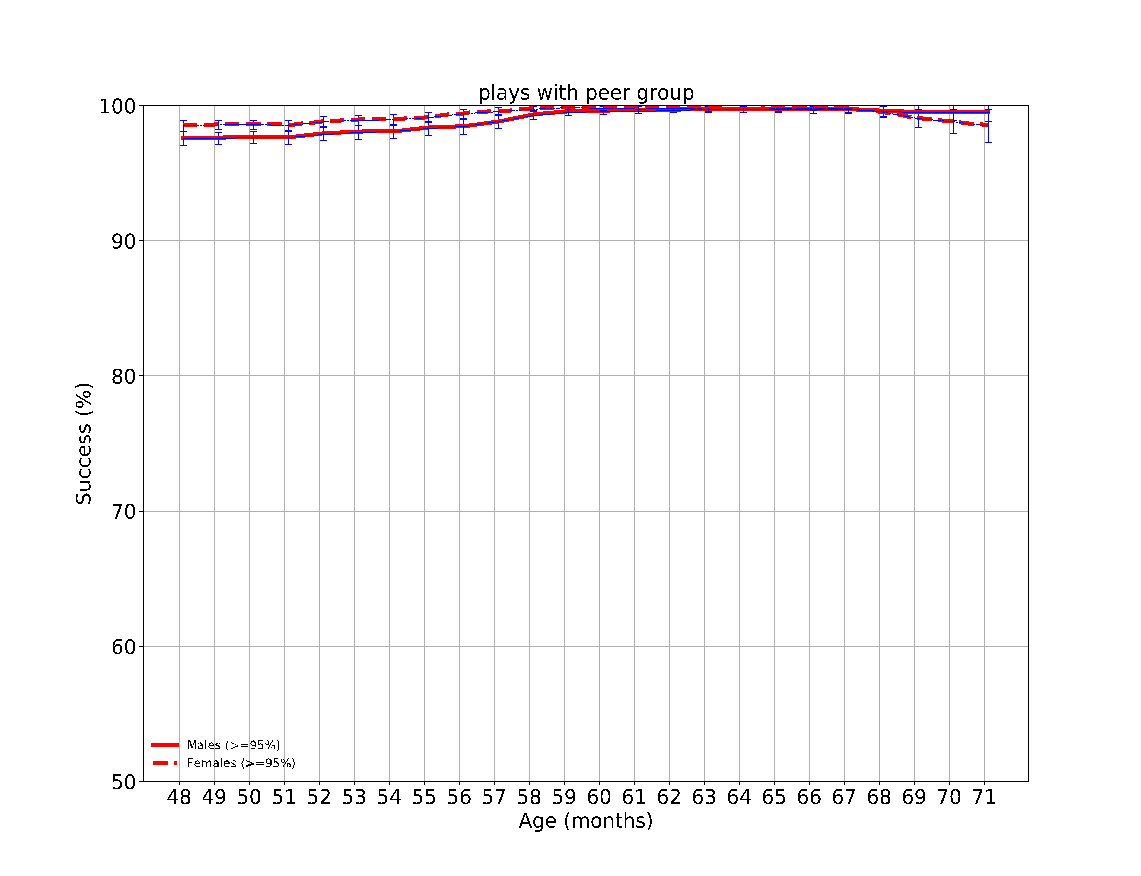

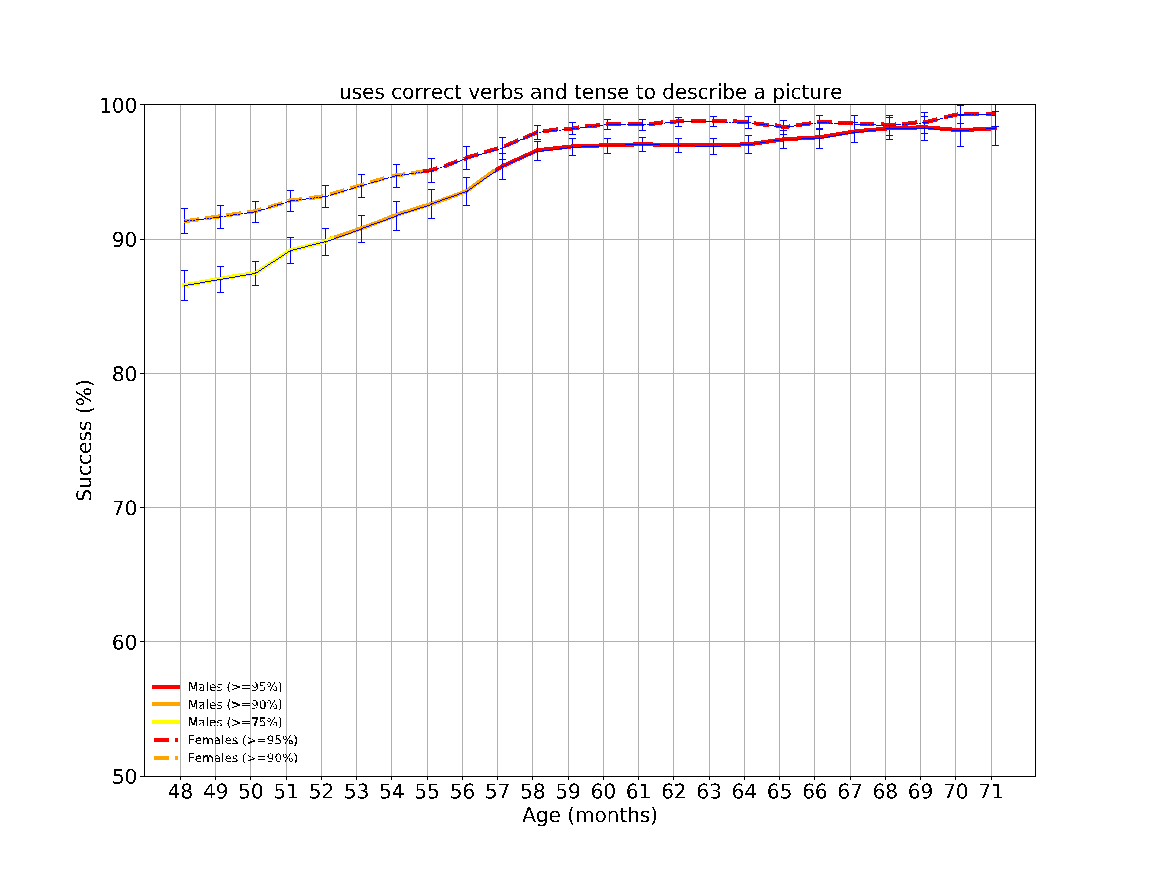

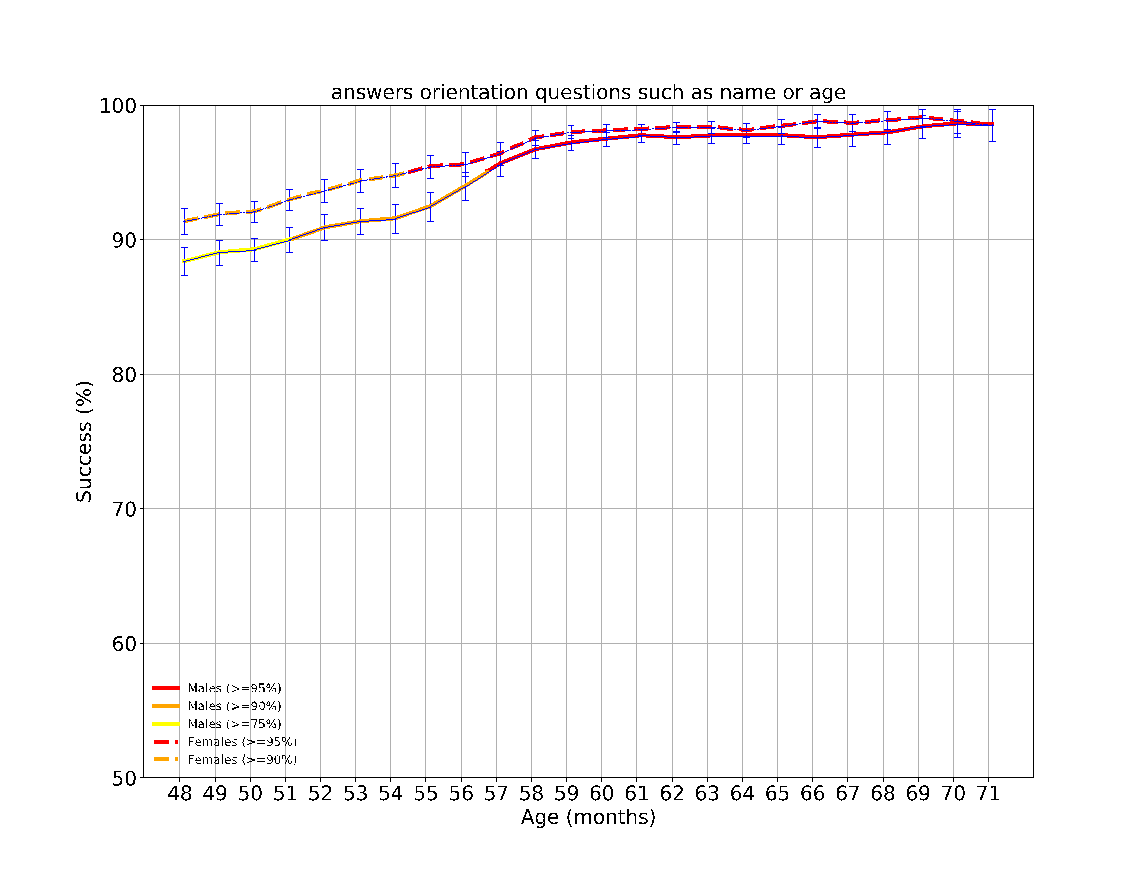

eFigure 10: The success rates over time of the different milestones evaluated between the ages 48-72 months. The achievement rates of < 75%, 75-90%, 90-95% and > 95% of children are indicated by the colors green, yellow, orange and red, respectively. The error bars are 95% confidence intervals of a binomial proportion, where larger bars indicate smaller sample size.

**eFigure 11 – Odds ratio for unattained developmental milestone**

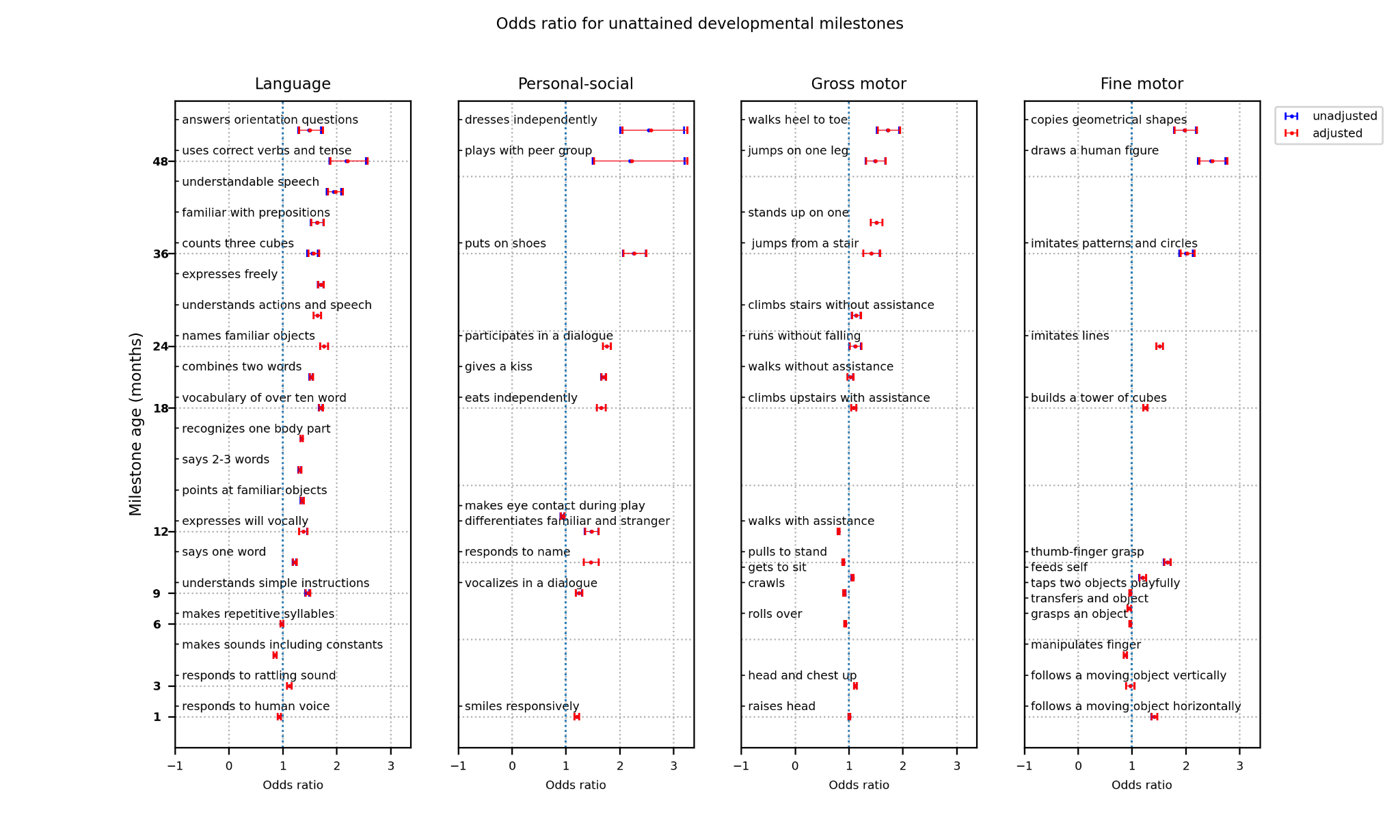

eFigure 11: The odds ratio of failing to achieve a milestone by boys, while controlling for the other predictor variables, including: the child’s sex, birth weight, gestatoinal age at birth, one-minute Apgar score, and the mother’s ethnicity, country of birth, education level, marital status, and age group. For example, boys are at a 2.63 times increased risk for not achieving the milestone dresses independently (CI 2.08-3.33).

**eFigure 12 – Comparison of milestones attainment rate between sexes of ‘successful’ children at the age of two years**

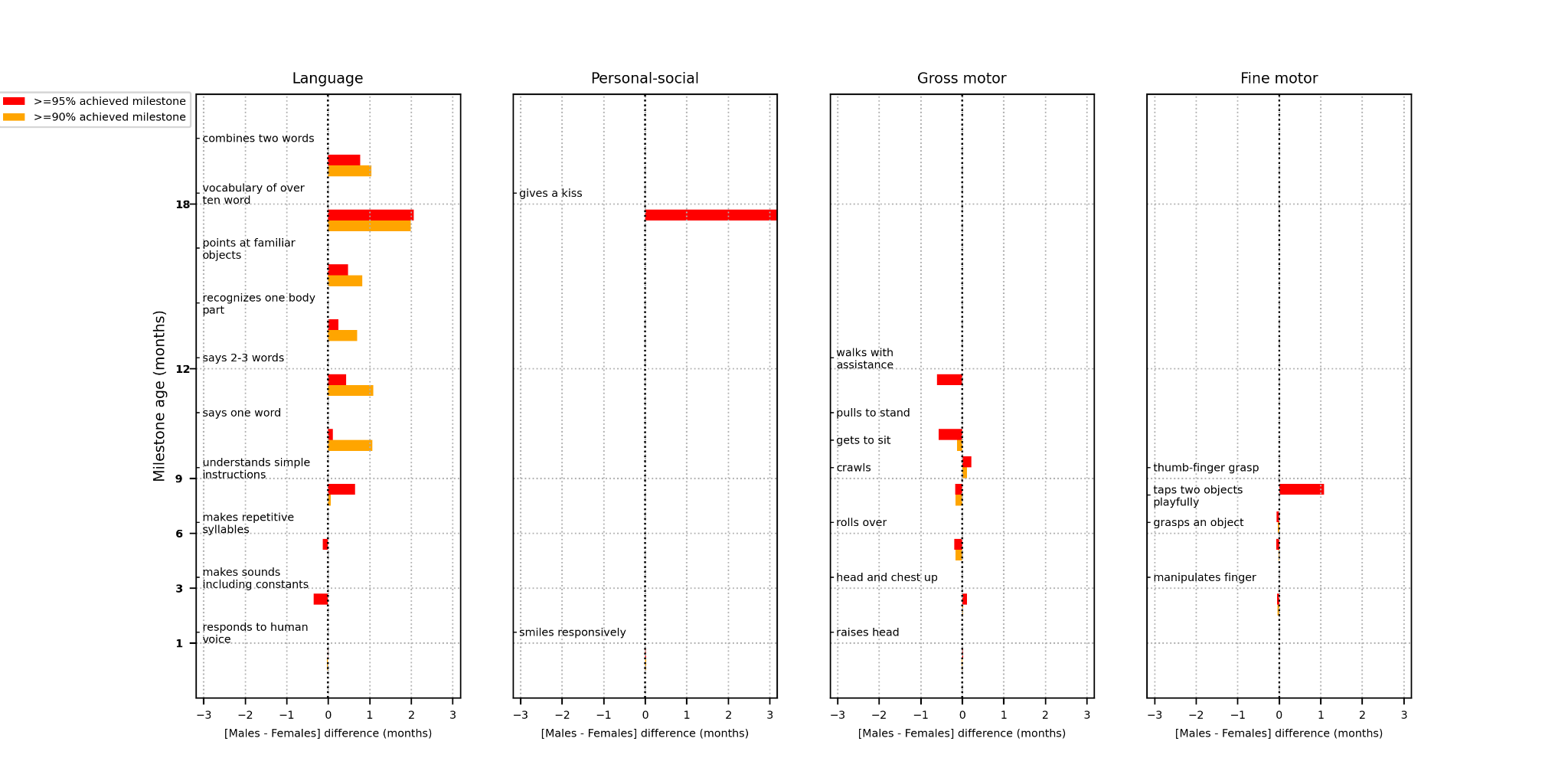

eFigure 12: Analysis of early milestones attainment rate of both sexes, among a subset of ‘successful’ children who attained all milestones at the age of two years.

**eTable 1 - Israeli Ministry of Health - A guide to performing developmental assessments for infants and toddlers from birth up to the age of six years**

| milestone | Method of evaluation | pass |
| --- | --- | --- |
| Age 6 weeks to 3 months | | |
|  | The baby lies on a flat surface, secure |  |
| Visually follows a moving object horizontally | While lying on the back, the baby focuses his gaze on an object such as a colorful ring or a ball, the face of the parent or the examiner.   The object, the face of the examiner or the parents should be within 20 to 30 centimetres from the baby’s face.   Place the object or face in front of the baby's face and move in slow motion approximately 45 degrees to one side, back to the center, and then to the other side.   The test should not be accompanied by sounds or voices. | The baby visually follows a moving object 45 degrees horizontally both directions |
| Vocalizes in response to human voice | Establishing eye contact with the baby is it preliminary condition to the accomplishment of this task.   it is recommended to guide the parent to make speech sounds to the baby, while providing a recess, to allow the baby to listen and respond. | The baby makes sounds of joy or vocalize differently according to different needs. |
| Smiles responsively | Establishing eye contact with the baby is a preliminary condition to the accomplishment of this task.   The child's response to a stimulus sound or human face during the interaction should be examined.   To evoke a smile, the examiner can bring his face closer to the baby with a wide smile and speak softly. | The baby smiles in response to human face or human voice or stimuli. |
| Raises head | The baby lies on his abdomen. To see spontaneous movements, the examiner should refrain from placing his hand on the baby’s back or the pelvis. | The baby lifts his head off the surface momentarily and turn to either side. |
| Age 3-6 months | | |
| Visually follows a moving object vertically | The baby follows a moving object vertically: up or down, within the middle line in a distance of 20 centimeters from his face | The baby follows a moving object vertically. |
| Responds to rattling sound | The baby lies down or is in his parents’ lap. A noise is produced by a rattle (a noisemaker, a knocking noise) in a distance of 50 centimeters from both sides of the baby's head and outside his vision range.   The noise production will be made in the same plane as the baby's ears and not higher.   Please note that there are no background noises during the evaluation. | The baby responds to the rattling sound in some manner, including: cessation of activity, eye blinking, head turning towards the sound origin. |
| Responds to human presence | When the baby is relaxed, a response to a human figure, touch or voice is examined. | The baby smiles spontaneously at a human figure, touch or human voice, or if when playing alone, stops crying when spoken to and sometimes mimics facial expressions. |
| makes various sounds including constants (I.e. Mm rr gg) | Various voices including constants such as ahh or goo.  During the entire visit attention should be paid to the production of these various voices.  The baby plays “vocally” by himself or when played with makes noises when he is excited or dissatisfied. | A variety of sounds are heard including consonants, such as – gg, grr, brr, mm, pp and etc. |
| Hands together, manipulates fingers | The action is observed while the baby lies on his back.   The disappearance of the initial congenital reflexes allows the baby to play with his hands in front of the center axis, in the midline. | The baby brings his hands to the midline, to his mouth or combines fingers.    Attention must be paid to the symmetry in hands movements to the midline while lying on the back. |
| grasps an object | During the examination the baby lies on his back. The task varies mildly according to the age in which the child arrives to the evaluation. | At the age 3-4 months the baby grasps an object that was placed in his palm, in each palm separately.   At the age of 4-5 months the baby reaches his hand to an object served to him at the middle line or placed on the chest and grasps with both hands.   At the age of 5-6 months the baby reaches his hand to objects that are within reachable distance, grabs them and approximates them to his body or looks at them. |
| Head and chest up in prone position | The baby lies on the abdomen, the examiner should refrain from intervening by placing hand on the baby’s back or pelvis. The baby's movements are directed towards achieving a goal, such as a directed look at an object or figure. His curiosity in exploring the environment should also be noted. | The baby lifts his head and chest while leaning on the forearms. |
| Age 6-9 months | | |
| transfers an object from one hand to the other | The baby is lying on the back or sitting on the parent’s lap, the examiner in front of him presents him with a toy in the middle line. | The baby reaches out one hand to the toy, grabs it and passes it to the other.   Watch the transfer in both directions and pay attention to the symmetry in the hand movements. |
| makes repetitive syllables-constant or vowels | Voice productions should be listened to throughout the visit.   During the "conversation" with the baby give time to allow him to listen and respond. | The baby makes mumbling sounds of syllables. The pronunciations are similar to the consonants and vowels of the language to which the baby is exposed, for example – mamha, dadha. |
| rolls over from abdomen to back and back to abdomen | please notice that the roll over is from the right side to the left and from the left side to the right.   The examiner can interest the baby with a toy placed within an angle of 45 degrees above the head. | The baby rolls over from the back to the abdomen and from the abdomen to the back. |
| Age 9-12 months | | |
| taps two objects playfully | The task should be performed when the baby lies on the back or sits on the parent’s lap. The examiner presents 2 toys or cubes, one for each hand. | The baby holds an object (cube) in each hand and taps between them.  The baby enjoys the sound and will continue to do so with pleasure.   Note symmetry in hands movements. |
| crawls | The child lies on his abdomen. The examiner should try to encourage the baby to move forward in crawling.  Observing the style of crawling and the symmetry of activation of both sides of the body alternately. | The baby is observed crawling. All or part of the crawling styles are possible. |
| vocalizes in a dialogue | Voice productions should be listened to throughout the visit. During the "conversation" with the baby give a break to allow him to listen and respond. | The baby initiates and produces double syllables in a dialogue such as "aa-dada", parts of words, or alternatively makes sounds of animals. |
| responds when addressed by name | The mother or the nurse refer to the child by name (the name in which the parents are accustomed of using) while being relaxed and in a soothing environment. The baby must be addressed once and then response should be waited for. this could be repeated. | the baby responds when addressed by name. Responses can be various including cessation of activity, referral of the head or look, vocal response |
| responds differently to familiar and stranger | During the visit, the child's reactions to the familiar parent and to the examiner whom the child does not know should be monitored and evaluated. | There is a different reaction to the familiar parent, opposed to a certain reluctance to the unfamiliar character - the child will check, examine, or cry as a reaction to the examiner. |
| feeds self | Serve the child food to hold in his hand or a bottle | The baby feeds himself hand-held food, such as a cookie or a cooked vegetable.  He Drinks from a bottle independently and from a cup with assistance. |
| uses thumb- fingers grasp | A small pea-sized object (such as a raisin or cereal) is served to the baby within a reachable distance, under observation.  Each hand should be evaluated separately. The child should be prevented from bringing the object to the mouth. Melted cereals should be used. | The child holds a small object in his thumb and 2 fingers. Later the child is able to grasp the object at the tip of the fingers between finger and thumb in a pinching motion. Symmetry should be monitored. |
| understands simple instructions | Understanding instructions is evaluated by looking or pointing, such as: "Where is the light?", "Where is the bottle / cup?", "Applause", "wave hello", etc. without guidance or demonstration. Ask the toddler to point or look at a familiar object in the room. | The baby follows three simple instructions, one of them must refer to a human figure, such as “where is mom\dad?”. |
| gets to sit without support | The child lies on his stomach or back. The child sits by himself from various positions. | The baby moves from lying to sitting position. Sitting can be in a variety of poses. |
| Age 12-18 months | | |
| says one word or pronounces meaningful sounds | Encourage the baby to converse while giving time to allow him to listen and respond.  It is important to listen to the baby during the entire visit. | The child says one word or pronounces meaningful sounds |
| expresses will vocally or with gestures | During the visit, the child's reactions to the familiar parent and to the examiner whom the child does not know should be monitored and evaluated.  The toddler can express desires in words, in a vote or by gesture. The toddler initiates. | The child initiates the expression of desires vocally - uses words or parts of words. By voting - for example on a bottle - "drink", or a door - "walk"; Or in a gesture - for example, raises his hands to be picked up. |
| makes eye contact and expresses reciprocity during joint game | The toddler plays with the examiner or parent with a ball, car, dice, doll - according to his preferences. | The child focuses his gaze and follows both the object and the parent or examiner during the game.  There is reciprocity - the child reacts to the other and shares the other while playing. |
| walks with assistance | The parent or examiner encourages walking by offering an interesting game at a suitable height and at a reasonable distance, when the child is assisted by furniture support (a stable chair, a stable stroller, etc.). | The toddler walks with support, moves between one furniture to the other, around walls and uses them for mobility, and / or with the help of an adult’s hand. |
| points at familiar objects to request | The toddler is asked to point to a familiar object in the room. For example: show me where is the ball. | The toddler understands and executes simple instructions with gestures, like pointing on an object upon request. |
| pulls to stand | The toddler should be encouraged to pull himself to a standing position with the help of a chair or a low table, without an adult’s assistance. | The toddler manages to rise from the floor with support in various manners, and sets himself up to a standing position. |
| says 2-3 words | Voice productions should be listened to throughout the visit. | The toddler consistently and regularly says 2-3 words including "dad" "mom" or pronounces consistent expressions understood to the environment and the family, such as: "Nana" for banana, "How" for dog.   In bilingual families, all words in each language must be referred to (for example, ball in both languages - is considered 2 words). |
| Age 18-24 months | | |
| climbs upstairs with assistance | The child should be encouraged to climb stairs with a handrail that he can hold under supervision. the parent or examiner may also hold his hand in support. | The toddler goes up and down the stairs with the assistance of a helping hand or a handrail. |
| walks without assistance | The parent or examiner encourages walking by offering an interesting game at a suitable height and at a reasonable distance. | The toddler walks without assistance. |
| eats independently with a spoon | Information from the parent or primary caregiver should be provided regarding the manner of eating, the times and types of foods and textures; Toddler integration at family meals and experience of new foods. | The toddler eats independently with a spoon. |
| familiar with at least one body part | The toddler will be asked to show one or more body parts upon request and can show this to the parent or doll. | The toddler recognizes at least one body part. |
| builds a tower of cubes | While sitting at a small table or on a carpet, the parent or examiner presents cubes to the toddler and asks him to build a tower. If the toddler does not perform by request, the parent or examiner will demonstrate building a tower. | At the age of 18 months - The toddler builds a tower from 3 cubes or more.  At the age of 21 months - The toddler builds a tower from5 cubes or more. Towards the age of two years - he builds a tower from 6 cubes or more. It should be noted that the toddler uses both hands during the tower building. |
| has a vocabulary of over ten words | The conversation between the parent and the toddler should be listened to at all times during the visit.  It is recommended to show the toddler a picture book and encourage him to talk, it is possible to use the parent’s assistance. | The toddler has a vocabulary of over ten words. You can get a report from the parent on the number of words or expressions in which the toddler uses. |
| squeezes and sticks out lips to give a kiss | The parent or examiner ask the toddler to kiss the parent or a doll. | The toddler squeezes and sticks out lips to give a kiss. |
| Age 2-3 years | | |
| composes a sentence of at least two words | The conversation between the parent and the toddler should be listened to at all times during the visit.  It is recommended to show the toddler a picture book and encourage him to talk, it is possible to use the parent’s assistance.  It is possible to get a report from the parent regarding sentences in which the toddler uses. | The toddler succesfully combines two words into a sentence, for example - father-come, mother-water, want-water.  It is possible to get a report from the parent regarding sentences in which the toddler uses. |
| runs well without falling | If possible, performed during the visit at the station. The examiner should pay attention to the child's organization before performing the task. Multiple falls requires attention and reference. | The toddler runs freely and confidently, stepping on each foot fully, while paying attention to obstacles in his path and his ability to bypass them without dropping them. |
| climbs up and down the stairs without an adult's assistance | If possible, performed during the visit at the station. The child should be encouraged to go up and down the stairs with a railing that he can hold under supervision. Do not hold his hand in support. | The toddler goes up and down the stairs with the help of a railing, without the support of an adult. |
| recognizes familiar objects and pronounces them by name | The toddler is asked to name a variety of items or pictures that the examiner points to in the room while asking "What is it? What is it called?" | The toddler will name a number of items from the familiar environment, such as: clothing, furniture, games, fruits, animals and more.  Mispronunciations are acceptable. |
| understands actions and speech without gestures | The toddler is required to understand and perform actions asked by the examiner, literally without marking with a gesture or hint, with objects from the assessment kit. For example: Show me what do we drink with, or give me the cup. | The toddler understands at least 3 actions such as: give, show, put. Understands at least one preposition such as - on, within. |
| participates in a dialogue | It is advisable to listen during the entire visit to the conversation that develops between the mother and the toddler. It should be monitored and seen whether a dialogue develops with the adults during the evaluation, whether the child expresses feelings and desires appropriately. Does he ask situation-appropriate questions like: who? what? where?. | The toddler initiates a verbal dialogue with the examiner or parent in a suitable situation, expresses his desires, responds to his interlocutor's response and is able to share experiences from everyday life. Please note: Reverberating words without meaning or intention of communication will not be considered as speech aimed at communication purposes, and requires further evaluation. |
| participates in daily activities | Interview with the parent regarding: dressing, eating, bathing, urine and bladder control, playing.   A question about diaper weaning should be asked with great sensitivity according to the nurse’s acquaintance with the family, its customs and culture, and following the proper previous guidance. |  |
| imitates horizontal, vertical and circle lines | The toddler sits on a chair at a small table next to the parent. The examiner or the parent hands the child a sheet of paper and a pencil, shows him how to draw horizontal, vertical and circle lines. | The toddler imitates horizontal, vertical and circle lines. |
| Age 3-4 years | | |
| expresses freely | The examiner can use individual pictures or a book (descriptions of the child's daily life), and ask open-ended questions: what's going on here? or tell me or mom a story about what is happening in the picture - to allow the toddler to answer in a complete sentence.  The toddler is able to describe a number of pictures or different situations performed by the nurse. | The toddler's ability to describe pictures and combine words is evaluated.  The child performs successfully if there are two-word combinations, one of which is a verb, such as: child traveling, drinking milk, and also combinations such as: want to eat, or two-words combinations (which are not verbs), which are connected by prepositional letters such as: on, in. For example: a doll in a stroller, a bear in bed, a child on a bicycle, a doll in a chair. |
| jumps from a stair | The child is asked to jump off a step, independently - without support. Up to 3 attempts should be allowed.  The action can be demonstrated. | The toddler jumps from a step and lands on both feet, independently - without support. |
| puts on shoes and dresses independently without buttoning | The examiner should ask if the child dresses by himself, independently. | The child dresses himself but needs help with buttoning and tying laces. Wearing only one item of clothing and shoes is sufficient.   The child should not be expected to differentiate between a right shoe and a left shoe. |
| imitates patterns (+) and copies circles | The child sits on a chair at a small table next to the parent. The examiner or the parent hands the child a sheet of paper and a pencil, the child is asked to draw a + pattern and an O pattern. | The child successfully draws a + or O pattern. |
| Age 4-5 years | | |
| stands up on one leg for 3 seconds | The child is asked to stand on one leg. Standing should be on one leg without support when the raised leg is free in the air. You can repeat the action up to 3 times. Initially the child will choose for himself on which leg to stand and then he should be encouraged to stand on the other leg.  Differences between the quality of action on one leg compared to the other leg are expected. | The child stands up on one leg for 3-4 seconds. |
| counts three cubes | The examiner places cubes in front of the toddler and asks him to count them. Going forward the examiner extends his palm and asks to put 3 cubes in it. After the toddler has completed the task, waiting, and counting along with him the number of cubes he placed in the palm of the examiners hand. The toddler should associate each number with a cube. | The child counts at least three cubes. |
| familiar with at least three prepositions | It is advisable to place an object in a certain place so that the toddler will be able to understand and use the appropriate prepositions accordingly. For example: put a cube underneath the table. You can use pictures / book. | The child is familiar with at least 3 prepositions such as: on top, next to, underneath and more. |
| plays with peer group | Parental report regarding playing with peer group. | The child plays in a joint game with other children, initiates and develops imagination and imitation games. Is able to concentrate on board games. Familiar with simple rules of the game. |
| dresses independently | When preparing for a weight and height test check if the toddler is capable to undress and dress by himself. The ability to fasten buttons and zipper can be evaluated with the appropriate item in the development kit. Demonstration is possible if necessary. | The child is able to undress and dress himself, including button fastening. |
| understandable speech | During the visit at the station, listen to the developing conversation between the parent and the toddler: the toddler is able to tell a story, repeat a recitation / song, knows his name, uses male / female correctly. | The speech is clear and understandable to everyone. |
| Age 5-6 years | | |
| uses correct verbs and tense to describe a picture | A book or pictures (including personal and family pictures) can be used to encourage the toddler to tell a story around the picture, or describing a simple story in sequence from pictures. In addition, you can evaluate the language skills during a free conversation with the toddler, from listening to him during the visit. | The child describes a picture using correst verbs and tenses. |
| answers orientation questions such as name or age | Ask the child to say his full name, age and address. | The chils says his full name, age and address. |
| jumps on one leg | The child is required to jump 2-3 times on one leg in a row, out of 2-3 attempts. Demonstration is possible if necessary. An attempt to jump on the other leg should be evaluated, although better control on one leg is expected. | The child is required to jump 2-3 times on one leg in a row, out of 2-3 attempts. |
| walks heel to toe | The child is asked to walk in a straight-line heel to toe (thumb side heel), 4-7 steps in 2 out of 3 attempts.  Demonstration is possible. | The child walks in a straight-line heel to toe (thumb side heel), 4-7 steps in 2 out of 3 attempts. |
| draws a human figure | The child sits on a chair at a small table next to the parent. The examiner or the parent hands the child a sheet of paper and a pencil, the child is asked to draw a human figure. Do not guide the child while performing the task to notice and complete missing parts (organs) in the drawing. | The child draws a human figure with at least 6 clear and understandable organs. |
| copies geometrical shapes such as X and triangle | The child sits on a chair at a small table next to the parent. The examiner or the parent hands the child a sheet of paper and a pencil. The examiner or parent shows the child a drawing of a X and triangle and asks the child to copy the shapes. | The child copies the X or triangle. |

eTable 1: The Israeli Ministry of Health protocol for developmental evaluation of infants and toddlers from birth up to the age of six years. The protocol describes the evaluated milestones for each age group, the method of evaluation and the definition of which behaviour qualifies as success.

**eTable 2: A comparison of the attainment age of each milestone by 90% and 95% of the children according to sex, alongside the time difference between groups**

| **Milestone** | **Domain** | **Boys 95% age (m)** | **Boys 90% age (m)** | **Girls  95% age (m)** | **Girls  90% age (m)** | **Unified scale 95% age (m)** | **Unified scale 90% age (m)** | **Boys-Girls 95% age difference (m)** | **P-value for different attainment rates by 95% of boys/girls** | **Boys-Girls 90% age difference (m)** | **P-value for different attainment rates by 90% of boys/girls** | **Unified scale - Boys 95% age difference (m)** | **Unified scale - Boys 90% age difference (m)** | **Unified scale - Girls 95% age difference (m)** | **Unified scale - Girls  90% age difference (m)** |
| --- | --- | --- | --- | --- | --- | --- | --- | --- | --- | --- | --- | --- | --- | --- | --- |
|  |  | 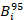 | 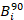 | 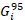 | 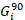 |  |  | 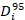 |  | 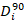 |  |  |  |  |  |
| visually follows a moving object horizontally | fine motor | 0.35 |  | 0.35 |  | 0.35 |  | 0.00 |  |  |  | 0.00 |  | 0.00 |  |
| vocalizes in response to human voice | language | 1.58 | 1.37 | 1.61 | 1.38 | 1.59 | 1.37 | -0.03 | 2.71E-04 | -0.01 | 8.96E-05 | 0.02 | 0.00 | -0.01 | -0.01 |
| smiles responsively | personal-social | 1.63 | 1.42 | 1.58 | 1.40 | 1.61 | 1.41 | 0.06 | 7.24E-24 | 0.02 | 2.18E-21 | -0.03 | -0.01 | 0.03 | 0.01 |
| raises head | gross motor | 3.50 | 2.76 | 3.46 | 2.71 | 3.48 | 2.74 | 0.04 | 7.39E-01 | 0.05 | 9.78E-04 | -0.02 | -0.02 | 0.02 | 0.02 |
| hands together, manipulates fingers | fine motor | 3.78 | 3.51 | 3.82 | 3.53 | 3.80 | 3.52 | -0.04 | 7.95E-10 | -0.02 | 5.31E-05 | 0.02 | 0.01 | -0.02 | -0.01 |
| grasps an object | fine motor | 5.11 | 4.05 | 5.16 | 4.08 | 5.13 | 4.07 | -0.05 | 1.27E-02 | -0.03 | 9.84E-03 | 0.02 | 0.02 | -0.03 | -0.01 |
| visually follows a moving object vertically | fine motor | 3.13 |  | 3.13 |  | 3.13 |  | 0.00 |  |  |  | 0.00 |  | 0.00 |  |
| Responds to rattling sound | language | 3.14 | 3.13 | 3.13 |  | 3.13 |  | 0.01 | 4.12E-02 |  |  | -0.01 |  | 0.00 |  |
| makes various sounds including constants (ah-goo) | language | 3.55 | 3.13 | 3.62 | 3.13 | 3.59 | 3.13 | -0.07 | 2.30E-12 | 0.00 |  | 0.04 | 0.00 | -0.03 | 0.00 |
| head and chest up in prone position | gross motor | 4.66 | 3.63 | 4.20 | 3.59 | 4.40 | 3.61 | 0.46 | 7.45E-26 | 0.04 | 9.55E-17 | -0.26 | -0.02 | 0.20 | 0.02 |
| rolls over from abdomen to back and back to abdomen | gross motor | 8.53 | 7.55 | 8.72 | 7.65 | 8.61 | 7.60 | -0.19 | 4.22E-03 | -0.10 | 3.14E-08 | 0.08 | 0.05 | -0.11 | -0.05 |
| crawls | gross motor | 8.82 | 7.59 | 9.13 | 7.77 | 8.96 | 7.67 | -0.31 | 1.19E-02 | -0.18 | 3.41E-08 | 0.14 | 0.08 | -0.17 | -0.10 |
| transfers an object from one hand to the other | fine motor | 6.13 |  | 6.13 |  | 6.13 |  | 0.00 |  |  |  | 0.00 |  | 0.00 |  |
| taps two objects playfully | fine motor | 8.80 | 8.32 | 8.83 | 8.38 | 8.81 | 8.35 | -0.03 | 6.20E-01 | -0.06 | 3.42E-02 | 0.01 | 0.03 | -0.02 | -0.03 |
| makes repetitive syllables-constant or vowels | language | 7.73 | 6.13 | 7.62 | 6.13 | 7.68 | 6.13 | 0.11 | 3.40E-02 | 0.00 |  | -0.05 | 0.00 | 0.06 | 0.00 |
| gets to sit without support | gross motor | 12.55 | 11.56 | 12.22 | 11.42 | 12.37 | 11.49 | 0.33 | 2.56E-04 | 0.14 | 4.64E-06 | -0.18 | -0.07 | 0.15 | 0.07 |
| pulls to stand | gross motor | 13.28 | 11.75 | 14.15 | 12.02 | 13.58 | 11.85 | -0.87 | 5.72E-04 | -0.27 | 1.00E-04 | 0.30 | 0.10 | -0.57 | -0.17 |
| uses thumb- fingers grasp | fine motor | 10.65 | 9.13 | 9.72 | 9.13 | 10.26 | 9.13 | 0.93 | 4.32E-196 | 0.00 |  | -0.39 | 0.00 | 0.54 | 0.00 |
| feeds self | fine motor | 9.13 |  | 9.13 |  | 9.13 |  | 0.00 |  |  |  | 0.00 |  | 0.00 |  |
| vocalizes in a dialogue | personal-social | 9.13 |  | 9.13 |  | 9.13 |  | 0.00 |  |  |  | 0.00 |  | 0.00 |  |
| understands simple instructions | language | 11.70 | 10.31 | 10.99 | 9.13 | 11.47 | 9.95 | 0.71 | 1.26E-28 | 1.18 | 2.11E-133 | -0.23 | -0.36 | 0.48 | 0.82 |
| responds when addressed by name | personal-social | 9.13 |  | 9.13 |  | 9.13 |  | 0.00 |  |  |  | 0.00 |  | 0.00 |  |
| says one word or pronounces meaningful sounds | language | 13.21 | 11.13 | 12.04 | 10.46 | 12.42 | 10.78 | 1.17 | 5.38E-12 | 0.67 | 1.45E-14 | -0.79 | -0.35 | 0.38 | 0.32 |
| responds differently to familiar and stranger | personal-social | 9.13 |  | 9.13 |  | 9.13 |  | 0.00 |  |  |  | 0.00 |  | 0.00 |  |
| walks with assistance | gross motor | 13.50 | 12.13 | 13.88 | 12.13 | 13.70 | 12.13 | -0.38 | 4.14E-12 | 0.00 |  | 0.20 | 0.00 | -0.18 | 0.00 |
| says 2-3 words | language | 18.29 | 15.05 | 15.60 | 14.25 | 16.54 | 14.72 | 2.69 | 1.29E-15 | 0.80 | 4.85E-25 | -1.75 | -0.33 | 0.94 | 0.47 |
| familiar with at least one body part | language | 19.65 | 17.90 | 18.57 | 17.11 | 18.81 | 17.55 | 1.08 | 7.33E-02 | 0.79 | 9.17E-08 | -0.84 | -0.35 | 0.24 | 0.44 |
| points at familiar objects to request | language | 15.45 | 14.32 | 15.12 | 13.27 | 15.31 | 13.80 | 0.33 | 9.56E-07 | 1.05 | 1.64E-77 | -0.14 | -0.52 | 0.19 | 0.53 |
| expresses will vocally or with gestures | language | 12.13 |  | 12.13 |  | 12.13 |  | 0.00 |  |  |  | 0.00 |  | 0.00 |  |
| makes eye contact and expresses reciprocity during joint game | personal-social | 12.13 |  | 12.13 |  | 12.13 |  | 0.00 |  |  |  | 0.00 |  | 0.00 |  |
| walks without assistance | gross motor | 18.13 |  | 18.13 |  | 18.13 |  | 0.00 |  |  |  | 0.00 |  | 0.00 |  |
| climbs upstairs with assistance | gross motor | 18.13 |  | 18.13 |  | 18.13 |  | 0.00 |  |  |  | 0.00 |  | 0.00 |  |
| builds a tower of cubes | fine motor | 21.02 | 18.13 | 19.91 | 18.13 | 20.52 | 18.13 | 1.11 | 3.08E-07 | 0.00 |  | -0.50 | 0.00 | 0.61 | 0.00 |
| eats independently with a spoon | personal-social | 18.13 |  | 18.13 |  | 18.13 |  | 0.00 |  |  |  | 0.00 |  | 0.00 |  |
| has a vocabulary of over ten words | language | 27.19 | 23.27 | 23.41 | 20.59 | 25.87 | 22.05 | 3.78 | 8.91E-21 | 2.68 | 2.49E-71 | -1.32 | -1.22 | 2.46 | 1.46 |
| composes a sentence of at least two words | language | 30.91 | 27.91 | 28.95 | 24.60 | 30.60 | 27.01 | 1.96 | 2.56E-04 | 3.31 | 9.69E-12 | -0.31 | -0.90 | 1.65 | 2.41 |
| squeezes and sticks out lips to give a kiss | personal-social | 25.19 | 20.45 | 22.47 | 18.13 | 24.05 | 19.21 | 2.72 | 1.43E-08 | 2.32 | 0.00E+00 | -1.14 | -1.24 | 1.58 | 1.08 |
| runs well without falling | gross motor | 24.13 |  | 24.13 |  | 24.13 |  | 0.00 |  |  |  | 0.00 |  | 0.00 |  |
| climbs up and down the stairs without an adult's assistance | gross motor | 24.13 |  | 24.13 |  | 24.13 |  | 0.00 |  |  |  | 0.00 |  | 0.00 |  |
| imitates horizontal, vertical and circle lines | fine motor |  | 29.47 | 32.11 | 24.13 |  | 27.36 |  |  | 5.34 | 6.94E-230 |  | -2.11 |  | 3.23 |
| recognizes familiar objects and pronounces them by name | language | 32.07 | 28.36 | 29.28 | 24.13 | 30.95 | 26.42 | 2.79 | 2.38E-83 | 4.23 | 0.00E+00 | -1.12 | -1.94 | 1.67 | 2.29 |
| understands actions and speech without gestures | language | 34.11 | 29.59 | 30.44 | 24.13 | 31.92 | 27.91 | 3.67 | 3.24E-44 | 5.46 | 0.00E+00 | -2.19 | -1.68 | 1.48 | 3.78 |
| expresses freely | language |  | 34.43 |  | 30.98 |  | 32.27 |  |  | 3.45 | 6.82E-102 |  | -2.16 |  | 1.29 |
| participates in a dialogue | personal-social | 33.42 | 29.32 | 29.94 | 24.13 | 31.61 | 27.72 | 3.48 | 3.58E-94 | 5.19 | 0.00E+00 | -1.81 | -1.60 | 1.67 | 3.59 |
| stands up on one leg for 3 seconds | gross motor | 43.44 | 39.38 | 41.07 | 36.13 | 42.61 | 36.13 | 2.37 | 1.02E-43 | 3.25 | 6.35E-49 | -0.83 | -3.25 | 1.54 | 0.00 |
| jumps from a stair | gross motor | 36.13 |  | 36.13 |  | 36.13 |  | 0.00 |  |  |  | 0.00 |  | 0.00 |  |
| puts on shoes and dresses independently without buttoning | personal-social | 42.12 | 36.13 | 36.13 |  | 37.38 | 36.13 | 6.00 | 2.52E-109 |  |  | -4.74 | 0.00 | 1.25 |  |
| imitates patterns (+) and copies circles | fine motor |  | 42.59 | 41.94 | 36.13 | 44.07 | 40.84 |  |  | 6.47 | 2.63E-198 |  | -1.75 | 2.13 | 4.71 |
| counts three cubes | language |  | 42.15 | 42.95 | 36.41 | 45.17 | 40.35 |  |  | 5.75 | 2.08E-104 |  | -1.80 | 2.22 | 3.94 |
| familiar with at least three prepositions | language |  | 40.99 | 42.76 | 36.13 | 44.95 | 36.79 |  |  | 4.86 | 4.42E-77 |  | -4.20 | 2.19 | 0.66 |
| understandable speech | language |  | 43.06 | 43.27 | 36.13 |  | 39.58 |  |  | 6.94 | 7.00E-138 |  | -3.48 |  | 3.45 |
| jumps on one leg | gross motor | 57.81 | 54.31 | 55.54 | 51.37 | 57.06 | 52.85 | 2.27 | 1.05E-06 | 2.94 | 3.79E-08 | -0.75 | -1.46 | 1.52 | 1.48 |
| walks heel to toe | gross motor | 62.49 | 57.32 | 57.24 | 52.53 | 58.48 | 55.57 | 5.25 | 1.04E-13 | 4.79 | 2.61E-10 | -4.01 | -1.75 | 1.24 | 3.04 |
| dresses independently | personal-social | 52.08 | 48.13 | 48.13 |  | 48.13 |  | 3.95 | 2.34E-08 |  |  | -3.95 |  | 0.00 |  |
| copies geometrical shapes such as X and triangle | fine motor | 65.10 | 58.42 | 57.86 | 55.78 | 62.18 | 57.27 | 7.24 | 6.85E-18 | 2.64 | 4.92E-11 | -2.92 | -1.15 | 4.32 | 1.49 |
| draws a human figure | fine motor | 62.50 | 58.54 | 57.74 | 55.82 | 59.66 | 57.60 | 4.76 | 2.79E-25 | 2.72 | 6.83E-22 | -2.84 | -0.94 | 1.92 | 1.78 |
| plays with peer group | personal-social | 48.13 |  | 48.13 |  | 48.13 |  | 0.00 |  |  |  | 0.00 |  | 0.00 |  |
| uses correct verbs and tense to describe a picture | language | 56.91 | 52.33 | 54.91 | 48.13 | 56.30 | 50.39 | 2.00 | 5.09E-04 | 4.20 | 1.14E-10 | -0.61 | -1.94 | 1.39 | 2.26 |
| answers orientation questions such as name or age | language | 56.77 | 51.17 | 54.50 | 48.13 | 56.33 | 48.44 | 2.27 | 2.68E-05 | 3.04 | 3.04E-05 | -0.44 | -2.73 | 1.83 | 0.31 |

**eTable 3 – Milestones names legend for Figure 2**

| Milestone full name  (THIS developmental scale) | Milestone short name  (Figures 2 & 3) |
| --- | --- |
| 6 weeks to 3 months | |
| Visually follows a moving object horizontally | follows a moving object horizontally |
| Vocalizes in response to human voice | responds to human voice |
| smiles responsively |  |
| raises head |  |
| 3-6 months | |
| Hands together, manipulates fingers | manipulates fingers |
| grasps an object |  |
| Visually follows a moving object vertically | follows a moving object vertically |
| Responds to rattling sound |  |
| makes various sounds including constants (ah-goo) | makes sounds including constants |
| Head and chest up in prone position | head and chest up |
| 6-9 months | |
| transfers an object from one hand to the other | transfers an object |
| makes repetitive syllables-constant or vowels | makes repetitive syllables |
| crawls |  |
| rolls over from abdomen to back and back to abdomen | rolls over |
| taps two objects playfully | taps two objects |
| 9-12 months | |
| gets to sit without support | gets to sit |
| pulls to stand |  |
| uses thumb- finger grasp | thumb- finger grasp |
| feeds self |  |
| vocalizes in a dialogue |  |
| understands simple instructions |  |
| responds when addressed by name | responds to name |
| responds differently to familiar and stranger | differentiates familiar and stranger |
| says one word or pronounces meaningful sounds | says one word |
| 12-18 months | |
| walks with assistance |  |
| says 2-3 words |  |
| familiar with at least one body part | recognizes one body part |
| points at familiar objects to request | points at familiar objects |
| expresses will vocally or with gestures | expresses will vocally |
| makes eye contact and expresses reciprocity during joint game | makes eye contact during play |
| 18-24 months | |
| walks without assistance |  |
| climbs upstairs with assistance |  |
| builds a tower of cubes |  |
| eats independently with a spoon | eats independently |
| has a vocabulary of over ten words | vocabulary of over ten words |
| composes a sentence of at least two words | combines two words |
| squeezes and sticks out lips to give a kiss | kisses |
| 2-3 years | |
| runs well without falling | runs without falling |
| climbs up and down the stairs without an adult's assistance | climbs stairs without assistance |
| imitates horizontal, vertical and circle lines | imitates lines |
| recognizes familiar objects and pronounces them by name | names familiar objects |
| understands actions and speech without gestures | understands actions and speech |
| participates in a dialogue |  |
| expresses freely |  |
| 3-4 years | |
| stands up on one leg for 3 seconds | stands up on one leg |
| jumps from a stair |  |
| puts on shoes and dresses independently without buttoning | puts on shoes |
| imitates patterns (+) and copies circles | imitates patterns and circles |
| counts three cubes |  |
| familiar with at least three prepositions | familiar with prepositions |
| understandable speech |  |
| 4-6 years | |
| jumps on one leg |  |
| walks heel to toe |  |
| dresses independently |  |
| copies geometrical shapes such as X and triangle | copies geometrical shapes |
| draws a human figure |  |
| plays with peer group |  |
| uses correct verbs and tense to describe a picture | uses correct verbs and tense |
| answers orientation questions such as name or age | answers orientation questions |
